# The cost of the COVID-19 pandemic vs the cost-effectiveness of mitigation strategies in the EU/UK/EEA and OECD countries: a systematic review

**DOI:** 10.1101/2022.05.31.22275813

**Authors:** Constantine Vardavas, Konstantinos Zisis, Katerina Nikitara, Ioanna Lagou, Katerina Aslanoglou, Kostas Athanasakis, Revati Phalkey, Jo Leonardi-Bee, Esteve Fernandez, Orla Condell, Favelle Lamb, Frank Sandmann, Anastasia Pharris, Charlotte Deogan, Jonathan E. Suk

## Abstract

**Objectives:** COVID-19 poses a threat of loss of life, economic instability, and social disruption. We conducted a systematic review of published economic analyses to assess the direct and indirect costs of the SARS-CoV-2 pandemic, and to contrast these with the costs and the cost-benefit of public health surveillance, preparedness, and response measures in averting and/or responding to SARS-CoV-2 pandemic.

**Setting:** A systematic literature review was conducted to identify peer-reviewed articles estimating the cost of the COVID-19 pandemic and the cost-effectiveness of pharmaceutical or non-pharmaceutical interventions in EU/EEA/UK and OECD countries, published from the 1st of January 2020 through 22nd April 2021 in Ovid Medline and EMBASE. The cost-effectiveness of interventions was assessed through a dominance ranking matrix approach. All cost data were adjusted to the 2021 Euro, with interventions compared with the null.

**Primary and secondary outcome measures:** Direct and indirect costs for SARS-CoV-2 and preparedness and/or response or cost-benefit and cost-effectiveness were measured.

**Results:** We included data from 41 economic studies. Ten studies evaluated the cost of COVID-19 pandemic, while 31 assessed the cost-benefit of public health surveillance, preparedness, and response measures. Overall, the economic burden of SARS-CoV-2 was found to be substantial for both the general population and within specific population subgroups. Community screening, bed provision policies, investing in personal protective equipment and vaccination strategies were cost-effective, in most cases due to the representative economic value of below acceptable cost-effectiveness thresholds. Physical distancing measures were associated with health benefits; however, their cost-effectiveness was dependent on the duration, compliance and the phase of the epidemic in which it was implemented.

**Conclusions:** SARS-CoV-2 is associated with substantial economic costs to healthcare systems, payers, and societies, both short term and long term, while interventions including testing and screening policies, vaccination and physical distancing policies were identified as those presenting cost-effective options to deal with the pandemic, dependent on population vaccination and the Re at the stage of the pandemic.

## INTRODUCTION

In the aftermath of significant outbreaks since the beginning of the 21st century, including among others those of Ebola, avian influenza (H5N1, H7N9), the 2009 H1N1 influenza virus pandemic, Severe Acute Respiratory Syndrome (SARS) and the Middle East Respiratory Syndrome Coronavirus (MERS-CoV), it was acknowledged that large-scale infectious disease outbreaks represent a menace for loss of life, economic disturbance, and social disruption (1). The most recent pandemic caused by SARS-CoV-2 has undoubtedly hit hard on societies and healthcare systems. According to the situation update conducted by ECDC, as of week 17 of 2022, 512 690 034 cases of COVID-19 have been reported, including 6 252 316 deaths (2). To mitigate the pandemic and given the lack of preventative treatments until the rollout of vaccines in late 2020, governments worldwide implemented various non-pharmaceutical interventions (NPIs). These measures included personal protective, environmental, and physical distancing strategies as well as travel restrictions (3) which have been loosened and reinforced depending on the variation of the epidemiological situation (4), leading to an unprecedented decline in global economic activities and an economic burden both from direct costs of the NPIs and indirect costs. The closure of businesses led to significant disruptions to the global value chains in the first quarter of 2020 through increased unemployment rates, revenue loss, and a sharp decrease in personal incomes while the earlier action of NPIs implementation contributed to reduced economic impact as was shown in a study assessing the short-term economic consequences of the first-wave Covid-19 pandemic (5).

In the US, the cost of the COVID-19 pandemic has been estimated at $13tn for the first 20 weeks of the pandemic (90% of the country’s annual GDP) when drastic NPIs were predominately implemented, and approximately half of that figure corresponds to the projected 10-year decline in GDP attributable to the pandemic (6). Additionally, the global cost of the COVID-19 crisis has been estimated at 14% of 2019 GDP (around 12,206 mm$), while for Europe, it is even higher: 16% for the Eurozone and 19% for the UK (7). Considering the cost-effectiveness of NPIs, in a recent systematic review and meta-analysis conducted by Zhou et al. (2021), the pooled incremental net benefit (INB) for NPIs was estimated at $972·05, with subgroup analyses indicating the highest pooled INBs were for screening ($2390·89) and suppression ($2156·00) interventions (8).

Robust national preparedness and response strategies require recent data on the health impacts and the economic burden of respiratory infectious disease outbreaks in contrast to those associated with emergency response and preparedness actions. This evidence will ensure well-informed decisions regarding the proper allocation of resources (9, 10), information relevant not only for the COVID-19 pandemic but also for future pandemics.

This systematic review aims to a) summarise the total direct and indirect costs of the COVID-19 pandemic across EU/EEA/UK and OECD countries, and b) to contrast with the costs and the cost-benefit of public health surveillance, preparedness, and response measures in averting and/or responding to COVID-19 pandemic in this setting.

## METHODS

### Search strategy and selection criteria

A systematic review was conducted to identify peer-reviewed articles published from the 1^st^ January 2020 through 22^nd^ April 2021 in Ovid Medline and EMBASE. The review is reported adhering to the PRISMA framework (Preferred Reporting Items for Systematic Reviews and Meta-analyses) (11) and the Consolidated Health Economic Evaluation Reporting Standards (CHEERS) (12). Two sets of inclusion criteria were used to determine the eligibility of the studies based on the two objectives; however, a single search strategy was used to capture eligible studies. The complete search strategy and search terms is available in **Online Supplementary Appendix 1**.

The inclusion criteria for Objective 1: To summarise the total direct and indirect costs of the COVID-19 pandemic) were as follows:

✓ Population: EU/EEA/UK and remaining OECD countries
✓ Exposure: COVID-19 outbreak or public health preparedness measures or interventions.
✓ Outcome measures: direct and indirect costs for disease and preparedness and/or response.
✓ Perspective: All direct and indirect costs were considered pertaining to all relevant perspectives (e.g., individual, health and social care, and societal-including national and regional).
✓ Study designs: We included all relevant analytical epidemiological designs which estimate cost, including partial cost evaluation studies, cost studies, cost-outcome description studies, cost-description studies, and economic modelling studies.

The inclusion criteria for Objective 2: To contrast with the costs and the cost-effectiveness of public health surveillance, preparedness, and response measures in averting and/or responding to COVID-19 were as follows:

✓ Population: EU/EEA/UK and remaining OECD countries
✓ Intervention: Public health preparedness measures or interventions.
✓ Comparator: (i) No intervention (cost of inaction) or current practice; (ii) cost of preparedness vs cost of response (for studies reporting cost and benefit of public health preparedness).
✓ Outcome measures: Cost-benefit and cost-effectiveness outcomes (e.g., cost per life-year saved). Studies that use other methods to formally combine cost and clinical outcome data (pertinent to infectious diseases) were also included. Typical outcome measures of economic evaluations include: Life years gained (or cost per life-year achieved with the intervention under investigation when incremental costs are combined), Quality-Adjusted Life Years (cost per QALY gained), Cases averted (e.g., cost per case that is averted with the intervention versus the comparator) and monetary outcomes (in the case of a cost-benefit analysis). Where available the incremental cost-effectiveness ratio (ICER) was also provided.
✓ Perspective: All direct and indirect costs were considered pertaining to all relevant perspectives (e.g., individual, health and social care, and societal-including national and regional).
✓ Study designs: We included all designs specific to cost benefit questions either as full economic evaluation studies, including cost-minimization, cost-effectiveness, cost-utility and cost-benefit analyses; cost-outcome and economic modelling studies; or partial economic evaluations

### Data analysis and extraction

Studies identified from the searches were uploaded into a bibliographic database (Covidence) in which duplicate entries were removed. Initially, a random sample of 100 titles and abstracts were screened independently for eligibility by two reviewers to enable consistency in screening and identify areas for amendments in the inclusion criteria. Since a high measure of inter-rater agreement was achieved (inter-rater agreement = 88.7%), the remaining titles and abstracts were screened for eligibility by one reviewer. Full-text articles of potentially eligible studies were retrieved and screened independently by two reviewers (inter-rater agreement = 89.3%). Data were extracted with the use of a predefined data extraction sheet. Initially, two reviewers piloted the data extraction template independently on a random sample of five included studies, and given the high consistency in data extraction, the remaining studies were extracted only by one reviewer. Disagreements in every step of the process were subsequently discussed with a third reviewer and agreed upon.

### Appraisal of methodological quality

The Consensus on Health Economic Criteria (CHEC) checklist was used (13) to evaluate the methodological quality of full health economic evaluations, which comprises of 19 questions with answers of “Yes” or “No”. For each positive response of full health economic evaluations, a single point was assigned for the methodological quality, with a maximum score of 19. For the quality appraisal of partial economic evaluations, we used items from the CHEC checklist that was applicable – hence the maximum score was 17. In cases of insufficient information or details reported in both full and partial economic evaluation studies, with regard to a specific item, no point was awarded for that question. Studies were categorised as high (>80%), good (60%–80%), medium (40%–60%), or low (<40%) quality. The quality appraisal process was completed by one reviewer, since piloting of three studies by two independent reviewers had an inter-rater agreement of 85.6%.

### Comparative economic analysis approach

All cost data were adjusted to a common currency (Euro in 2021) and price year, using the Campbell and Cochrane Economics Methods Group–Evidence for Policy and Practice Information and Coordinating Centre cost converter (13). A two-stage computation is used, where the 2021 implied conversion factor is US $1 = € 0.88. The 2021 implied conversion factor of British pounds is £1 = € 1.18. In our study, we first inflated the cost from the original price year to 2021, using a Gross Domestic Product deflator index (GDP values), obtained from the International Monetary Fund World Economic Outlook Database GDP deflator index data set. After that, we converted the original currency to the next rounded 2021 Euros, using conversion rates based on Purchasing Power Parities for GDP (PPP values). For studies that did not state the year of cost calculation, the costs were calculated one year before the publication year.

### Synthesis of cost-effectiveness

The Dominance Ranking Matrix (DRM) was used to assess the cost-effectiveness of the interventions noted within the identified studies (14). The DRM is a three-by-three matrix with the following classification options:

a. Strong dominance for the intervention when the incremental cost-effectiveness measure shows the intervention vs. the comparator is (i) more effective and less costly; or (ii) as effective and less costly; or (iii) equally costly and more effective. Strong dominance suggests that under the similar circumstances, the intervention is preferable to the comparator.
b. Weak dominance for the intervention vs. the comparator is notd when the measure shows the intervention as (iv) equally costly and effective; or (v) more effective and more costly; or (vi) less effective and less costly. In principle weak dominance indicates that no conclusion can be drawn and hence judgment by policymakers is required on whether the intervention is preferable when taking into consideration whether the cost/benefit tradeoffs are worth the introduction of the intervention within the particular context.
c. Non-dominance for the intervention vs. the comparator when the measure shows the intervention as (vii) more costly and less effective ; or (viii) equally as costly and less effective; or (ix) more costly and as effective. In this case the comparator is preferable to the measured intervention, at least under the circumstances of the specific study.

### Patient and public involvement

This study was performed under contract for the European Center for Disease Prevention and Control (ECDC). Patients or the public were not explicitly involved in the design, or conduct, or reporting, or dissemination plans of our research.

## RESULTS

10,314 non-duplicate studies were identified, of which 403 proceeded to full text screening with362 full text studies were excluded for the following reasons: inadequate data on costs and/or cost-effectiveness (n=269), reviews/editorials/perspectives/views/opinion papers (n=68), not referring to outbreaks of Covid-19 infectious disease (n=23), and no full text available (2). Subsequently, 41 studies met all inclusion criteria and were included in the systematic review (**Figure 1**).

**Figure 1.**
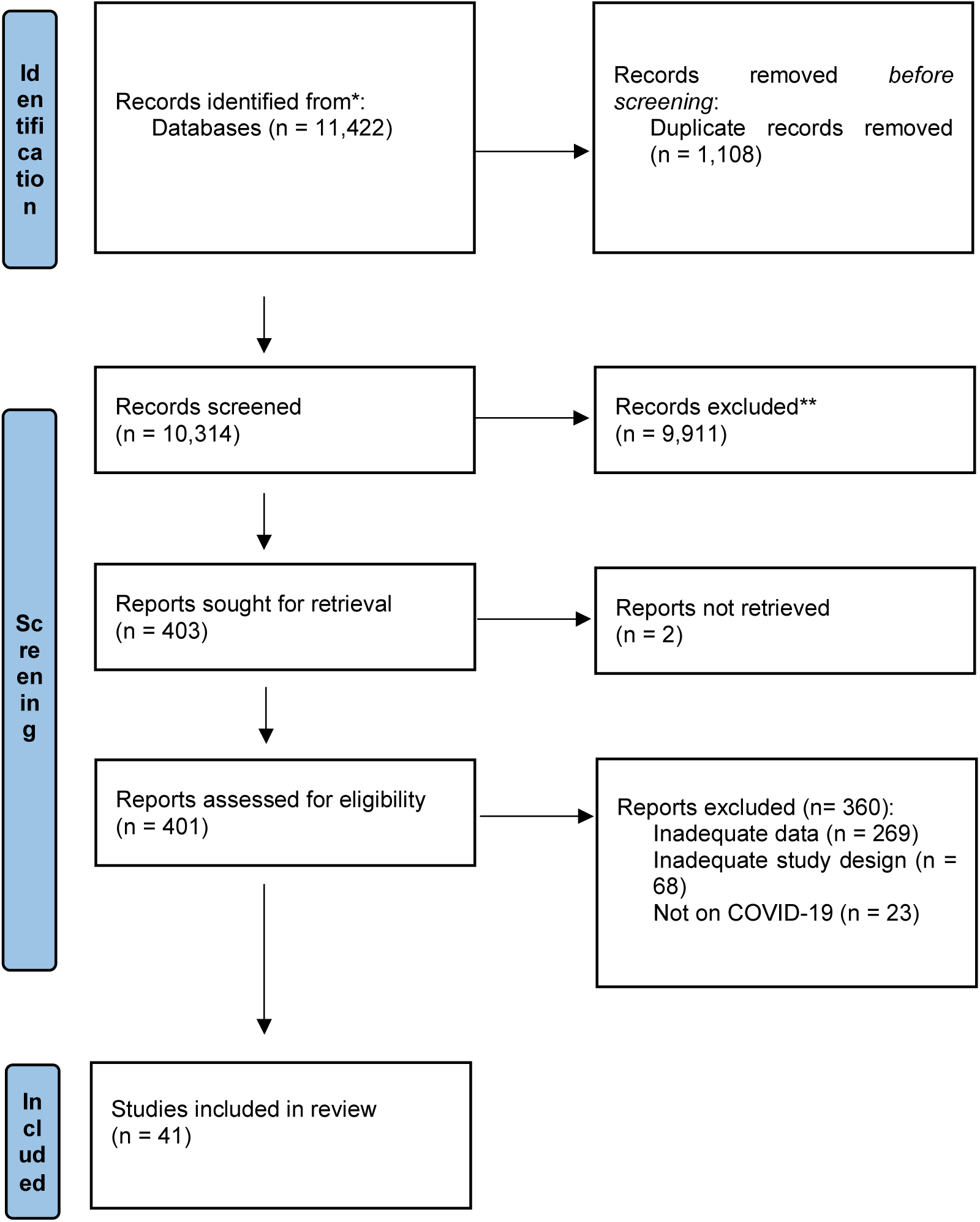
PRISMA flow chart of the search strategy.

Approximately half of the studies (20/41) were of high methodological quality, ten had good quality, and the remaining 11 studies were of medium quality (**Online Supplementary Appendix 2)**. Lower scores were mainly due to missing information relating to the comparative intervention, lack of sensitivity analysis, not reporting incremental costs among comparative interventions (since in some studies alternative strategies were not included).

### Objective 1: Economic impact of COVID-19 infection to healthcare systems and societies

Ten studies evaluated the cost of the COVID-19 pandemic within EU/EEA/UK and OECD countries (**Table 1)**, among which eight studies used a cost of illness analysis approach (15–22). The direct medical costs were presented in seven out of ten studies (15-17, 19, 20, 22, 23), while in eight studies the costs were estimated for a time horizon shorter than one year (16-18, 20-24), and specific discount rates were not applied in seven studies (15-17, 20, 22-24). With regards to the country of origin, five out of the ten studies took place in the European region, including in the UK and Turkey (15–19), three were from the USA (20–22), one from Korea (23) and one from Australia (24). Six of the 10 studies were observational (17-20, 22, 23), while a few used modelling techniques (15, 16, 24) or simulation methods (21). Furthermore, it should be noted that the healthcare system perspective was included in the analysis of four studies (15, 16, 21, 22), the societal in two (17, 18), the third payer perspective was assessed in three (19, 21, 23), and one study (24) assessed data from the public health payer perspective. The perspective considered within onestudy was unclear (20).

**Table 1.**
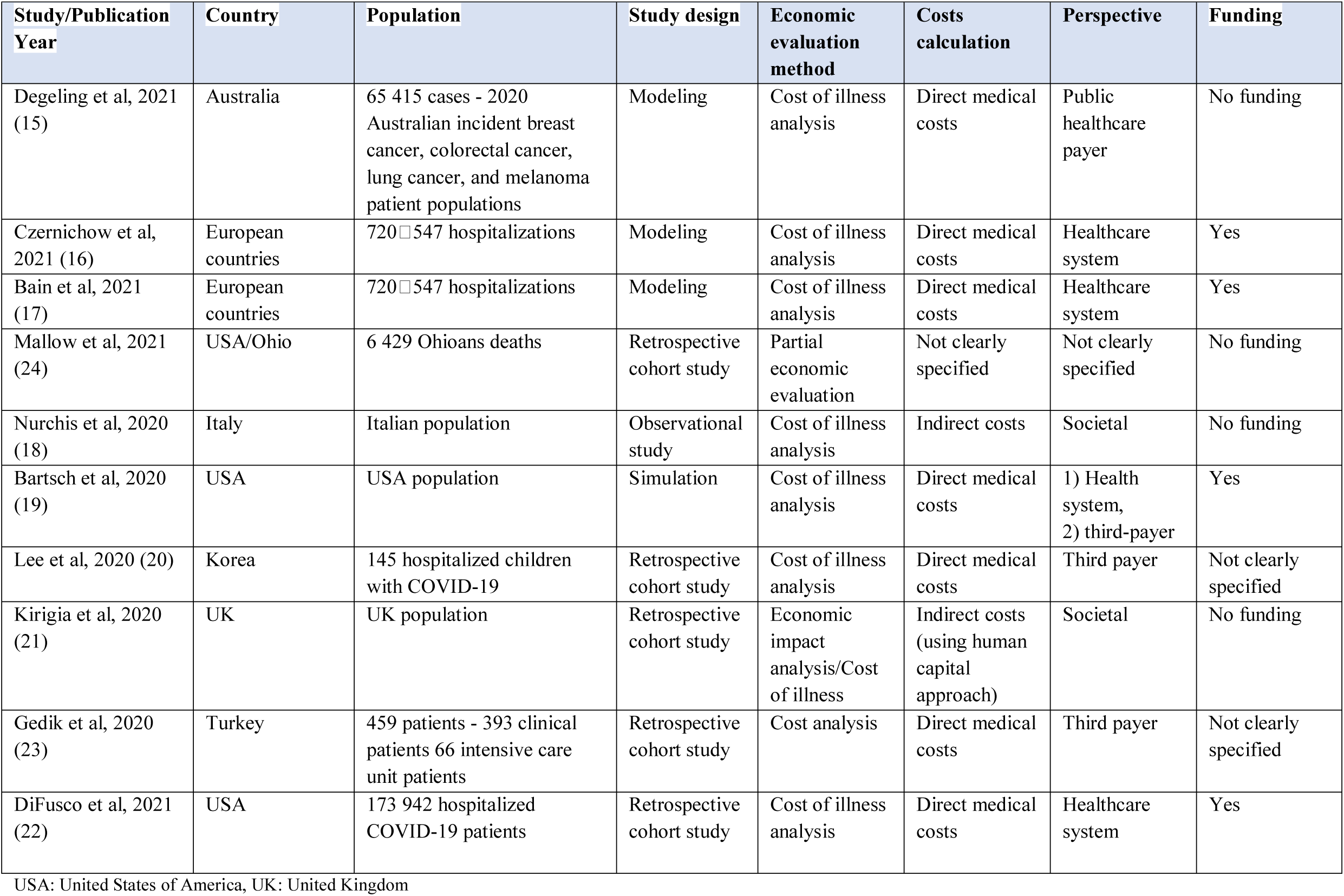
Overview of included studies evaluating the economic impact of COVID-19 pandemic in EU/EEA/UK and OECD countries (n=10)

#### Overall economic burden on the population and to the healthcare system

Overall, the economic burden of COVID-19 pandemic was found to be substantial (**Table 2**). The socioeconomic implications of COVID-19 in Italy, appraised by Nurchis et al. (2020) (18) through a cost of illness analysis of indirect costs, showed a temporary productivity loss reaching 114,120,531 € for 110,868 cases (1,029 € per case). Individuals at the age of > 40 years old were found to be affected the most. Furthermore, the permanent productivity losses were estimated at 333,063,591 € for 3,926 deaths (84,836 € per death) with ages >50 years old to consistently having higher indirect costs due to death. Bartsch et al. (2020) (19) at the start of the pandemic, when no vaccines were available showed that the more people infected, the higher medical costs are presented using aa Monte Carlo simulation model to highlight the devastating impact of the pandemic on the US healthcare system and third payers. In the case of 20% of the US population to be infected with COVID-19 over the course of the pandemic (and not accounting for reinfections), the total costs accrued were estimated at 129.8 billion € and reaching 170.3 billion € including post discharge costs after one year. In the case that 80% of the US population is infected, the direct medical costs reached an estimated 519.4 billion €, and 682.6 billion € with the post discharge costs after one year of infection. Mallow et al. (2021) (24) assessed the value of statistical life in a total of 6 429 deaths in Ohio USA, where the economic burden of premature deaths was estimated at 13.8 billion € as of 30 November 2020. Additional analyses have been performed by Kirigia et al. (2020) (21) using the human capital approach to estimate the total present value of human lives lost due to COVID-19 in the UK as of July 2, 2020. Notably, the value for 43 906 lives lost approached 7.8 billion € at a 3% discount rate with, approximately 76.2% of the total present value sustained by those aged 30 - 79 years. Lee et al. (2020) (20) aimed to determine the hospitalisation periods and medical costs among 145 hospitalised children with COVID-19 in Korea. According to the results, the estimated medical expenses reached 252,389 € totally and increased per age as 54 patients in the ages of 16-19y accounted for 156,738 € (more than 60% of the total cost) and per patient at 2 903€ for a mean hospitalization period of approximately 10 days, indicating that these ages contributed to higher costs than younger ages included in the study.

**Table 2.** Econometric outcomes of studies ealuating the economic impact of COVID-19 in the EU/EEA/UK and OECD countries (n=10)

| Study/<br>Publication<br>Year | Time horizon | Discounting | Modeling -<br>uncertainty<br>assessment | Economic evaluation outcomes, €, 2021 | Uncertainty assessment results, €, 2021 |
| --- | --- | --- | --- | --- | --- |
| Degeling<br>et al, 2021 | 5 and 10-year | Not applied | Inverse<br>stage/shift<br>model<br>Uncertainty<br>assessment<br>Not applied | **Incremental cost – 3 month delay**<br>Breast cancer: 1 09 million €<br>Coloctreral cancer: 227 082 €<br>Lung cancer: -<br>Melanoma: 5 02 million €<br>**Incremental cost – 6 month delay**<br>Breast cancer: 4 25 million €<br>Coloctreral cancer: 895 070 €<br>Lung cancer: -<br>Melanoma: 20 1 million € | Not applied |
| Czernichow<br>et al, 2021 | 6 months | Not applied | Healthcare<br>cost model<br>Uncertainty<br>assessment<br>Not applied | **Costs per average hospital admission - By BMI - Average (weighted for all countries)**<br>2) BMI <25: 16 173 €<br>3) BMI 25–29.9: 17 389 € - 24 324 €<br>4) BMI 30–34.9: 24 324 €<br>5) BMI 35–39.9: 25 409 €<br>6) BMI ≥40: 31 652 €<br>**Total direct costs of secondary care of COVID-19 – by BMI**<br>1) Totally (720 547 hosp. people): 14 2 billion €<br>2) BMI <25 (211 407 hosp. people): 3 37 billion €<br>3) BMI 25–29.9 (259 764 people): 4 5 billion €<br>4) BMI 30–34.9 (174 278 hosp. people): 4 2 billion €<br>5) BMI 35–39.9 (54 673 hosp. people): 1 38 billion €<br>6) BMI ≥40 (20 424 hosp. people): 664 million € | Not applied |
| Bain<br>et al, 2021 | 6 months | Not applied | Healthcare<br>cost model<br>Uncertainty<br>assessment<br>Not applied | **Total direct costs of secondary care of COVID-19**<br>1) Totally (720 547 hosp. cases): 14 2 billion €<br>2) With diabetes (95 255 hosp. cases): 10 8 billion €<br>3) Without diabetes (625 292 hosp. cases): 3 34 billion €<br>**Total direct costs of secondary care of COVID-19 – per diabetes type**<br>1) <u>Type 1</u><br>a) Good glycaemic control (2 656 hosp. cases): 78 6 million € | **Lower and upper bound estimations**<br>The range for total direct secondary care costs is 11 3 billion € - 21 8 billion € |
|  |  |  |  | b) Poor glycaemic control (6 869 hosp. cases): 400 4 million €<br>2) <u>Type 2</u><br>a) Good glycaemic control (54 028 hosp. cases): 1 367 billion<br>b) Poor glycaemic control (31 701 hosp. cases): 1 498 billion |  |
| Mallow et al, 2021 | From the beginning of Covid-19 infection as of 30 November 2020 in Ohio | Not applied | Not applied<br>Uncertainty assessment<br>Not applied | **Value of statistical life**<br>Low: 15 492 826 678 €<br><sup>1</sup> Base: 13 814 246 372 €<br>High: 17 979 395 429 €<br>**Value of statistical life, per sex**<br>Male: 7 443 681 213 €<br>Female: 6 351 426 503 €<br>Unknown: 19 138 654 €<br>**Value of statistical life, per age**<br>Less than 60 y: 3 092 373 227 €<br>60 y or older: 10 720 747 342 €<br>Unknown: N/A | Not applied |
| Nurchis et al, 2020 | Approximately 3 months | 3% for the individual future annual wages | Not applied<br>Uncertainty assessment<br>Not applied | **Temporary productivity loss cost**<br>Totally (110 868 cases): 114 120 531 € or 1 029 € per case<br>20-29 y (10 369 cases): 9 191 575 €<br>30-39y (14 858 cases): 15 088 029 €<br>40-49y (25 420 cases): 25 813 548 €<br>50-59y (35 068 cases): 37 284 537 €<br>60-69y (25 153 cases): 26 742 841 €<br>**Permanent productivity loss cost**<br>Totally (3 926 deaths): 333 063 591 € or 84 836 € per death<br>20-29 y (8 deaths): 1 755 371 €<br>30-39y (49 deaths): 10 854 412 €<br>40-49y (224 deaths): 40 636 472 €<br>50-59y (918 deaths): 122 559 274 €<br>60-69y (227 deaths): 157 258 062 € | Not applied |
| Bartsch et al, 2020 | Incurred during the course of the infection and postdischarge for one year | 3% for costs | Monte Carlo simulation<br>Uncertainty assessment:<br>Scenarios and sensitivity analyses by | **Direct medical cost, per case**<br>1) A person with symptomatic infection, i) including only costs during the course of the infection, ii) one year after postdischarge:<br>Median: 2 418 € (95% UI 2 282 € - 2 545 €)<br>Median: 3 192 € (95% UI 3 017 € - 3 336 €)<br>2) A person with only mild symptoms by age group, including only costs during the course of the infection:<br>a) 0-17 y - Median: 72 € (95% UI 48 € - 98 €)<br>b) 18-64 y - Median: 45 € (95% UI 43 € - 53 €) | **20% attack (infection) rate:**<br>Decreasing post-discharge costs by 50 percent would lower that amount to a median of 149 8 billion € [95% UI: 142 0 €, 158 7 billion €].<br>Decreasing the currently reported values for the probability of death by a relative 95 percent had little impact on direct medical costs - median: 170 7 billion € [95% UI: |
|  |  |  | varying several key parameters to determine their impact on our results | c) 65 y or older Median: 76 € (95% UI 71 € - 82 €)<br>3) A person with symptomatic infection requiring hospitalization, by age group - including only costs during the course of the infection:<br>a) 0-17 y - Median: 9 027 € (95% UI 7 203 – 10 991 €)<br>b) 18-44 y - Median: 10 429 € (95% UI 9 141 € - 11 707 €)<br>c) 45-64 y - Median: 12 661 € (95% UI 11 451 € - 13 943 €)<br>d) 65-84 y - Median: 12 661 € (95% UI 11 261 € - 12 340 €)<br>e) 85 y or older - Median: 9 451 € (95% UI 8 807 € - 10 193 €)<br>**Direct medical costs,**<br>i) incurred during the course of the infection,<br>ii) post discharge **(with current reported values) - Attack (infection) rates in US**<br>1) 20% infection rate: 129 8 billion (95% UI 122 7 – 137 5), ii) 170 3 billion (95% 160 7 –181)<br>2) 50% infection rate: i) 324 6 billion (95% UI 306 7 – 344 2), ii) 426 3 billion (95% 403 1 –453 3)<br>3) 80% infection rate: i) 519 4 billion (95% UI 489 05 – 550), ii) 682 6 billion (95% 642 8 – 724 05) | 161 06 billion €, 180 4 €].<br>**50% attack (infection) rate:**<br>Reducing the post-discharge costs by 50 percent decreased the median direct medical costs to 375 2 billion € [95% UI: 355 billion €, 398 1 billion €].<br>**80% attack (infection) rate:**<br>If post-discharge costs were 50 percent lower, median costs would be reduced from 682 7 billion € to 600 5 billion € [95% UI: 565 8 billion €, 637 4 billion €]. The decrease of the probability of severe disease leads to decreased costs by a relative 49 7%, to 261 2 billion €. Decreasing the reported value for the probability of death by a relative 95% had no impact on cost when costs that might be incurred after discharge were included[median: 682 7 billion €; 95% UI: 646 2 billion €, 722 2 billion €] |
| Lee et al, 2020 | 2 months | Not applied | Not applied<br>Uncertainty assessment<br>Not applied | **Medical costs, per age-group and mean hospitalization days**<br>a) Totally (145 patients): 252 389 € for 10.38 days (95% UI 8 38 – 12 38 hospitalization days)<br>Per patient: 1.740 6 €<br>b) 0-5y (57 patients): 33 927 € for 4.63 days (95% UI 2 99 – 6 27 hospitalization days)<br>Per patient: 594 8 €<br>c) 6-10y (15 patients): 30 786 € for 10.87 days (95% UI 6 85 – 14 89 hospitalization days)<br>Per patient: 2.051 8 €<br>d) 11-15y (19 patients): 30 937 € for 14.88 days (95% UI 8 86 – 14 72 hospitalization days)<br>Per patient: 1.628 05 € | Not applied |
|  |  |  |  | e) 16-19y (54 patients): 156 738 € for 15.81 days (95% UI 11 42 – 20 20 hospitalization days)<br>Per patient: 2.902 7 € |  |
| Kirigia et al, 2020 | Onset and as of July 2020 | 3%, 5% and 10% | Not applied<br>Uncertainty assessment<br>Not applied | **Present value of human lives lost, at 3%, 5% and 10% discount rate**<br>1) Totally (43 906 lives lost): 7 849 137 714 €, 6 929 149 668 €, 5 419 985 376 €<br>2) 0-9y: -<br>3) 10-19y: 10 034 444 €, 6 929 149 668 €, 3 486 414 €<br>4) 20-29y: 61 632 496 €, 42 586 289 €, 22 600 723 €<br>5) 30-39y: 462 939 584 €, 113 659 147 €, 62 493 943 €<br>6) 40-49y: 462 939 584 €, 348 997 800 €, 202 697 939 €<br>7) 50-59y: 1 272 389 584 €, 1 016 620 912 €, 641 312 072 €<br>8) 60-69y: 1 950 013 557 €, 1 669 784 612 €, 1 188 059 923 €<br>9) 70-79y: 2 140 118 842 €, 1 987 634 724 €, 1 672 314 872 €<br>10) 80 & older: 1 793 826 428 €, 1 743 147 320 €, 1 627 019 486 € | Not applied |
| Gedik et al, 2020 | Approx. 2 months | Not applied | Not applied<br>Uncertainty assessment<br>Not applied | **Mean cost, per patient**<br>Clinical patients: 700 26 € (SD 529 96 €) for 8.97 (SD 4 64) mean hospitalization days<br>One-day hospitalization: 82 05 € (SD 61 58 €)<br>Intensive care unit patients: 2 322 € (SD 1 864 €) for mean hospitalization days of 14 74 (SD 13.19)<br>One-day hospitalization: 188 01 € (SD 75 02 €) | Not applied |
| DiFusco et al, 2021 | 8 months | Not applied | Not applied<br>Uncertainty assessment:<br>A sensitivity analysis using the U07.1 ICD-10 code in the primary position only | **Direct costs (hospital costs)**<br>1) Overall study population (n= 173 942) for 8.3 mean LOS: 19 513 € (mean), 32 545 € (SD)<br>2) Without ICU or IMV (n= 128 063) for 6.1 mean LOS: 11 376 € (mean), 13 302 € (SD)<br>3) With ICU but without IMV (n= 16 496) for 9.6 mean LOS: 20 400 € (mean), 21 850 € (SD)<br>4) Without ICU but with IMV (n= 7 751) for 12.1 mean LOS: 33 171 € (mean), 40 810 € (SD)<br>5) With ICU and IMV (n= 21 632) for 18.6 mean LOS: 62 139 € (mean), 64 821 € (SD) | The sensitivity analysis of hospitalized COVID-19 patients identified 106 801 patients, representing 61.4% of the hospitalized COVID-19 patients included in the main analyses. Mean hospital charges: mean [SD]: 61 484 € - 108 472 € - consistent with the main analyses but slightly lower Mean hospital costs: mean [SD]: 16 834 € - 28 588 € - slightly lower than in the main analyses |
|  |  |  |  | **Direct costs (hospital charges)**<br>1) Overall study population (n= 173 942) for 8.3 mean LOS:<br>72 132 € (mean), 126 212 € (SD)<br>2) Without ICU or IMV (n= 128 063) for 6.1 mean LOS: 40 157 € (mean),<br>43 931 € (SD)<br>3) With ICU but without IMV (n= 16 496) for 9.6 mean LOS: 75 879 €<br>(mean), 79 852 € (SD)<br>4) Without ICU but with IMV (n= 7 751) for 12.1 mean LOS: 125 887 €<br>(mean), 156 833 € (SD)<br>5) With ICU and IMV (n= 21 632) for 18.6 mean LOS: 239 308 € (mean),<br>260 664 € (SD) |  |

BMI: Body mass index, US: United States, y: years, n: number, N/A: Not applicable SD: Standard deviation, ICU: Intensive Care Unit, IMV: Invasive mechanical ventilation, LOS: Length of stay
1 The base case is the best estimate of YPLL and associated value of statistical life years lost. The low and high represent the estimated range

DiFusco et al. (2021) (22) by conducting a cost of illness analysis to estimate the direct medical costs of inpatient setting for a time horizon of 8 months, showed that the mean total of hospital costs related to COVID-19 was 19,513 € for 8.3 mean hospital length of stay days (LOS) (SD 9.1). Higher costs were presented for those people treated in the ICU, reaching a mean cost of 20,400€, which represent 21,850€ in case of ICU usage without invasive mechanical ventilation (IMV) for a mean LOS of 9.1 days and estimated at a mean cost of 62,139€ if both ICU and IMV are used for mean LOS of 18.6 days.

Moreover, Gedik et al. (2020) (23) performed a simple cost-analysis of COVID-19 patients in Turkey and showed that the mean cost per ICU patient was much higher than clinical patients in wards and particularly estimated at 2,322 € for mean hospitalization days of 14.7 compared to 700. € for 9 mean hospitalization days.

#### Overall economic burden to specific population subgroups

With regards to the costs for specific sub-populations, Degeling et al. (2021) (15) estimated the health and economic impacts of delays in treatment initiation of 65,415 cancer cases (breast cancer, colorectal cancer, lung cancer and melanoma) due to COVID-19 infection in Australia. Apart from the excess deaths and life-years lost, costs to the health care system exceeded more than 6 million € for the 3-month delay while more than 25 million € for a 6-month delay. Czernichow et al. (2021) (16) identified a strong relationship between obese and overweight patients (BMI 25.0 to ≥40) with increased direct medical costs of secondary care related to COVID-19 across the EU, UK and EFTA countries, with 44% of the total treatment costs of COVID□19 in Europe to be associated with those populations (due to the higher probability of being hospitalized, a longer length of stay, and higher risk of severe outcomes). The total costs of 14.2 billion € were the total secondary medical care costs for a 6-month time horizon analysis, with cases with a BMI ≥40 accounting for the highest direct costs per case. In a similar study conducted by Bain et al. (2021) (17), whose aim was to estimate the impact of diabetes on the total secondary care costs of COVID-19 and for the same timeframe and population numbers in regards to the same European countries, poor glycemic control was associated with excess direct medical costs of secondary care due to COVID-19 infection, estimated at 400.4 million € for 6 869 hospitalizations of Type 1 diabetes cases and 1,498 billion € for 31 701 hospitalizations of Type 2 diabetes cases.

### Objective 2: Economic evaluation of strategies for the mitigation of COVID-19 virus transmission

Our systematic review identified 31 studies assessing the cost-effectiveness of interventions for reducing SARS-CoV-2 transmission within EU/EEA/UK and OECD countries (**Table 3**). We identified studies that assessed isolation, lockdown policies, physical distancing scenarios (25–35), testing/screening policies (36–42), personal protective equipment intervention (43), vaccinations (44–47) and pharmaceutical treatment strategies (n=4) (47–50). Multiple strategies were evaluated by four studies (51–54), that mainly assessed the combination of testing, isolation, and vaccinations, and one study (55) analysed an ICU bed provision scenario.

**Table 3.** Study characteristics of studies assessing the cost-effectiveness of strategies for mitigating the COVID-19 pandemic (n=31)

| Study/<br>Publication<br>Year, | Funding | Country* and<br>Population | Study design | Perspective | Intervention/strategy |
| --- | --- | --- | --- | --- | --- |
| Pandey et al, 2021 (54) | Yes | USA<br>328 2 million (US population) | Modeling and simulation | Societal | 1. Daily test plus 1-week isolation, 2. Daily test plus 2-week isolation, 3. Test every 7 days plus 1-week isolation, 4. Test every 7 days plus 2-week isolation, 5. Test every 14 days plus 1-week isolation, 6. Test every 14 days plus 2-week isolation, 7. Test every 28 days plus 1-week isolation, 8. Test every 28 days plus 2-week isolation |
| Lee et al, 2021 (48) | Yes | USA<br>327 167 434 persons (US population) | Modeling and simulation | Third payer and societal perspective | Starting treatment (medication of vaccination) |
| Kouidere et al, 2021 (35) | Not mentioned | Worldwide<br>Diabetics | Modeling and simulation | Not clearly mentioned | 1. Quarantine,<br>2. Sensitization and prevention,<br>3. Awareness Program and Quarantine |
| Bastos et al, 2021 (39) | Yes | Worldwide<br>7 169 Participants (inpatients and outpatients) | Systematic review and meta-analysis | Societal | 1. Saliva sampling (collection of specimens by saliva) VS<br>2. nasopharyngeal swab (collection of specimens by nasopharyngeal swab) |
| Risko et al, 2020 (43) | No funding | Low- and middle-income countries<sup>1</sup><br>166 689 862 simulated healthcare worker cases | Modeling and simulation | Societal | 1. Unmitigated (low PPE),<br>2. Unmitigated (full PPE),<br>3. Mitigated (low PPE),<br>4. Mitigated (full PPE),<br>5. Suppressed (low PPE),<br>6. Suppressed (full PPE) |
| Stevenson et al, 2021 (42) | Yes | UK<br>Patients admitted to hospital | Modeling and simulation | Healthcare system | 30 testing strategies - All of these used the base-case values for the laboratory-based test and the POCT<br>1. Test on admission to the hospital with SARS-CoV-2 laboratory-based tests with a time to test result of 35.9 hours, and routine retesting following admission to hospital in a non-SARS-CoV-2 infection-suspected bay with SARS-CoV-2 laboratory-based tests with a time to test result of 35.9 hours<br>2. Test on admission to the hospital with SARS-CoV-2 laboratory-based tests with a time to test<br>result of 35.9 hours, retest those with clinical symptoms suggestive of COVID-19 but with a negative SARS-CoV-2 test with SARS-CoV-2 laboratory-based tests with a time to test result of 35.9 hours, and routine retesting following admission to hospital in a non-SARS-CoV-2 infection-suspected bay with SARS-CoV-2 laboratory-based tests with a time to test result of 35.9 hours<br>3. Test on admission to the hospital with SARS-CoV-2 POCTs with a time to test result of 30 minutes, retest those with clinical symptoms suggestive of COVID-19 but with a negative SARS-CoV-2 test with SARS-CoV-2 POCTs and with a time to test result of 30 minutes, and routine retesting following admission to hospital in a non-SARS-CoV-2 infection-suspected bay test with SARS-CoV-2 POCTs with a time to test result of 30 minutes<br>4. Test on admission to the hospital with SARS-CoV-2 POCTs with a time to test result of 30 minutes, retest those with clinical symptoms suggestive of COVID-19 but with a negative SARS-CoV-2 test with SARS-CoV-2 POCTs and with a time to test result of 30 minutes, and routine retesting following admission to hospital in a non-SARS-CoV-2 infection-suspected bay test with SARS-CoV-2 laboratory-based tests with a time to test result of 35.9 hours<br>5. Test on admission to the hospital with SARS-CoV-2 laboratory-based tests with a time to test result of 35.9 hours, retest those with clinical symptoms suggestive of COVID-19 but with a negative SARS-CoV-2 test with SARS-CoV-2 laboratory-based tests with a time to test result of 35.9 hours, and routine retesting following admission to hospital in a non-SARS-CoV-2 infection-suspected bay with SARS-CoV-2 POCTs with a time to test result of 30 minutes<br>6. Test on admission to the hospital with SARS-CoV-2 laboratory-based tests with a time to test result of 35.9 hours, routine retesting following admission to hospital in a non-SARS-CoV-2 infection-suspected bay with SARS-CoV-2 laboratory-based tests with a time to test result of 35.9 hours, and weekly asymptomatic staff testing with SARS-CoV-2 laboratory-based tests with a time to test result of 35.9 hours<br>7. Test on admission to the hospital with SARS-CoV-2 laboratory-based tests with a time to test result of 35.9 hours, retest those with clinical symptoms suggestive of COVID-19 but with a negative SARS-CoV-2 test with SARS-CoV-2 laboratory-based tests with a time to test result of 35.9 hours, routine retesting following admission to hospital in a non-SARS-CoV-2 infection-suspected bay with SARS-CoV-2 laboratory-based tests with a time to test result of 35.9 hours, and weekly asymptomatic staff testing with SARS-CoV-2 laboratory-based tests with a time to test result of 35.9 hours<br>8. Test on admission to the hospital with SARS-CoV-2 POCTs with a time to test result of 30 minutes, retest those with clinical symptoms suggestive of COVID-19 but with a negative SARS-CoV-2 test with SARS-CoV-2 POCTs with a time to test result of 30 minutes, routine retesting following admission to hospital in a non-SARS-CoV-2 infection-suspected bay with SARS-CoV-2 POCTs with a time to test result of 30 minutes, and weekly asymptomatic staff testing with SARS-CoV-2 POCTs with a time to test result of 30 minutes<br>9. Test on admission to the hospital with SARS-CoV-2 POCTs with a time to test result of 30 minutes, retest those with clinical symptoms suggestive of COVID-19 but with a negative |
|  |  |  |  |  | SARS-CoV-2 test with SARS-CoV-2 POCTs with a time to test result of 30 minutes, routine retesting following admission to hospital in a non-SARS-CoV-2 infection-suspected bay with SARS-CoV-2 laboratory-based tests with a time to test result of 35.9 hours, and weekly asymptomatic staff testing with SARS-CoV-2 laboratory-based tests with a time to test result of 35.9 hours<br>10. Test on admission to the hospital with SARS-CoV-2 laboratory-based tests with a time to test result of 35.9 hours, retest those with clinical symptoms suggestive of COVID-19 but with a negative SARS-CoV-2 test with SARS-CoV-2 laboratory-based tests with a time to test result of 35.9 hours, routine retesting following admission to hospital in a non-SARS-CoV-2 infection-suspected bay with SARS-CoV-2 POCTs with a time to test result of 30 minutes, and weekly asymptomatic staff testing with SARS-CoV-2 POCTs with a time to test result of 30 minutes<br>11. As strategy 2 but using a time to test result of 24 hours for the laboratory-based test<br>12. As strategy 2 but using a time to test result of 6 hours for the laboratory-based test<br>13. As strategy 3 but using the acceptable TPP for POCTs<br>14. As strategy 3 but using data from currently available POCTs<br>15. As strategy 14 but using a time to test result of 8 hours<br>16. As strategy 4 but using the acceptable TPP for POCTs<br>17. As strategy 4 but using data from currently available POCTs,<br>18. As strategy 5 but using a time to test result of 24 hours for the laboratory-based test<br>19. As strategy 5 but using a time to test result of 6 hours for the laboratory-based test<br>20. As strategy 6 but using a time to test result of 24 hours for the laboratory-based test<br>21. As strategy 6 but using a time to test result of 6 hours for the laboratory-based test<br>22. As strategy 7 but using a time to test result of 24 hours for the laboratory-based test<br>23. As strategy 7 but using a time to test result of 6 hours for the laboratory-based test<br>24. As strategy 8 but using the acceptable TPP for POCTs<br>25. As strategy 8 but using data from currently available POCTs<br>26. As strategy 25 but using a time to test result of 8 hours<br>27. As strategy 9 but using the acceptable TPP for POCTs<br>28. As strategy 9 but using data from currently available POCTs<br>29. As strategy 10 but using a time to test result of 24 hours for the laboratory-based test<br>30. As strategy 10 but using a time to test result of 6 hours for the laboratory-based test |
| Shlomai et al, 2020 (34) | No funding | Israel Simulated population of Israel (9 million population) | Modeling and simulation | Societal | 1. Isolation – complete isolation of infected individuals or individuals at high exposure risk in a dedicated facility, 2. National lockdown of the susceptible population - national lockdown, isolation of all infected individuals, and a 14-day isolation period for the high exposure risk group |
| Keogh-Brown et al, | Yes | UK population | Modeling and simulation | Societal | 1. No action is taken to mitigate the virus spread VS<br>2. Mitigation policy, |
| 2020 (33) |  | based on UN population projections |  |  | 3. Suppression policy |
| Águas et al, 2021 (49) | Yes | UK 300 000 Hospitalised COVID-19 patients | Modeling and simulation | Healthcare system | 1. Dexamethasone treatment VS<br>2. No Dexamethasone treatment |
| Miles et al, 2020 (32) | Not clearly specified | UK Over 5 million citizens | Modeling | Societal | Lockdown policy |
| Shaker et al, 2021 (44) | Yes | USA Individual and 300 000 000 simulated persons | Simulation | 1) Healthcare and 2) Societal | 1. Universal COVID-19 vaccination VS<br>2. Risk-stratified vaccination<sup>2</sup> |
| Miles et al, 2021 (31) | Not clearly specified | UK 56 000 000 citizens | Modeling and simulation | Not clearly specified, however, societal could be ideal to mention | Physical restriction scenarios:<br>1. Base—no transition (0 wk High then 0 wk Medium then 26 wk Low),<br>2. I - Short transition (8 wk Medium then 18 wk Low),<br>3. II - Long transition (4 wk High then 12 wk stepped reduction - Medium),<br>4. III -Keep at high for 26 wk |
| Zala et al, 2020 (30) | No funding | UK UK population | Simulation | Societal | Suppression policies: 1) Suppression 1 (triggered “on” when there are 100 ICU cases in a week and “off” when weekly cases halve to 50 cases) VS unmitigated, 2) Suppression 1 (triggered “on” when there are 100 ICU cases in a week and “off” when weekly cases halve to 50 cases) VS mitigated, 3) Suppression 2 (triggered “on” when there are 400 ICU cases in a week and “off” when weekly cases halve to 200 cases) VS unmitigated, 4) Suppression 2 (triggered “on” when there are 400 ICU cases in a week and “off” when weekly cases halve to 200 cases) VS mitigated |
| Sheinson et al, 2021 (50) | Yes | USA Modeled population of hospitalized patients with COVID-19 | Modeling | 1) Healthcare, 2) Societal, 3) Payer | Treatment in the hospital with COVID-19 treatments VS Best supportive care (BSC) |
| Neilan et al, 2020 (38) | Yes | USA<br>Initial distribution of COVID-19 disease severity by age which has been derived from the Massachusetts Census and Department of Public Health | Modeling and simulation | Healthcare system | Testing strategies:<br>1) hospitalized: PCR testing only for patients with severe/critical symptoms warranting hospitalization,<br>2) symptomatic: PCR for any COVID-19-consistent symptoms, with self-isolation if positive,<br>3) symptomatic + asymptomatic once: symptomatic and 1-time PCR for the entire population,<br>4) symptomatic + asymptomatic monthly: symptomatic with monthly retesting for the entire population |
| Baggett et al, 2020 (40) | Yes | USA<br>Modeled cohort of 2258 adults residing in homeless shelters in Boston, Massachusetts | Modeling and simulation | Healthcare system | 1) No intervention VS<br>2) Symptom screening, PCR, and hospital,<br>3) Symptom screening, PCR, and ACS,<br>4) Universal PCR testing and hospital,<br>5) Universal PCR and ACS,<br>6) Universal PCR and temporary housing,<br>7) Hybrid hospital (symptom screening, PCR, and hospital strategy and adds shelter-based universal PCR testing every 2 weeks for those without symptoms),<br>8) Hybrid ACS ( symptom screening, PCR, and ACS strategy and adds shelter-based universal PCR testing every 2 weeks for those without symptoms) |
| Currie et al, 2020 (37) | No funding | Wales<br>192 suspected COVID-19 cases in community and standard hospital testing practices | Retrospective cohort study | Healthcare system | 1) Community testing by swabbing VS<br>2) Hospital standard testing |
| Sandmann et al, 2021 (53) | Yes | UK<br>UK population | Modeling and simulation | Healthcare system and society | Vaccination alongside physical distancing measures<br>1) No vaccination VS<br>2) Vaccination (worst case),<br>3) Vaccination (best case) |
| Barnett-Howell et al, 2021 (29) | Yes | High and low income countries (including the UK, US)<br>Susceptible modeled population at the start of the pandemic | Simulation | Not clearly specified | Physical distancing strategies:<br>1) Unmitigated spread of COVID-19, where the virus progresses through each country at a high level of infectivity,<br>2) Individual distancing policy, similar to the case of a country like Sweden where workplaces, schools and restaurants mostly remain open but large gatherings are discouraged, reducing infectivity by 15%,<br>3) Broad physical distancing policy, equivalent to the closure of schools and workplaces, that reduces infectivity by 35%,<br>4) Physical distancing+, which is an even more intensive physical distancing policy with targeted stay-at-home orders,<br>5) Full lockdown policies designed to entirely suppress physical contact, closing transit and stay-at-home orders for the entire population, reducing infectivity by 60% |
| Gandjour et al, 2020 (28) | No funding | Germany<br>German population | Modeling | Societal | 1) No intervention, VS<br>2) Shutdown in scenario of "Flattening the curve",<br>3) Shutdown in scenario of "Squashing the curve",<br>4) ICU capacity exceeded by 50 %,<br>5) ICU capacity exceeded by 100 %,<br>6) ICU capacity exceeded by 200 %,<br>7) ICU capacity exceeded by 300 % |
| Gandjour et al, 2021 (55) | No funding | Germany<br>German population | Modeling | Societal | ICU bed provision |
| Bartsch et al, 2021 (45) | Yes | USA<br>U.S. population | Modeling and simulation | a) third-party payer, b) societal | 1) Vaccine that reduces severe disease,<br>2) Vaccine that prevents infection, 5% have already been infected,<br>3) Vaccine that prevents infection, 25% have already been infected,<br>4) Vaccine that prevents infection, 50% have already been infected |
| Thom et al, 2021 (27) | Yes | European countries (UK, Ireland, Germany, Spain and Sweden)<br>Population of these European countries | Modeling | Health system | 1) Scenario A (base case) - Government response mitigation under $R_0=2.7$,<br>2) Scenario B - Government response mitigation under $R_0=1.6$,<br>3) Scenario C - Government response mitigation under $R_0=3.9$,<br>4) Scenario: All GDP reduction due to response |
| Kohli et al, | Yes | USA | Modeling and | Healthcare | 1) No vaccine VS |
| 2021 (46) |  | Every adult (18 years and older) in the US | simulation | system | 2) COVID-19 vaccine |
| Chen et al, 2020 (25) | Yes | USA US population | Modeling and simulation | Healthcare system | 1) Stay-home for 0 days (unmitigated scenario) VS 2) Stay-home for 30 days, with VHI & SHI 60% compliance rate, 3) Stay-home for 30 days, with VHI & SHI 70% compliance rate, 4) Stay-home for 30 days, with VHI & SHI 80% compliance rate, 5) Stay-home for 30 days, with VHI & SHI 90% compliance rate, 6) Stay-home for 45 days, with VHI & SHI 60% compliance rate, 7) Stay-home for 45 days, with VHI & SHI 70% compliance rate, 8) Stay-home for 45 days, with VHI & SHI 80% compliance rate, 9) Stay-home for 45 days, with VHI & SHI 90% compliance rate, 10) Stay-home for 60 days, with VHI & SHI 60% compliance rate, 11) Stay-home for 60 days, with VHI & SHI 70% compliance rate, 12) Stay-home for 60 days, with VHI & SHI 80% compliance rate, 13) Stay-home for 60 days, with VHI & SHI 90% compliance rate |
| Losina et al, 2020 (52) | Yes | USA 105 000 cohort size | Simulation | Modified societal | No intervention VS Isolation intervention, masks wearing and laboratory testing/self-screen strategies<br>a) Residence-based isolation, incl. self-screen, b) Designated space for isolation, incl. self-screen, c) Designated spaces for isolation + 1-time laboratory testing, d) Designated spaces for isolation + routine laboratory testing every 14 days, e) Designated spaces for isolation + routine laboratory testing every 7 days, f) Designated spaces for isolation + routine laboratory testing every 3 days |
| Chen et al, 2020 (26) | Yes | USA US population | Simulation | Societal | 1) No non-pharmaceutical interventions applied VS 2) Stay-home for 30 days, with 60%-70%-80%-90% compliance rate, 3) Stay-home for 45 days, with 60%-70%-80%-90% compliance rate, 4) Stay-home for 60 days, with 60%-70%-80%-90% compliance rate |
| Vernaz et al, 2020 (47) | No funding | Switzerland 840 Hospitalized patients with a SARS-CoV-2 infection in COVID-19 wards | Retrospective cohort study | Healthcare system | 1) Standard of Care (SoC) VS 2) 400 mg of lopinavir/ritonavir twice daily for 5 days under the age of 75 years and 400 mg of lopinavir/ritonavir in the morning and 200 mg of lopinavir/ritonavir in the evening for patients over 75, 3) A single dose of 800 mg of hydroxychloroquine, 4) Lopinavir/ritonavir and hydroxychloroquine dual therapy at the same dosages. Lopinavir/ritonavir was first prescribed on 5 March 2020, and hydroxychloroquine alone or combined with lopinavir/ritonavir on 20 March 2020 |
| Ebigbo et al, 2021 (51) | Not mentioned/Not specified | Germany Simulation of 10 000 asymptomatic patients, 20 | Simulation | Societal | 1) No routine pre-endoscopy virus test; use of surgical masks, goggles, gloves, and apron for all procedures, 2) No routine pre-endoscopy virus test; additional use of FFP-2 masks and water-resistant gowns for all procedures, 3) Decentralized POC antigen test; use of surgical masks, goggles, gloves, and apron for all procedures, 4) Decentralized POC antigen test; additional use of FFP-2 and water-resistant gowns for all procedures irrespective of test result, 5) Centralized |
|  |  | full-time HCWs with two FFP-2 masks per day |  |  | laboratory-based rapid PCR test; use of surgical masks, goggles, gloves, and apron for all procedures, 6) Centralized laboratory-based rapid PCR test; additional use of FFP-2 and water-resistant gowns for all procedures irrespective of test result, 7) Centralized laboratory-based standard PCR test; use of surgical masks, goggles, gloves, and apron for all procedures, 8) Centralized laboratory-based standard PCR test; additional use of FFP-2 and water-resistant gowns for all procedures irrespective of test result |
| Paltiel et al, 2020 (41) | Yes | USA<br>4 990 students without SARS-CoV-2 infection and 10 students with undetected asymptomatic cases of SARS-CoV-2 | Modeling | Societal | 1) Symptom-based screening, with no test sensitivity, 2) Weekly screening, with 70% test sensitivity, 3) Weekly screening, with 80% test sensitivity, 4) Weekly screening, with 90% test sensitivity, 5) Every 3 days screening,, with 70% test sensitivity, 6) Every 3 days screening,, with 80% test sensitivity, 7) Every 3 days screening, with 90% test sensitivity, 8) Every 2 days screening, with 70% test sensitivity, 9) Every 2 days screening, with 80% test sensitivity, 10) Every 2 days screening, with 90% test sensitivity, 11) Daily screening, with 70% test sensitivity, 12) Daily screening, with 80% test sensitivity, 13) Daily screening, with 70% test sensitivity |
| Paltiel et al, 2021 (36) | Yes | USA<br>U.S. population | Modeling | Societal | 1) No home-based SARS-CoV-2 antigen testing VS<br>2) home-based SARS-CoV-2 antigen testing |

USA: United States of America, US: United States, PPE: Personal Protective Equipment, UK: United Kingdom, HWC: Home Healthcare Worker, VHI: Voluntary Home Isolation, SHI: Stay Home Isolation, GDP: Gross Domestic Product, PPE: Personal protective equipment, POCT: Point-of-care test, PCR: Polymerase Chain Reaction, ACS: Alternative Care Site, TPP: Target product profile, wk: weekLOS: Length of Stay, SARS-CoV-2: severe acute respiratory syndrome coronavirus 2, FFP-2: Filtering Facepiece 2
<sup>1</sup> Including countries in Europe
<sup>2</sup> based on history of anaphylaxis attributable to any trigger, where vaccination is deferred in patients with a history of self-reported anaphylaxis

Concerning study characteristics, thirteen out of 31 studies were from the USA (25, 26, 36, 38, 40, 41, 44–46, 48, 50, 52, 54), seven from the UK (30-33, 42, 49, 53), three from Germany (28, 51, 55), one from Switzerland (47), Israel (34), and Wales (37), while five of these studies included analyses for multiple OECD countries (27, 29, 35, 39, 43). The societal perspective was followed in 14 studies (26, 28, 30, 31, 33, 34, 36, 39, 41, 43, 51, 52, 54, 55), while 9 studies performed an analysis via a healthcare perspective (25, 27, 37, 38, 40, 42, 46, 47, 49, 53). Various perspectives were included in 4 studies (44, 45, 48, 50), including the payer for some of them (45, 48, 50), while the perspective considered was unclear in three studies (29, 32, 35).

#### Cost-effectiveness of testing & screening policies

Seven studies (**Table 4**) assessed the cost-effectiveness of testing and screening interventions. Stevenson et al. (2021) (42) compared 30 interventions and found that the least costly testing intervention was testing on hospital admission, and routine retesting following hospital admission. The authors noted that strategies with shorter times to test results were more cost-effective, all other things being equal, as were SARS-CoV-2 tests with greater diagnostic accuracy. trategies that included saliva sampling and nasopharyngeal have been assessed within the systematic review and meta-analysis by Bastos et al. (2021) (39). The incremental cost per additional SARS-CoV-2 infection detected with nasopharyngeal swabs versus saliva sampling was 6,427 € if the prevalence was 1%, and the cost-savings of saliva sampling were estimated at 505,177 € (95% CI 371,217-660,568) if 100 000 persons are tested. The clinical and economic value of PCR testing was investigated by Neilan et al. (2020) (38), who showed that under all epidemic growth scenarios considered, testing people with any COVID-19-consistent symptoms would be cost-saving compared to restricting testing to only those with symptoms severe enough to warrant hospital care, leading to lower costs and reducing infections and deaths. According to their results, and within the context of the first wave of the pandemic, the symptomatic and asymptomatic monthly testing scenario ws a cost-effective strategy only with an (R_e_) > 1.6 but would be no longer cost-effective option for lower reproduction scenarios, unless testing cost was lower. . A cost-minimization analysis performed by Currie et al. (2020) (37) comparing community testing undertaking swabbing of suspected cases in Wales with standard hospital testing showed that community testing for COVID-19 was a superior strategy, resulting in significant benefits for the healthcare system.

**Table 4.**
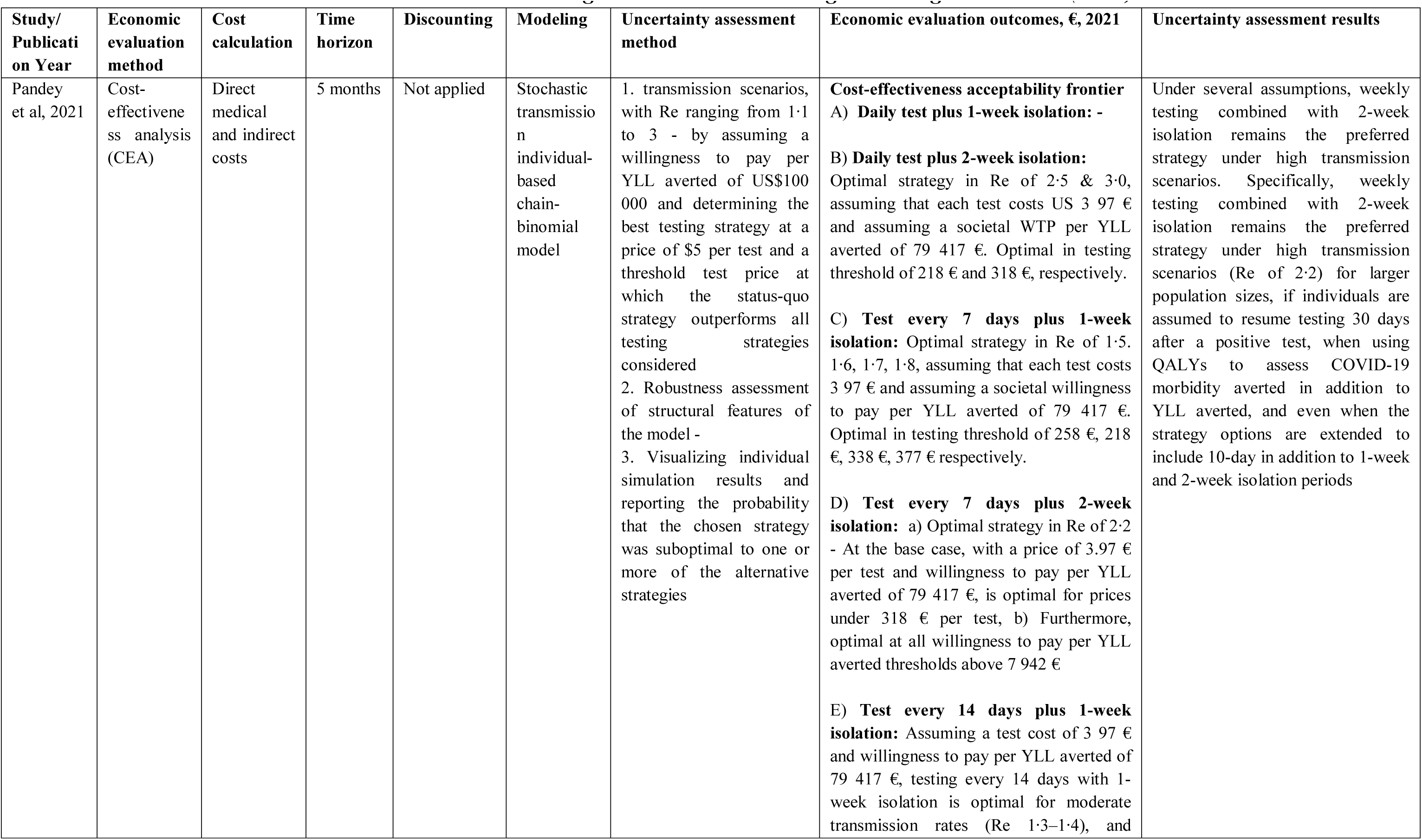
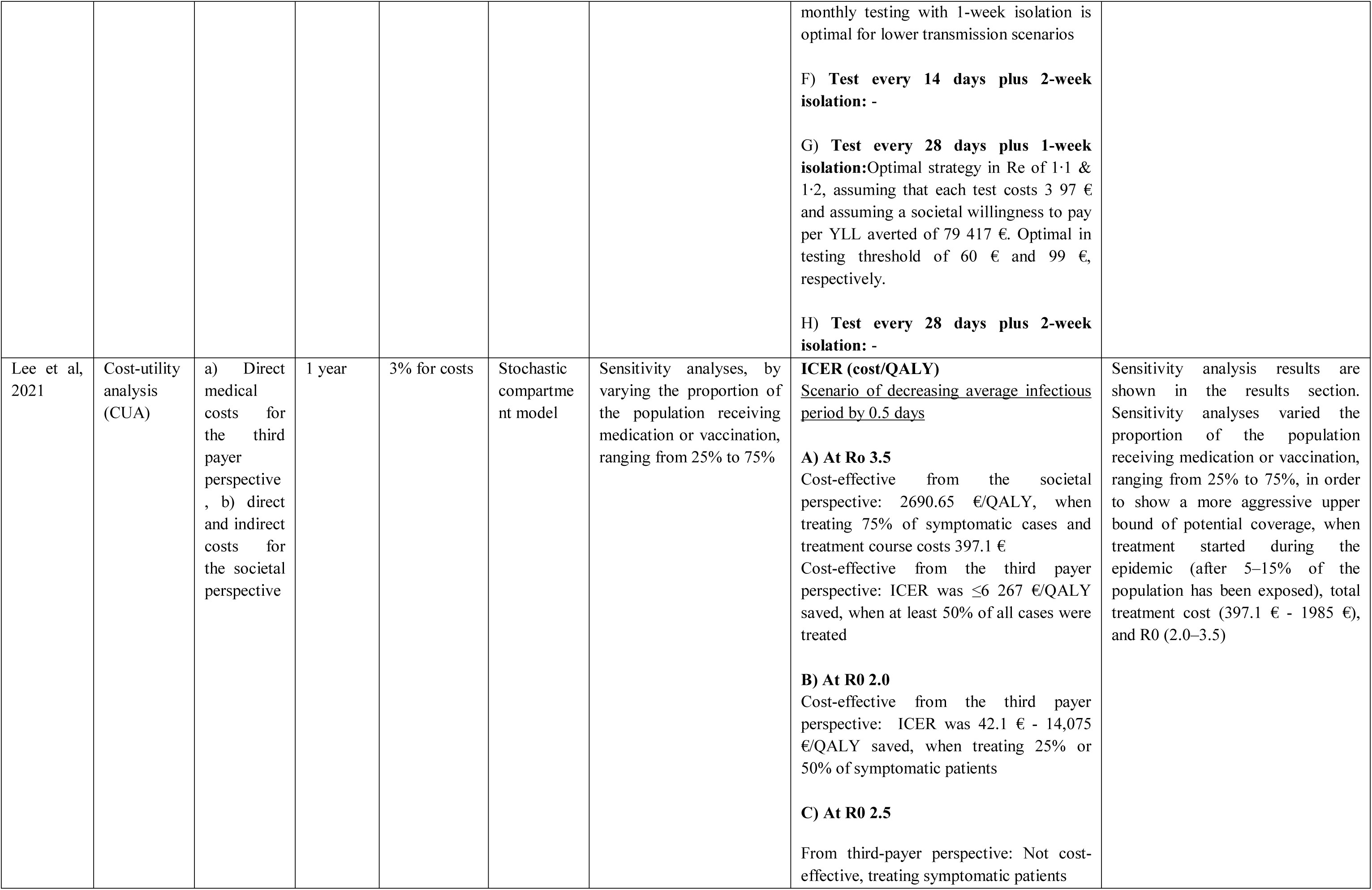
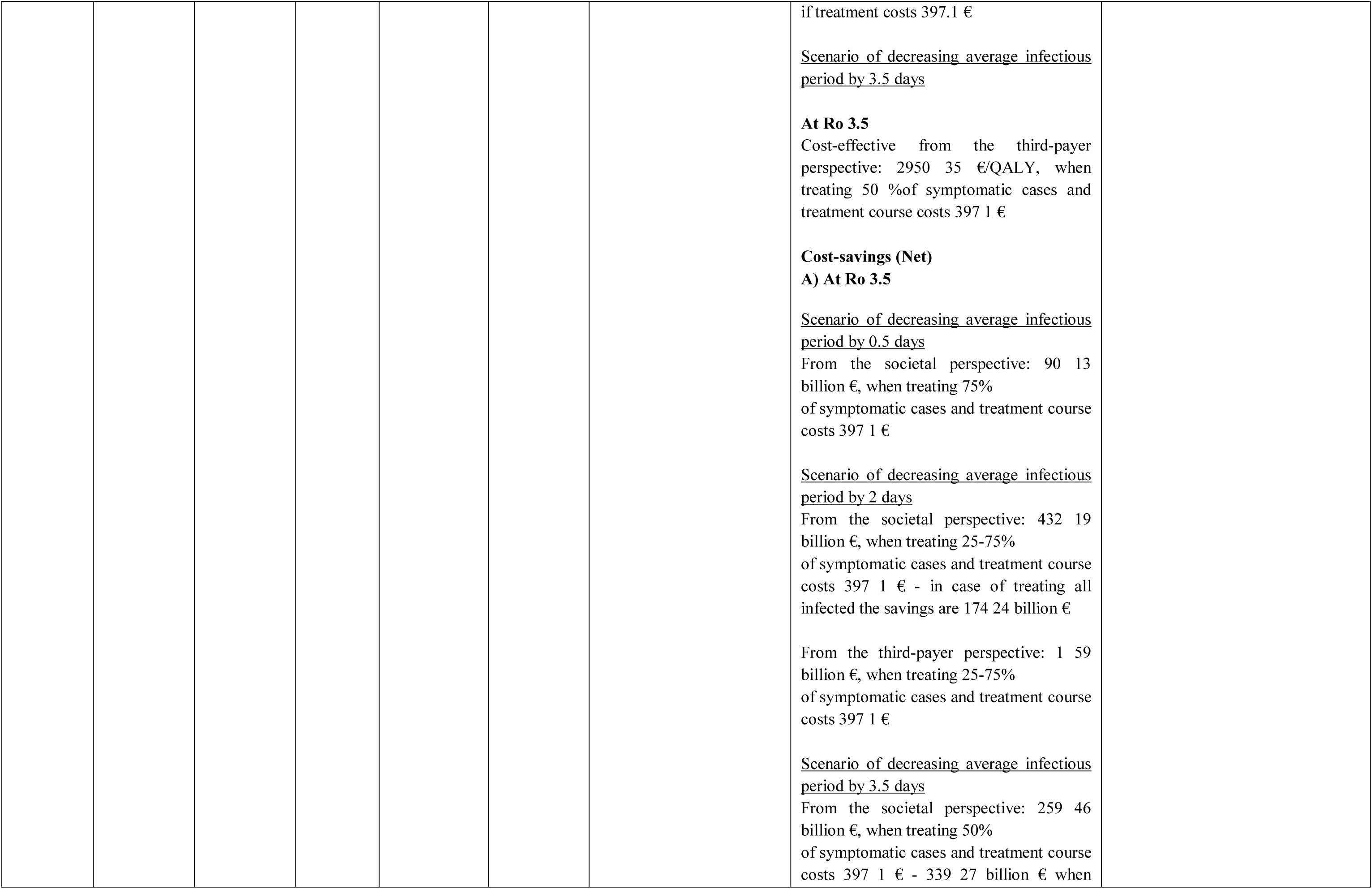
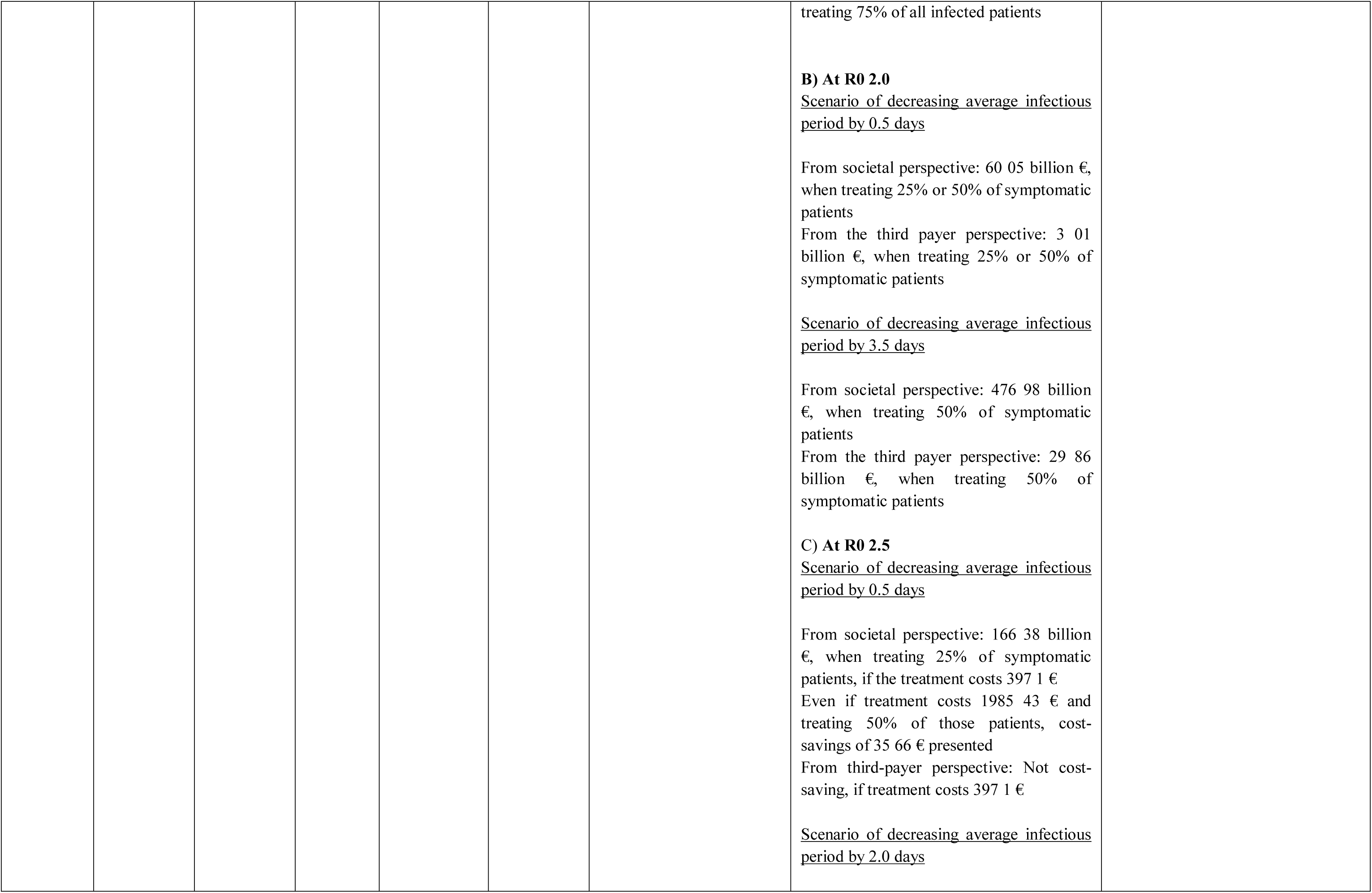
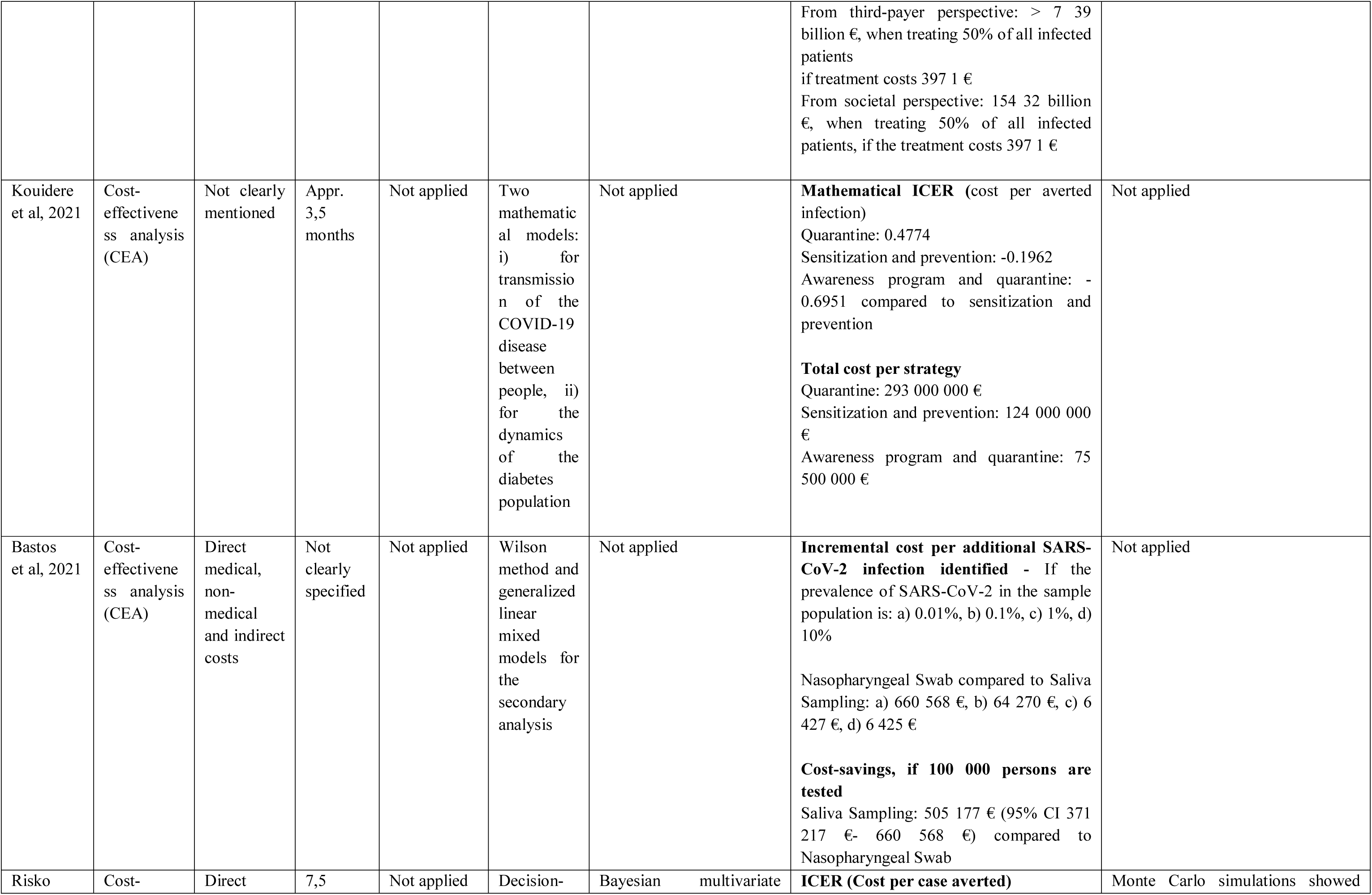
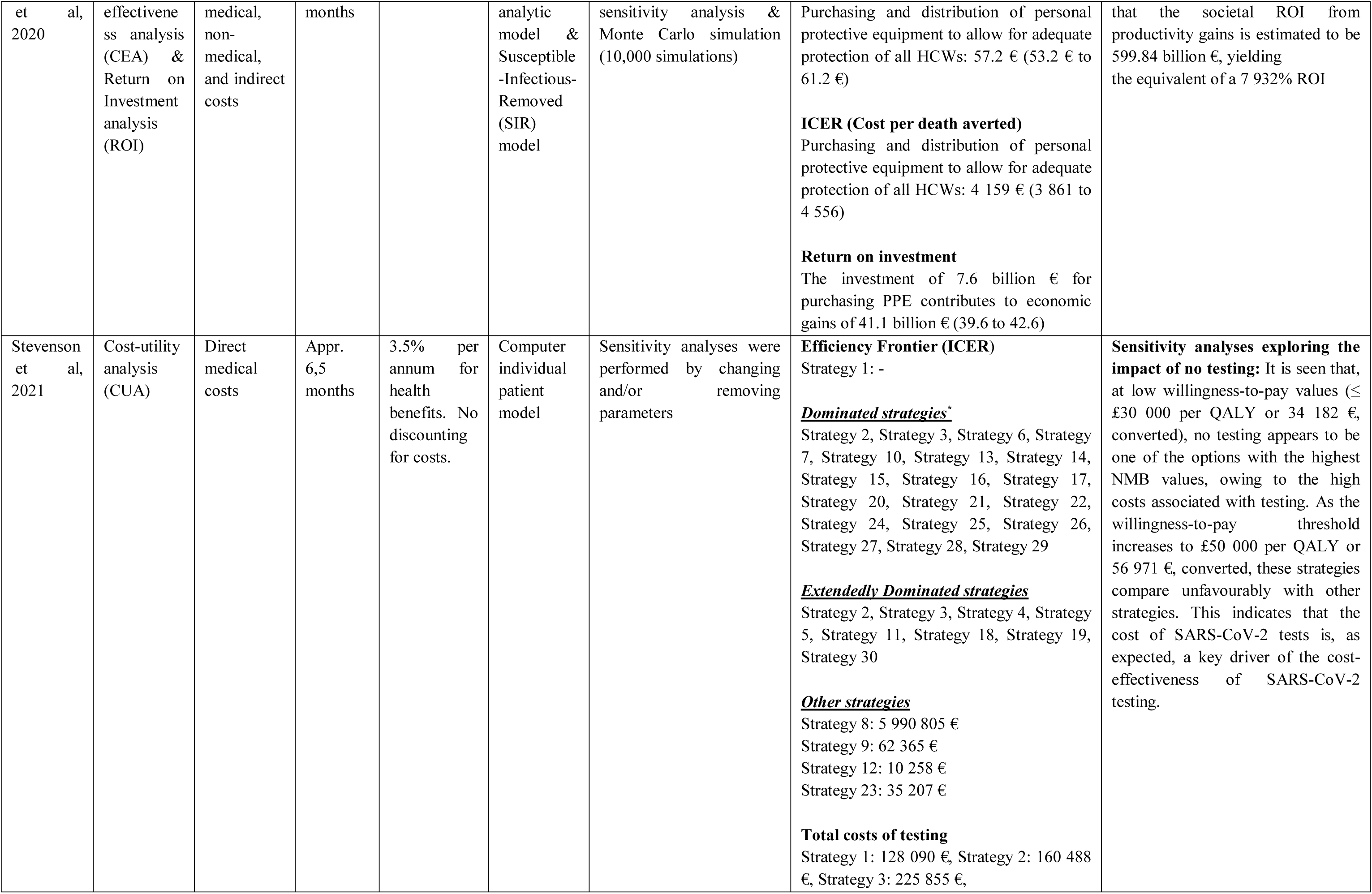
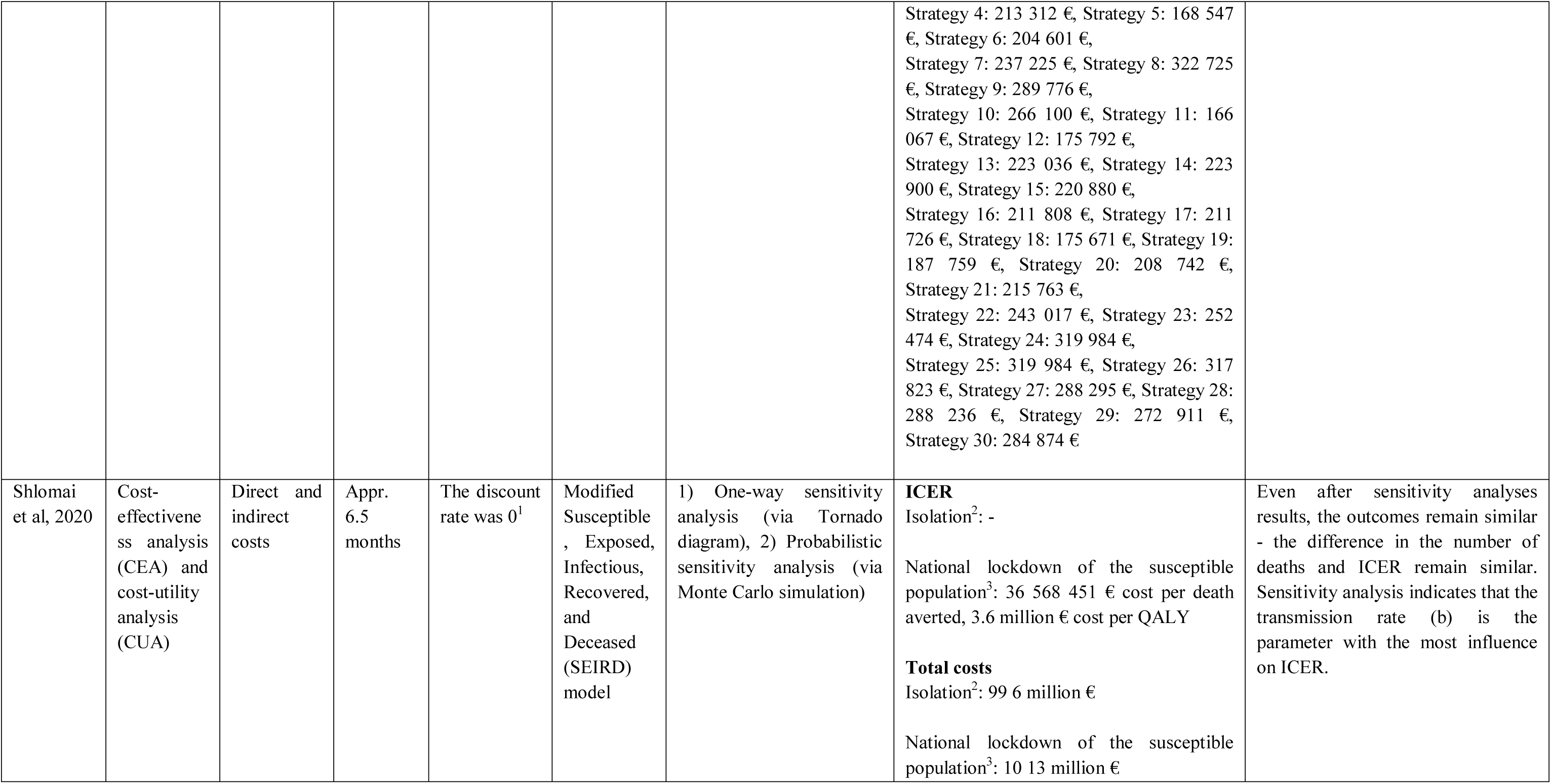
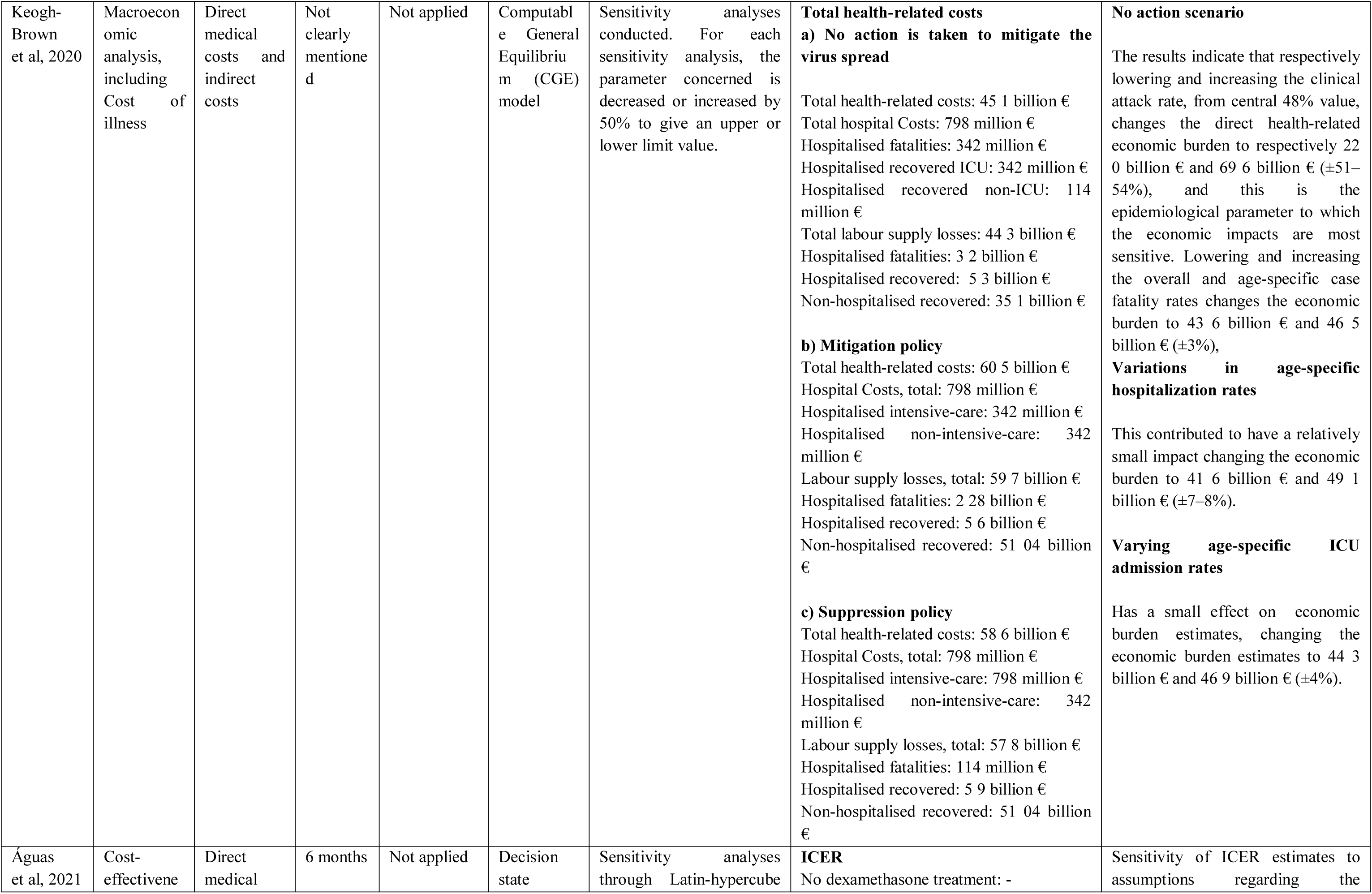
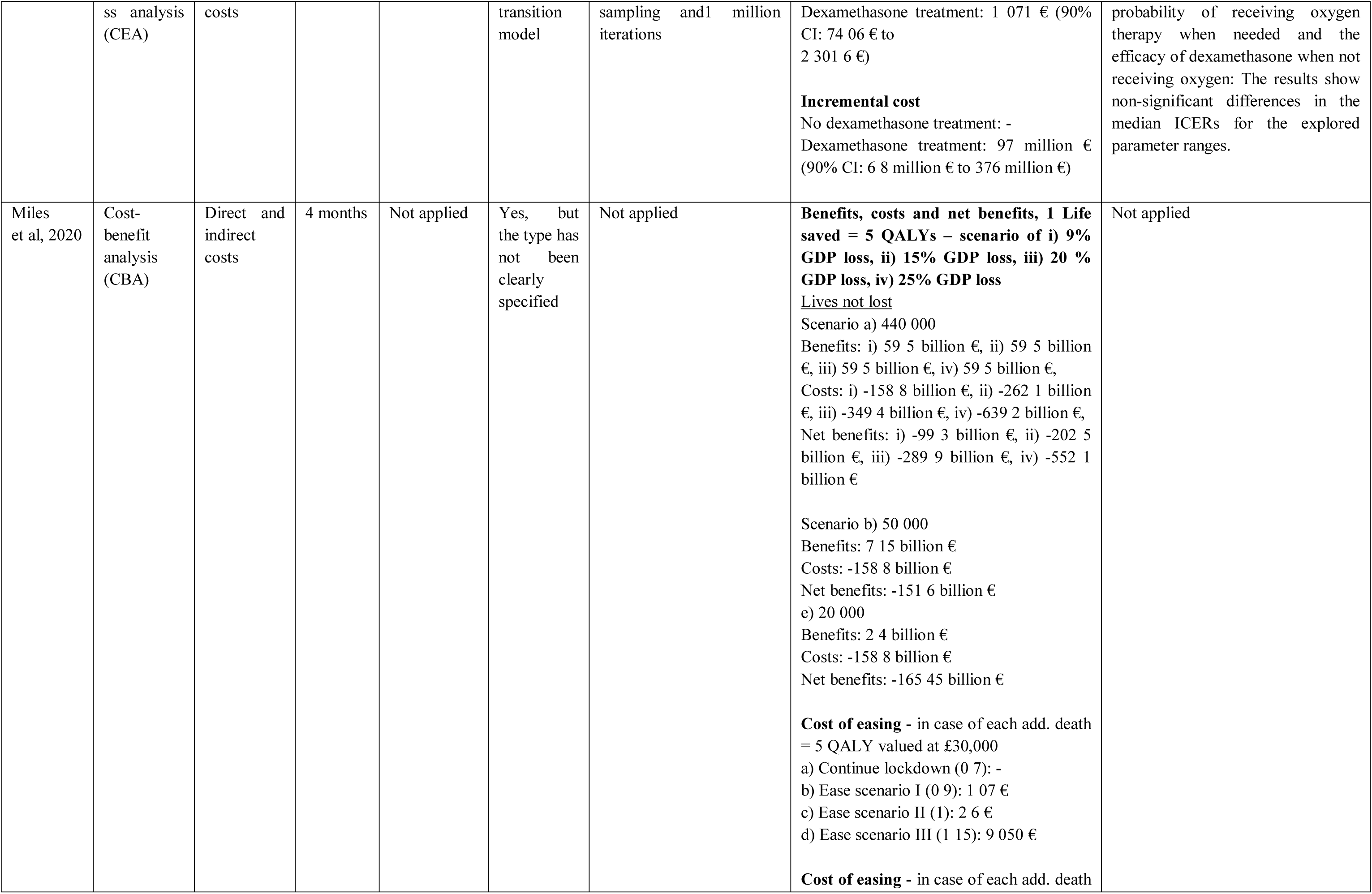
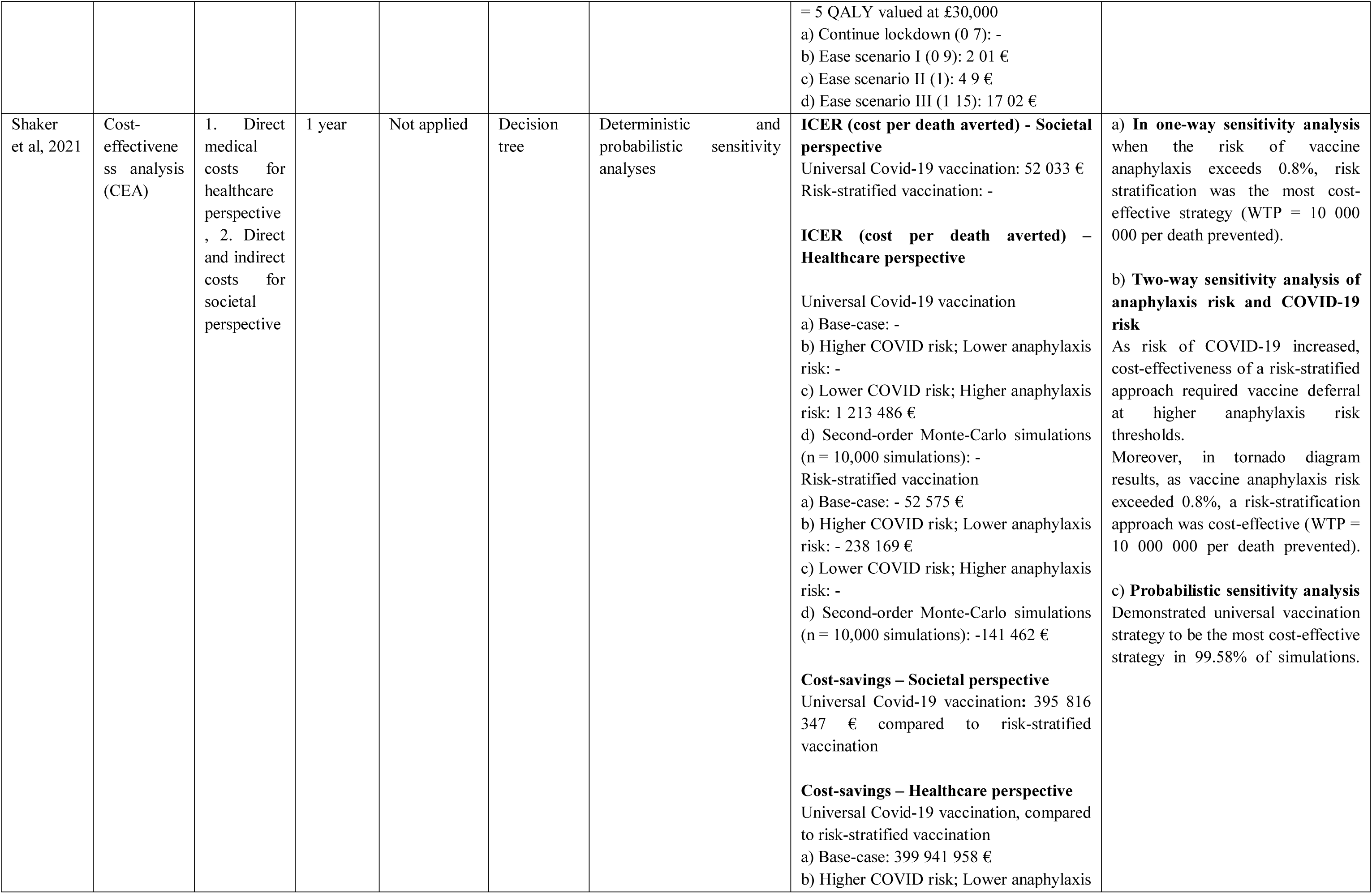
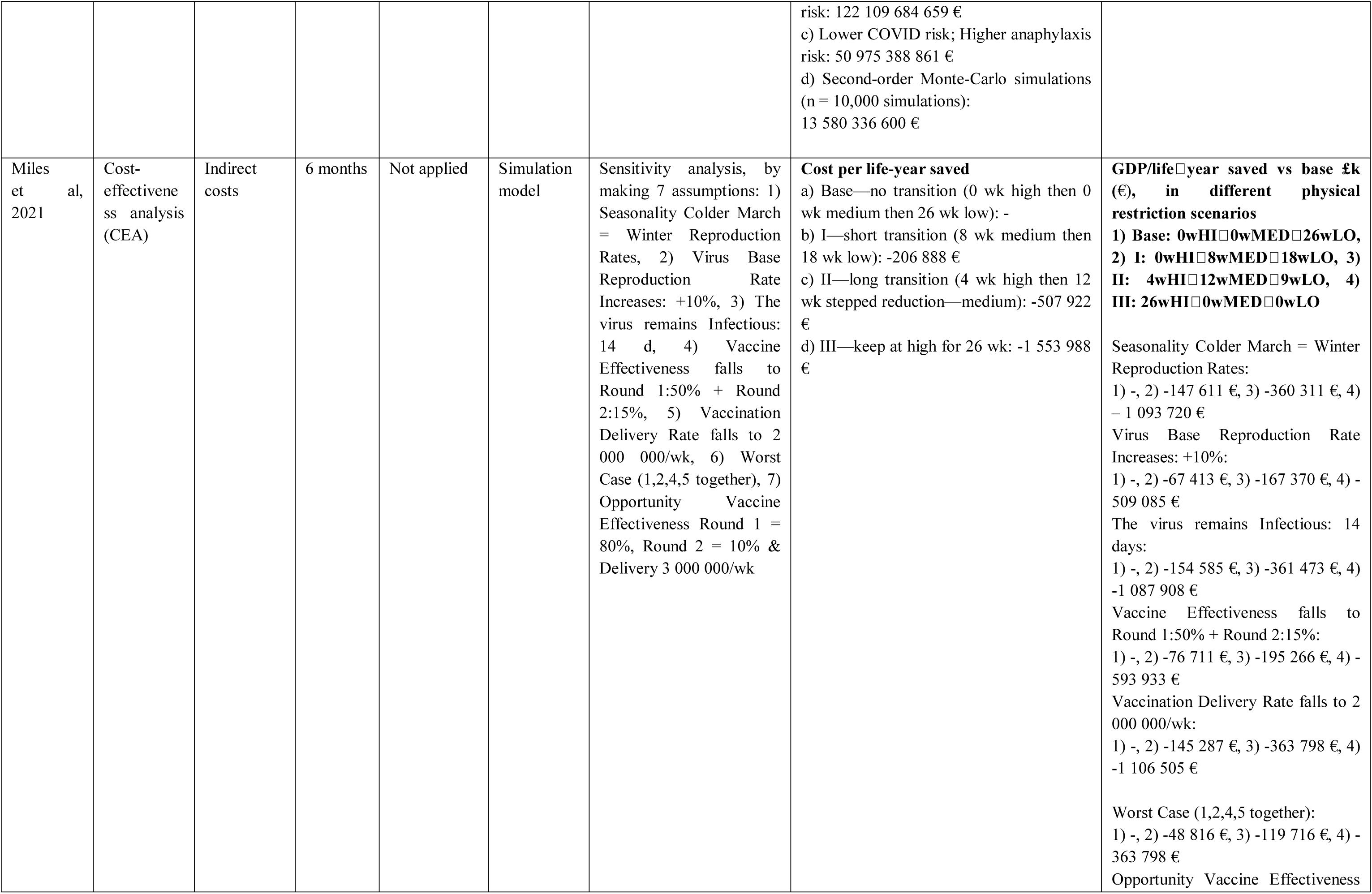
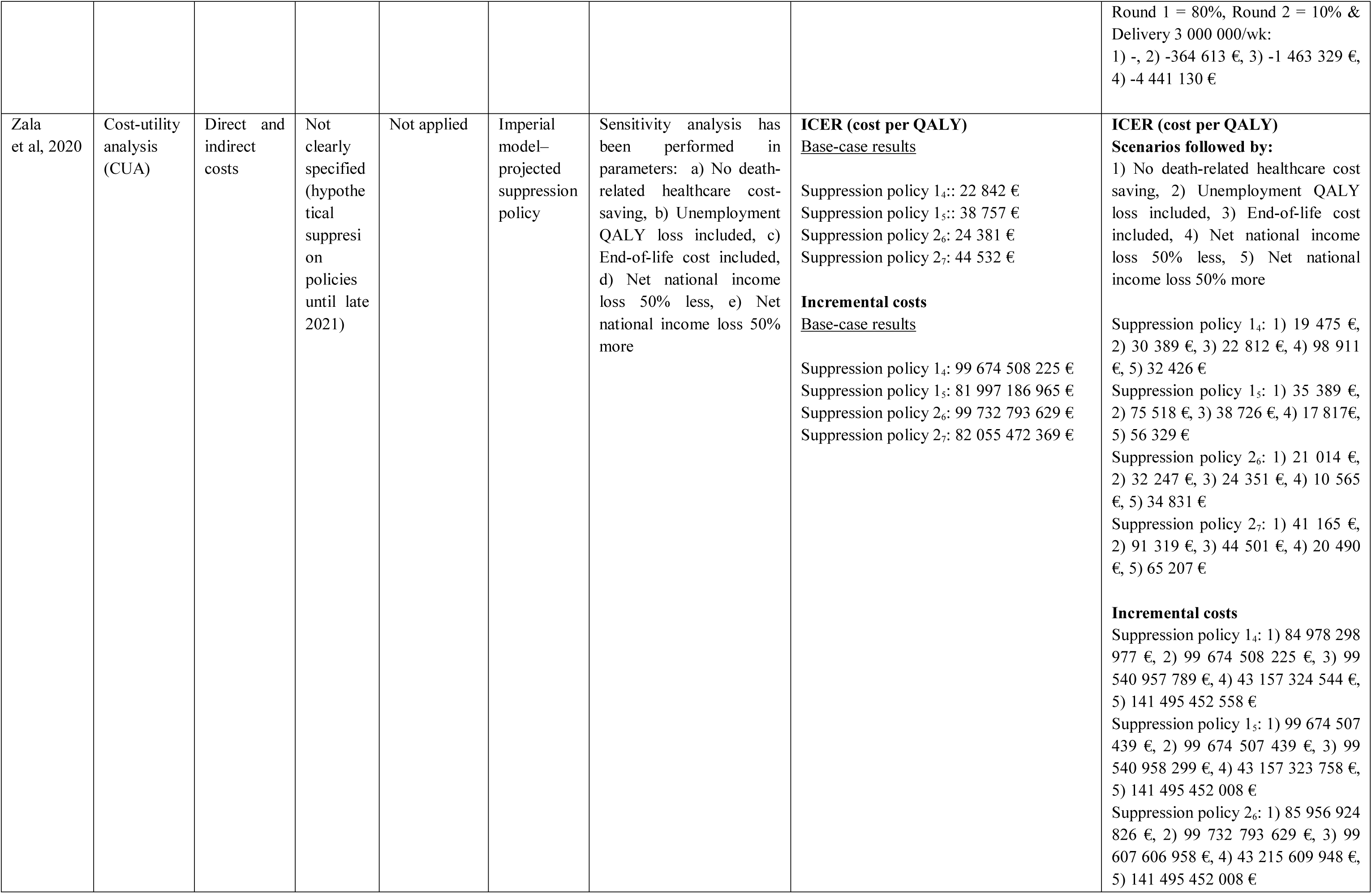
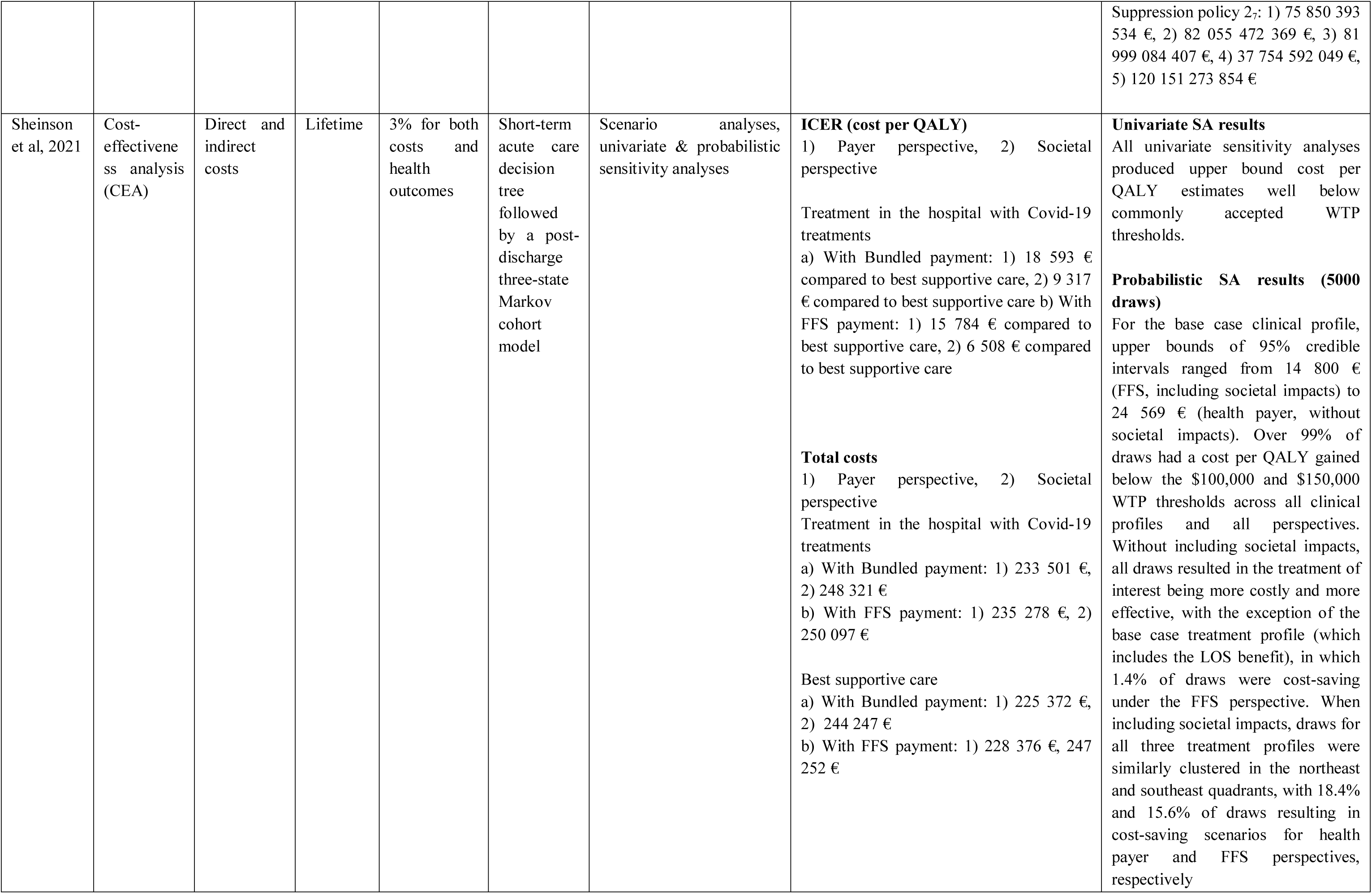
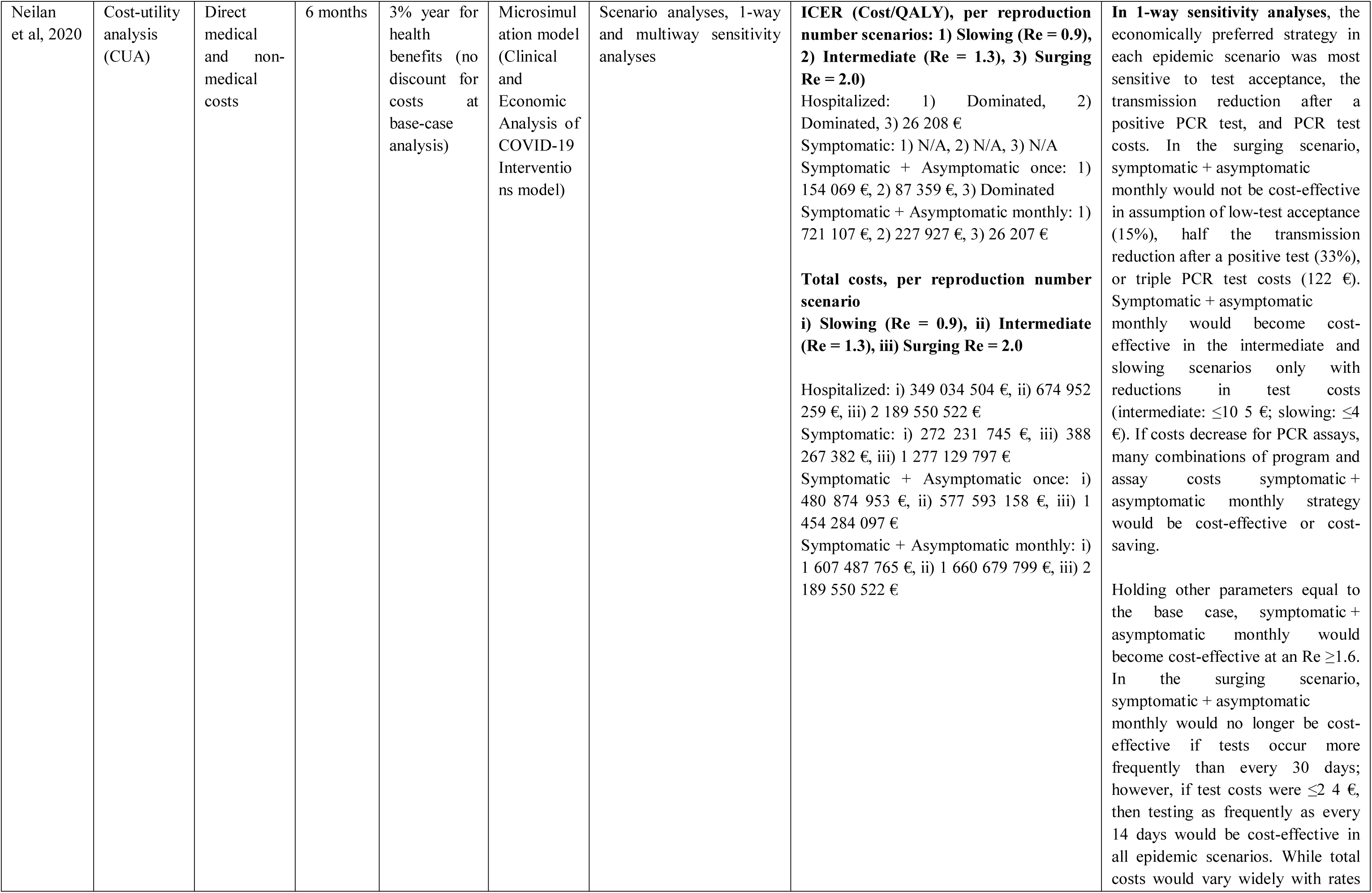
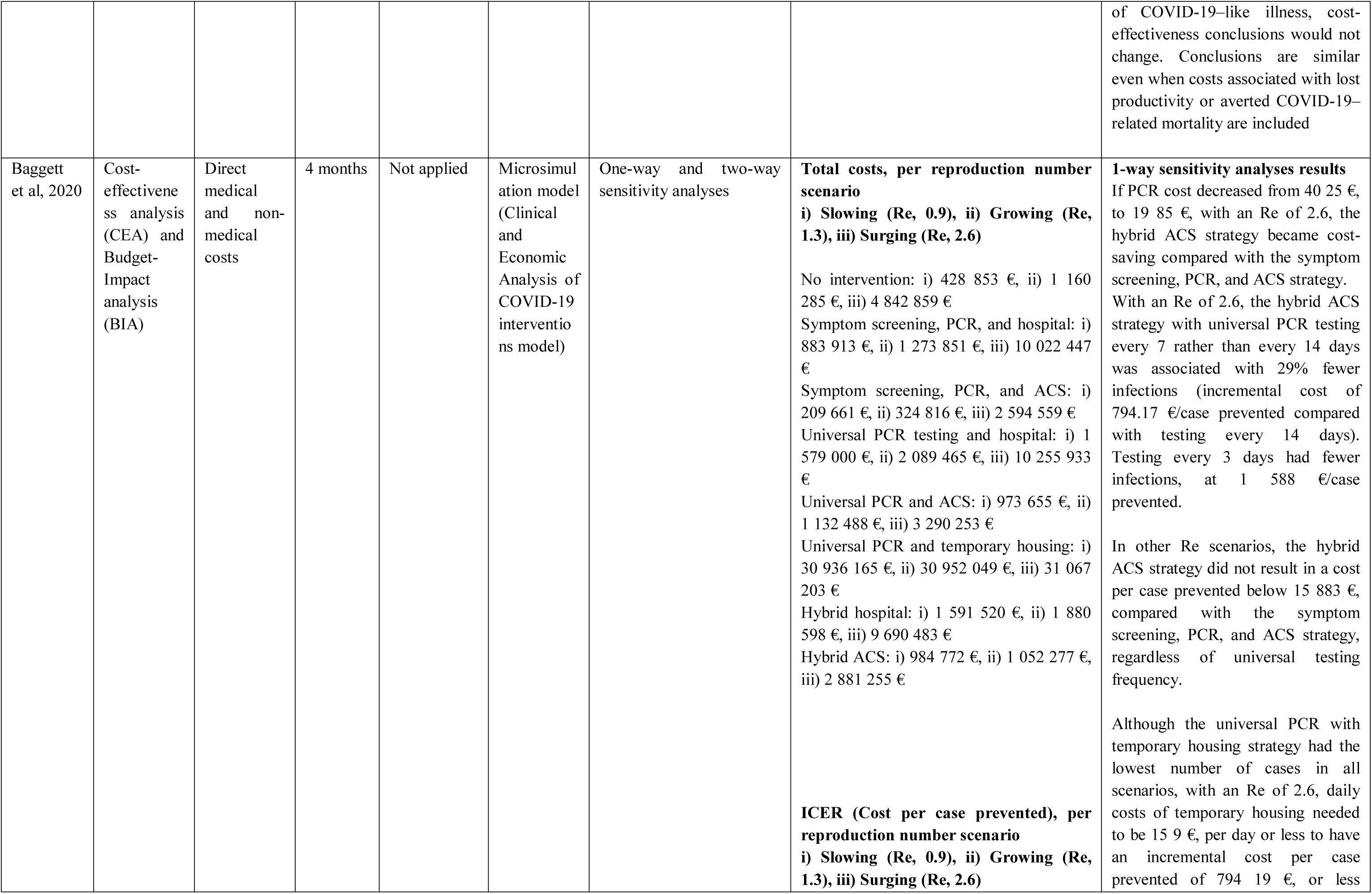
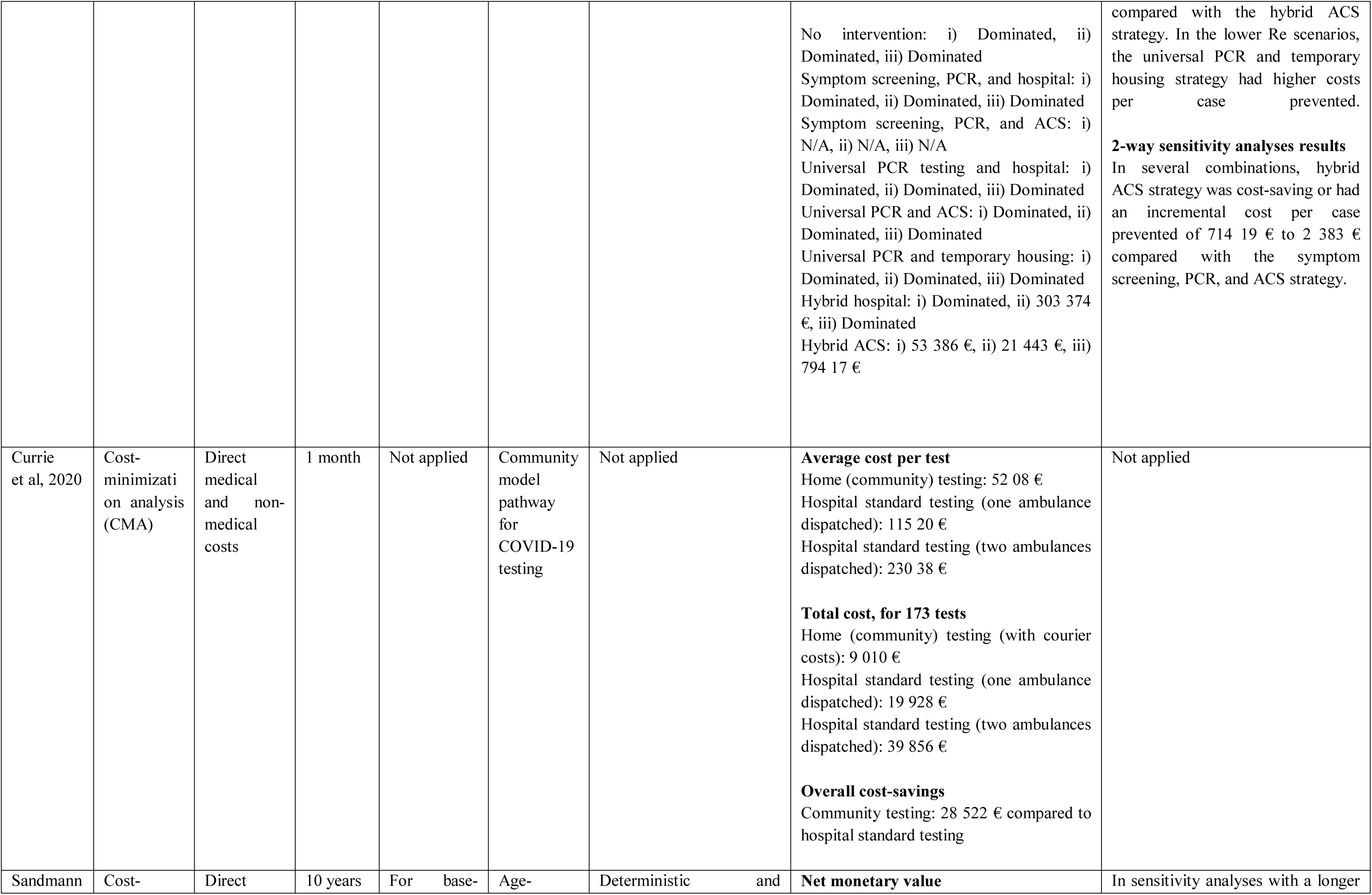
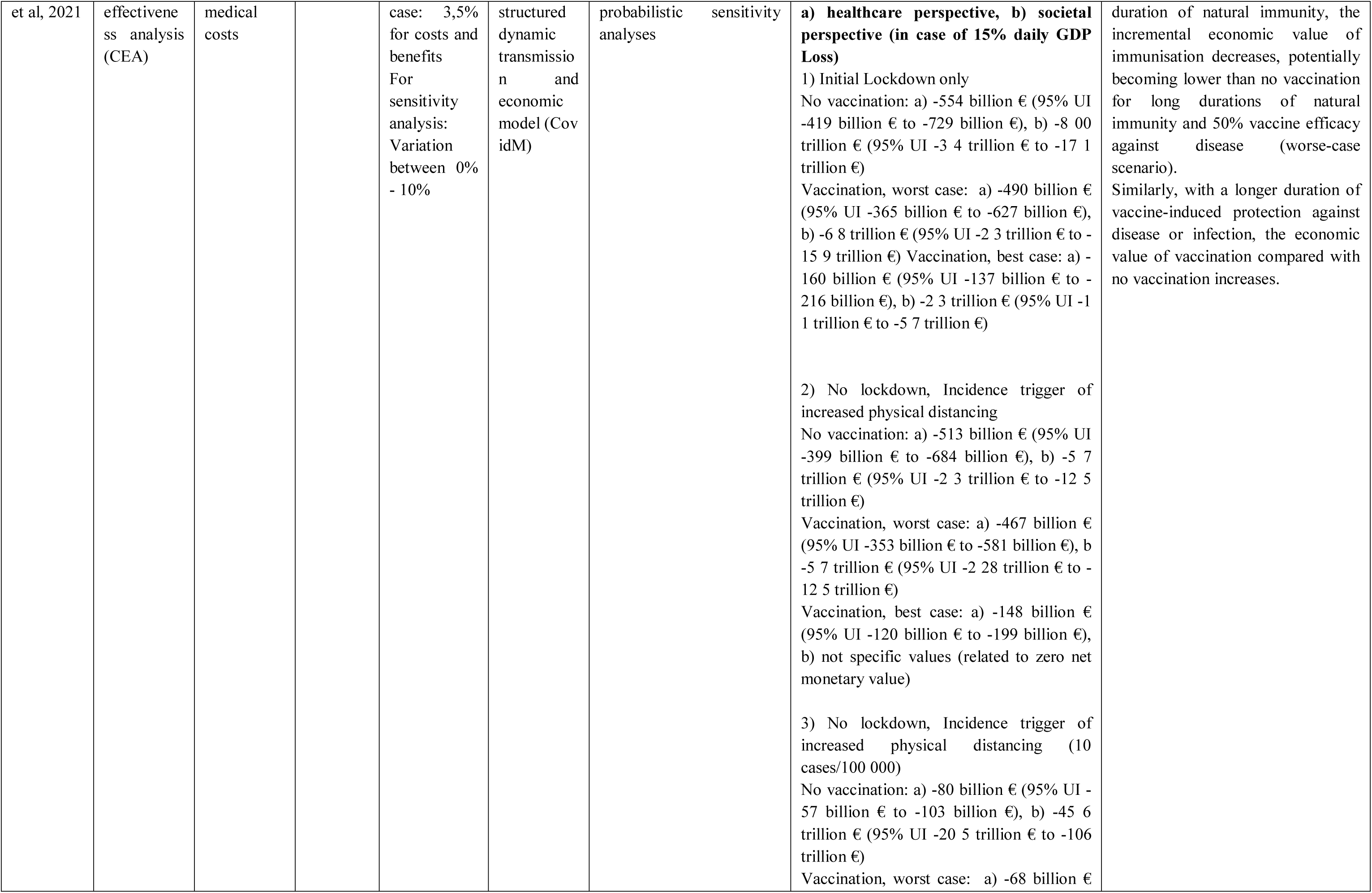
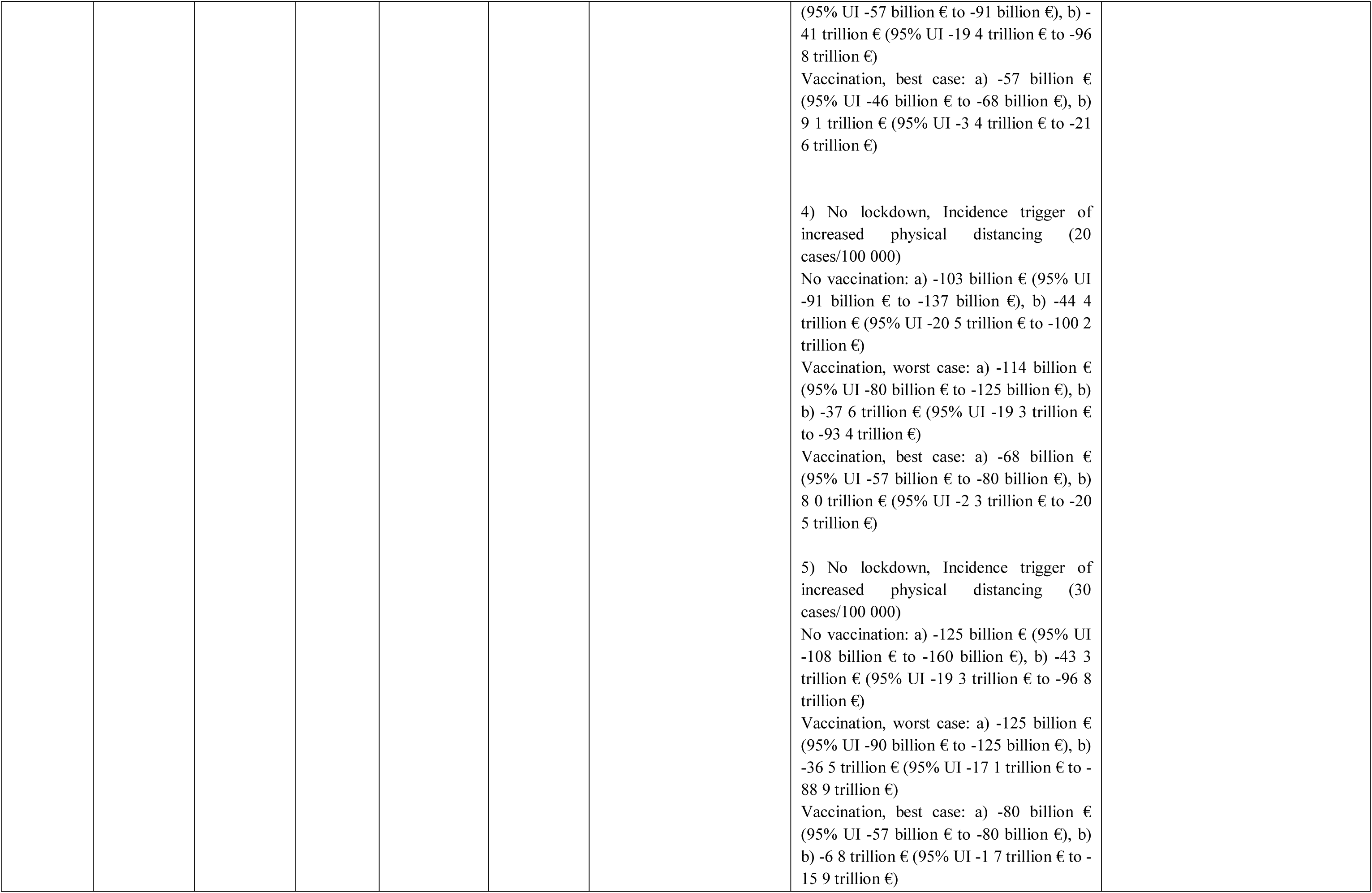
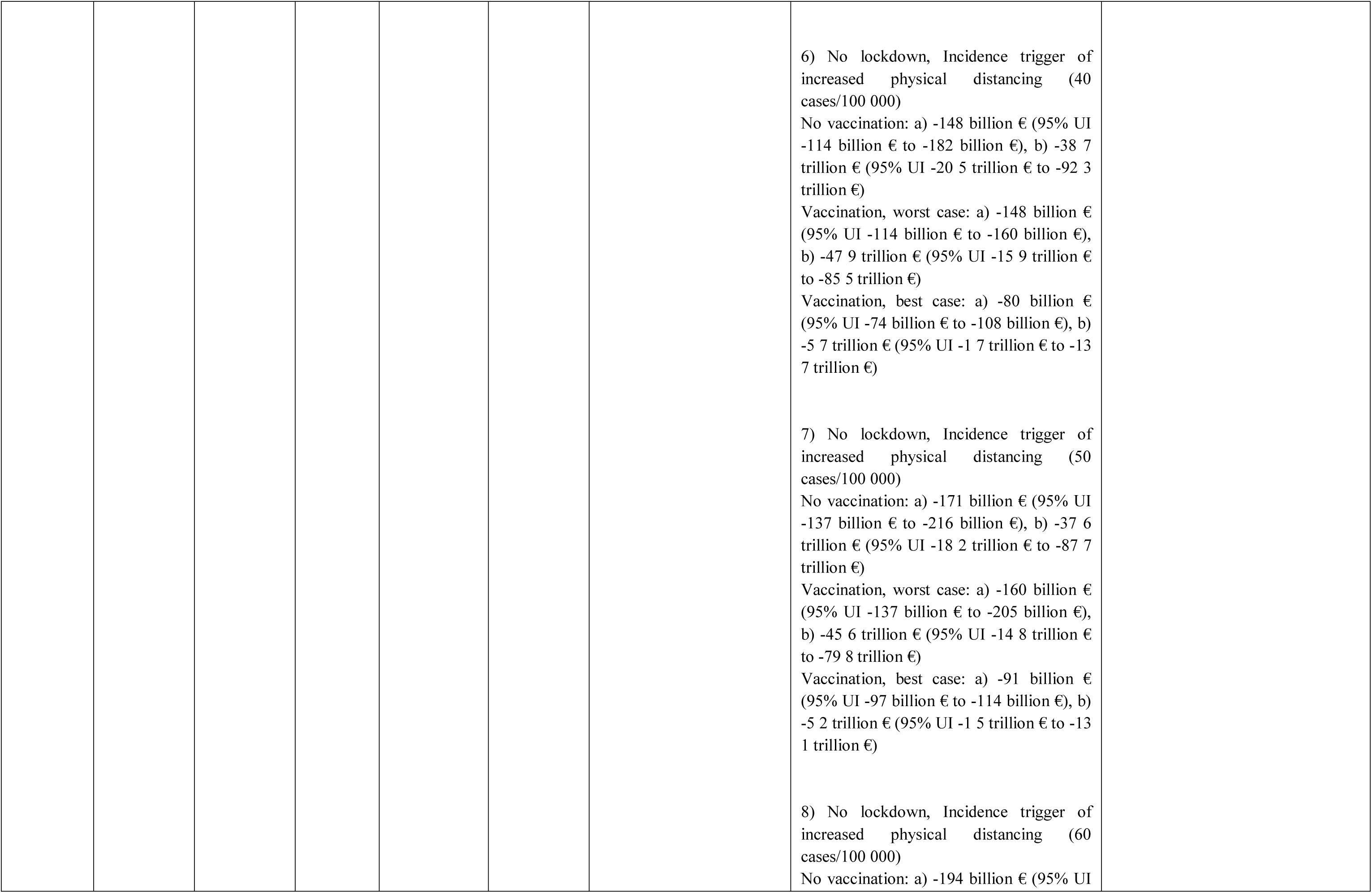
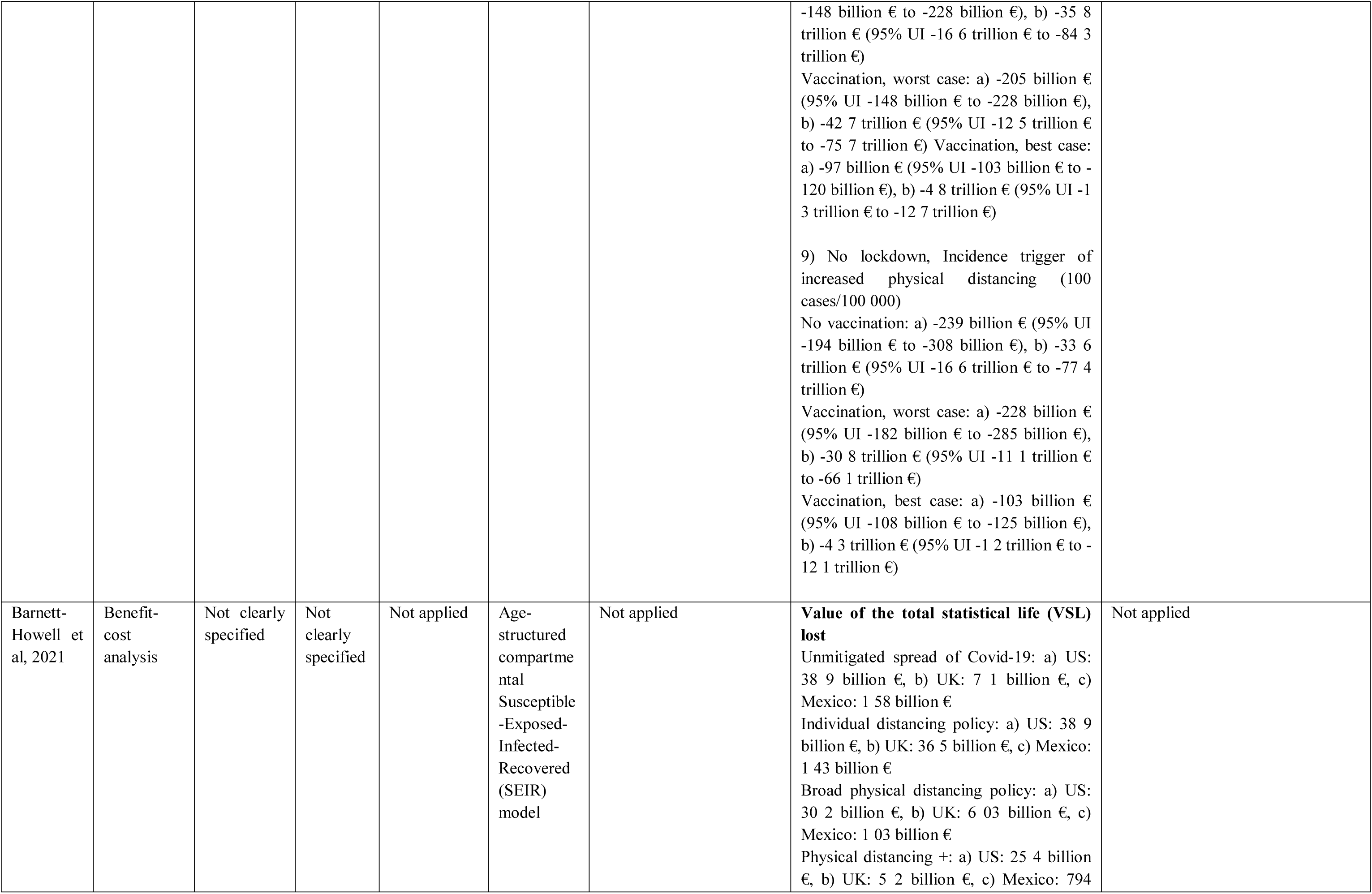
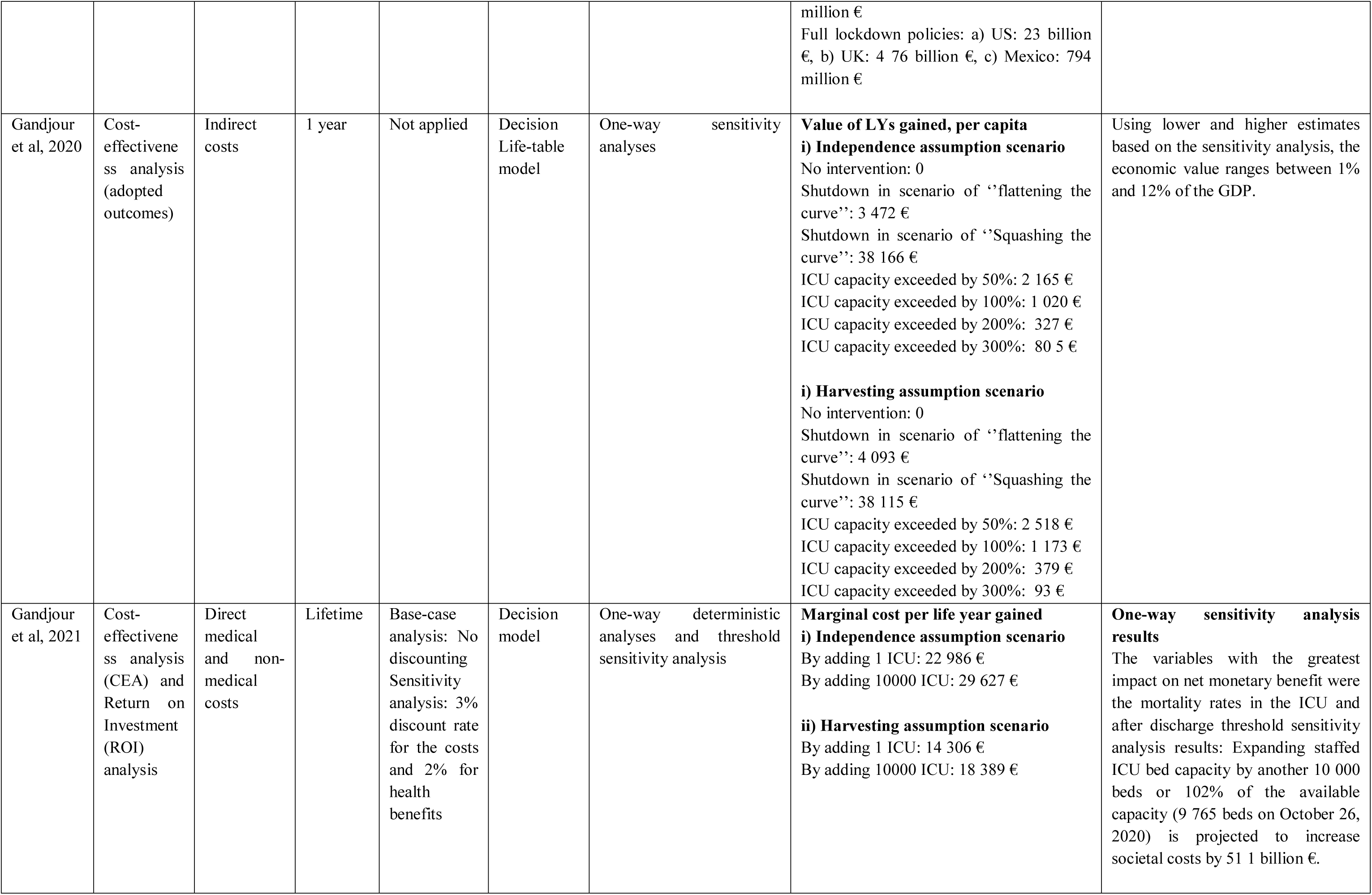
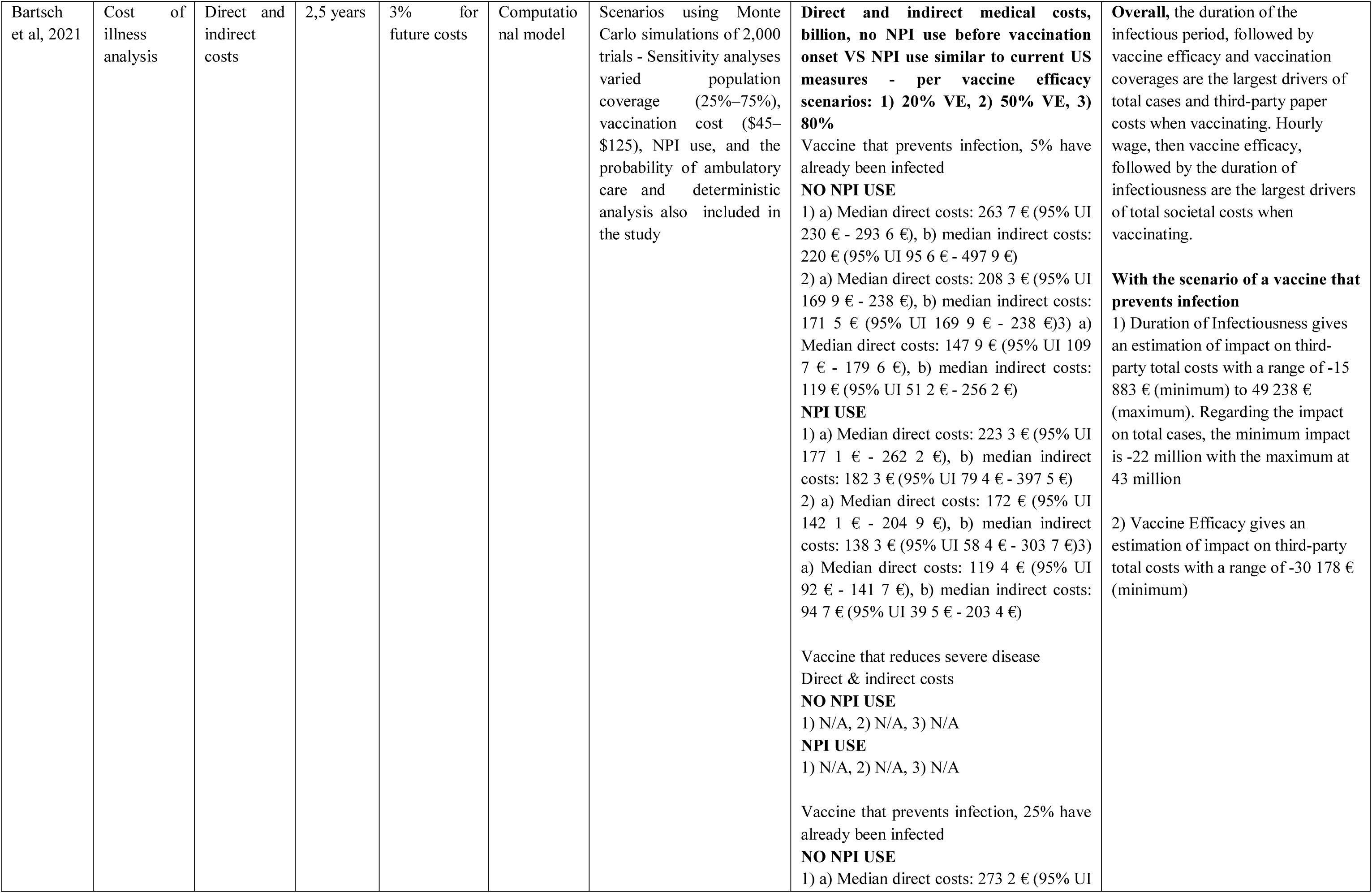
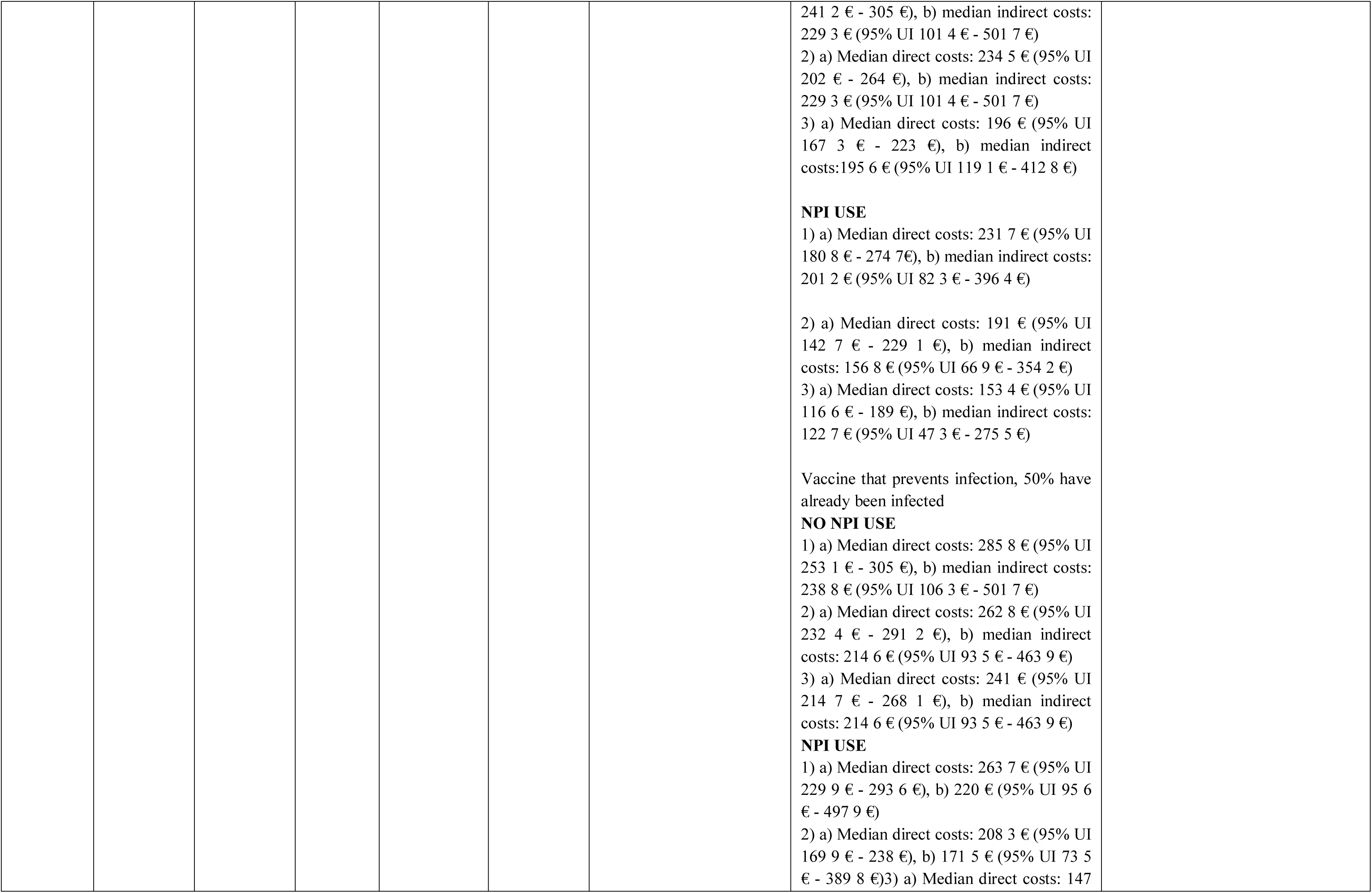
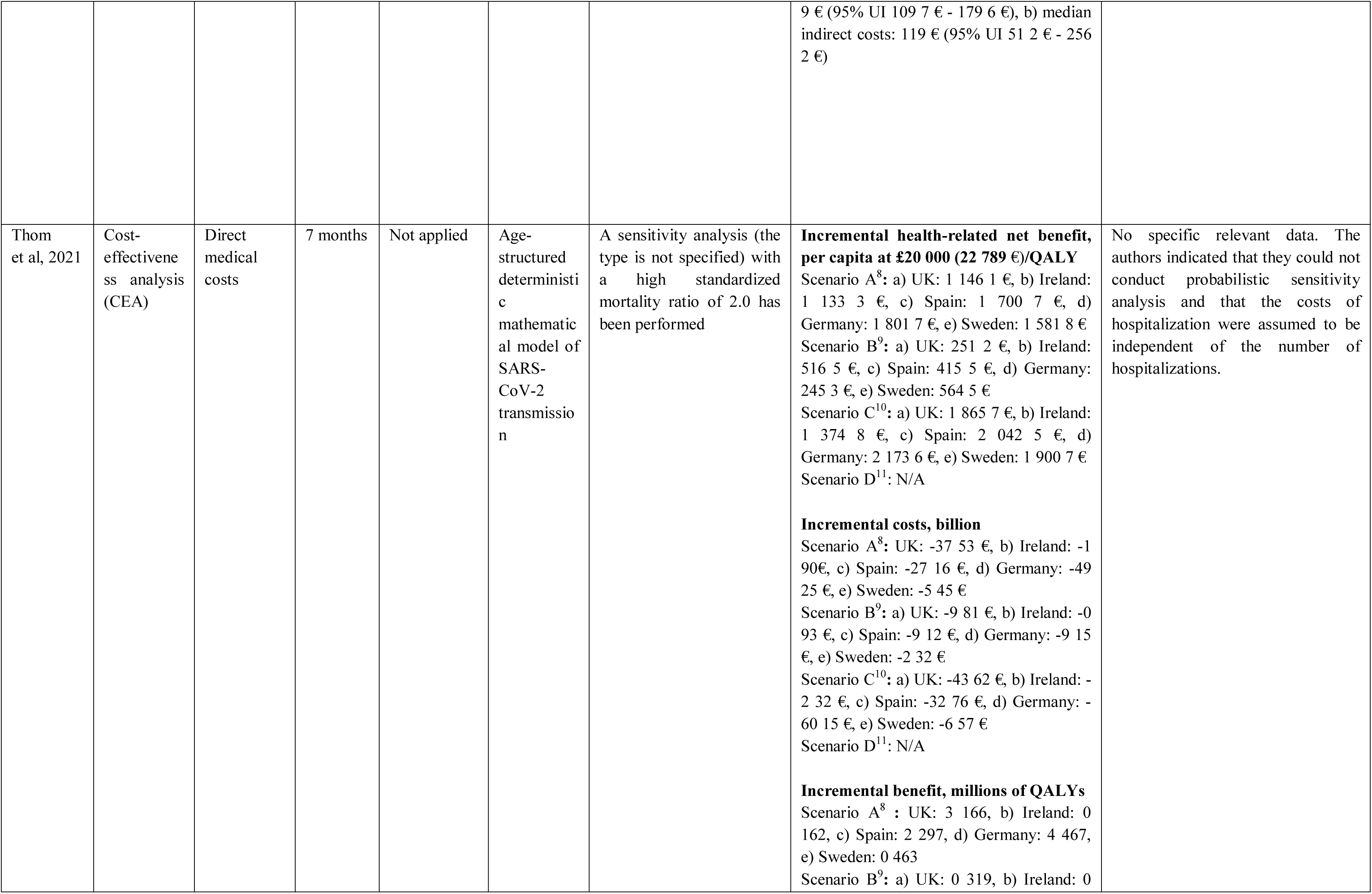
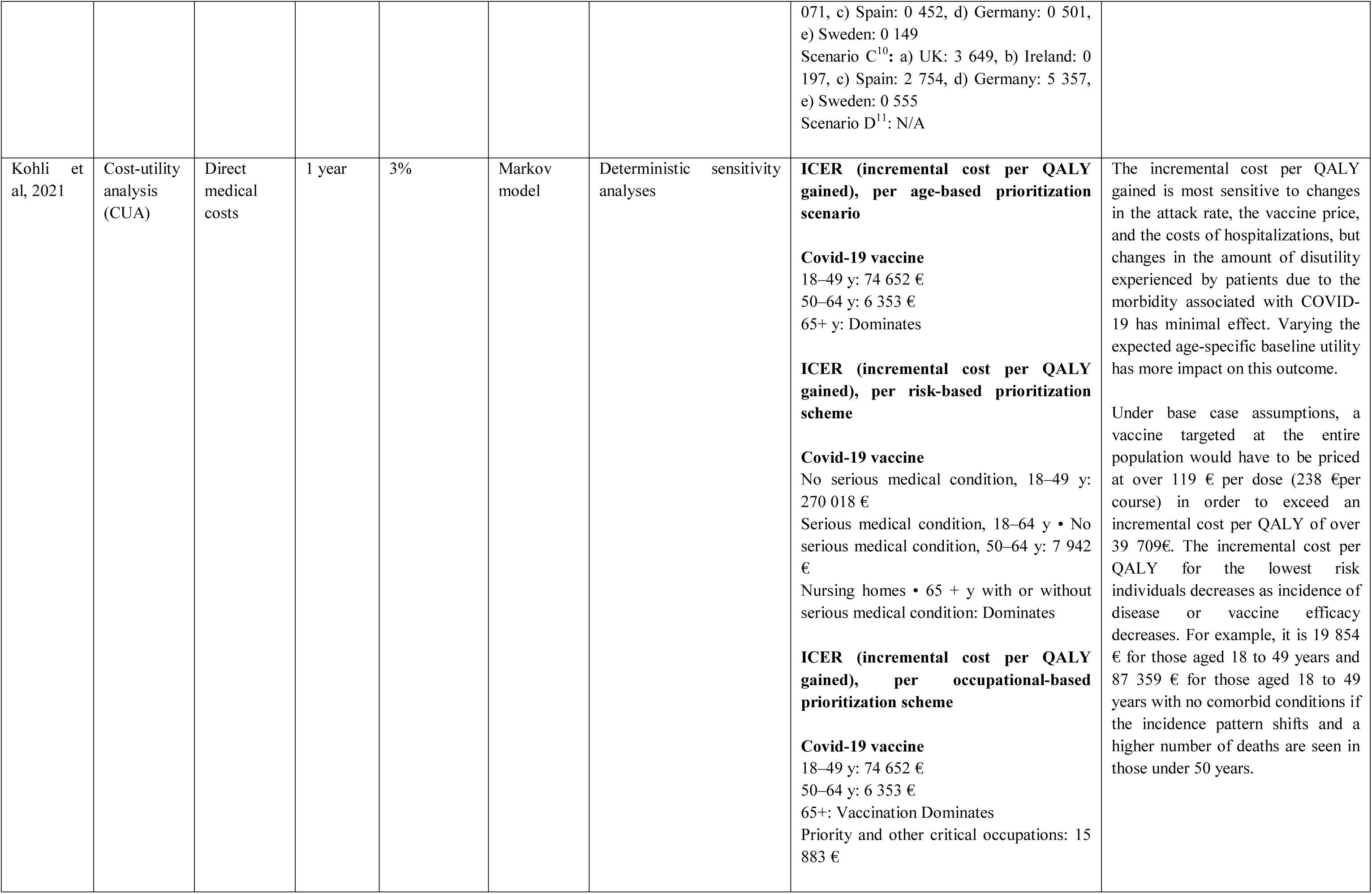
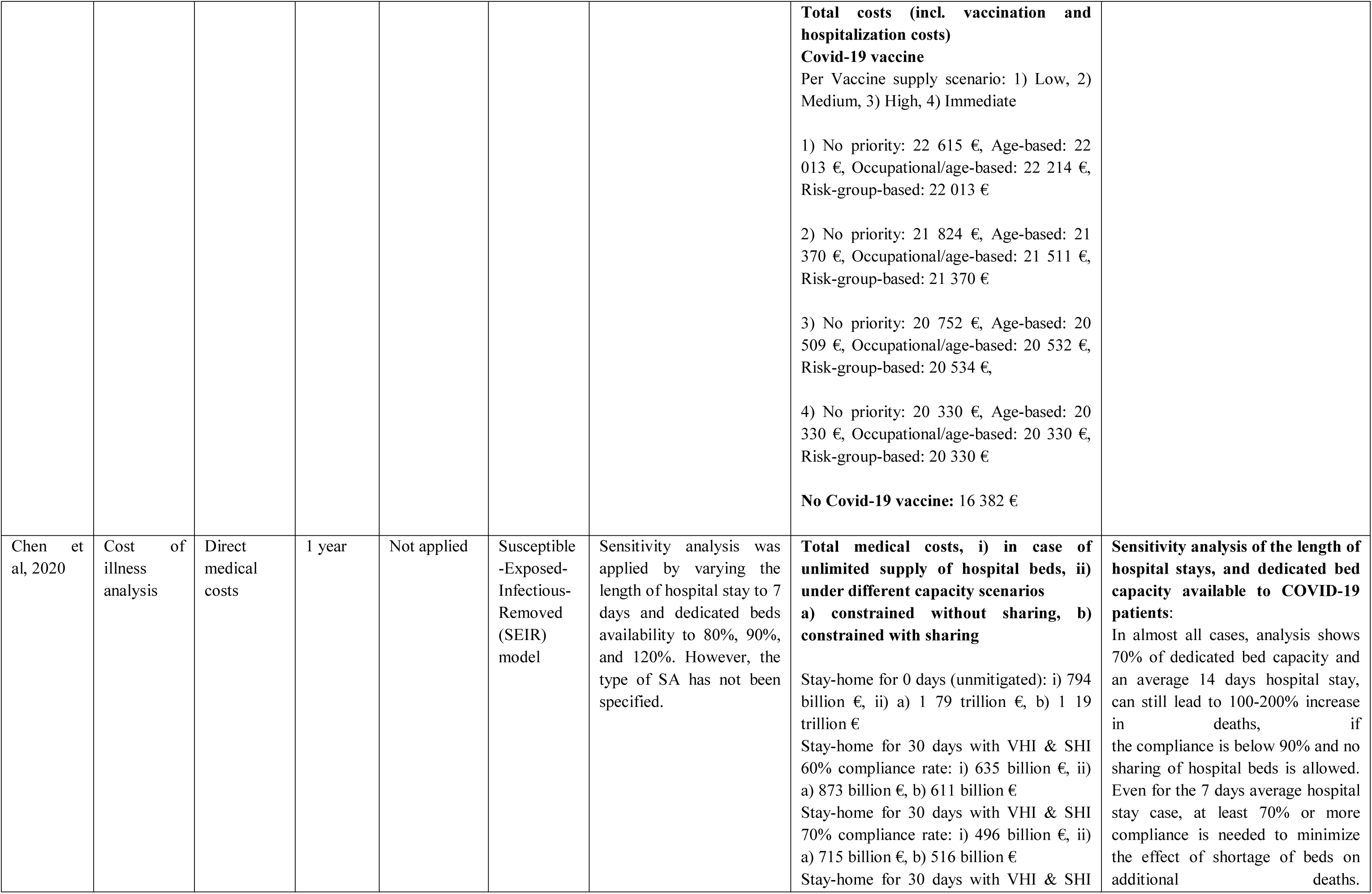
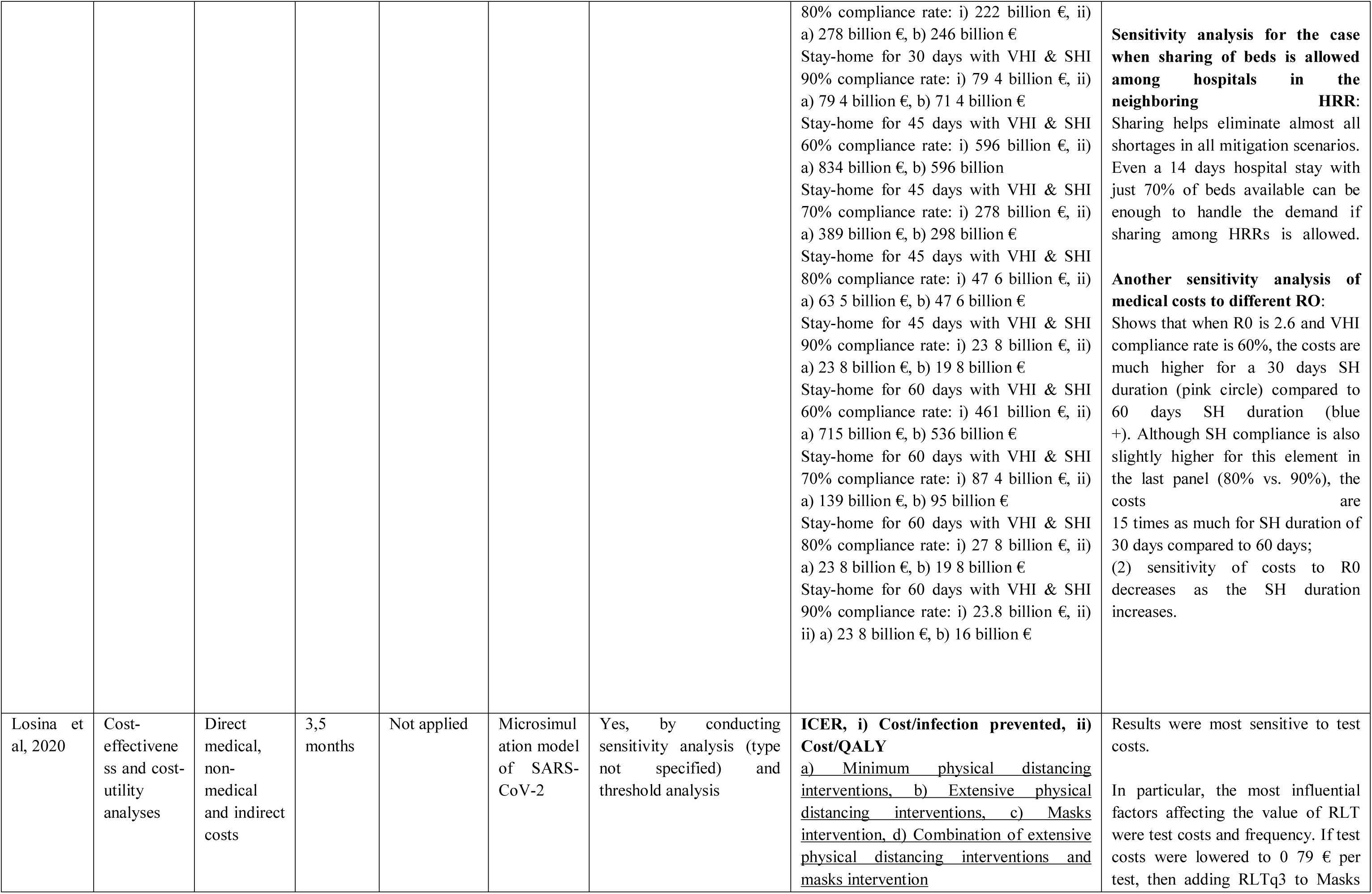
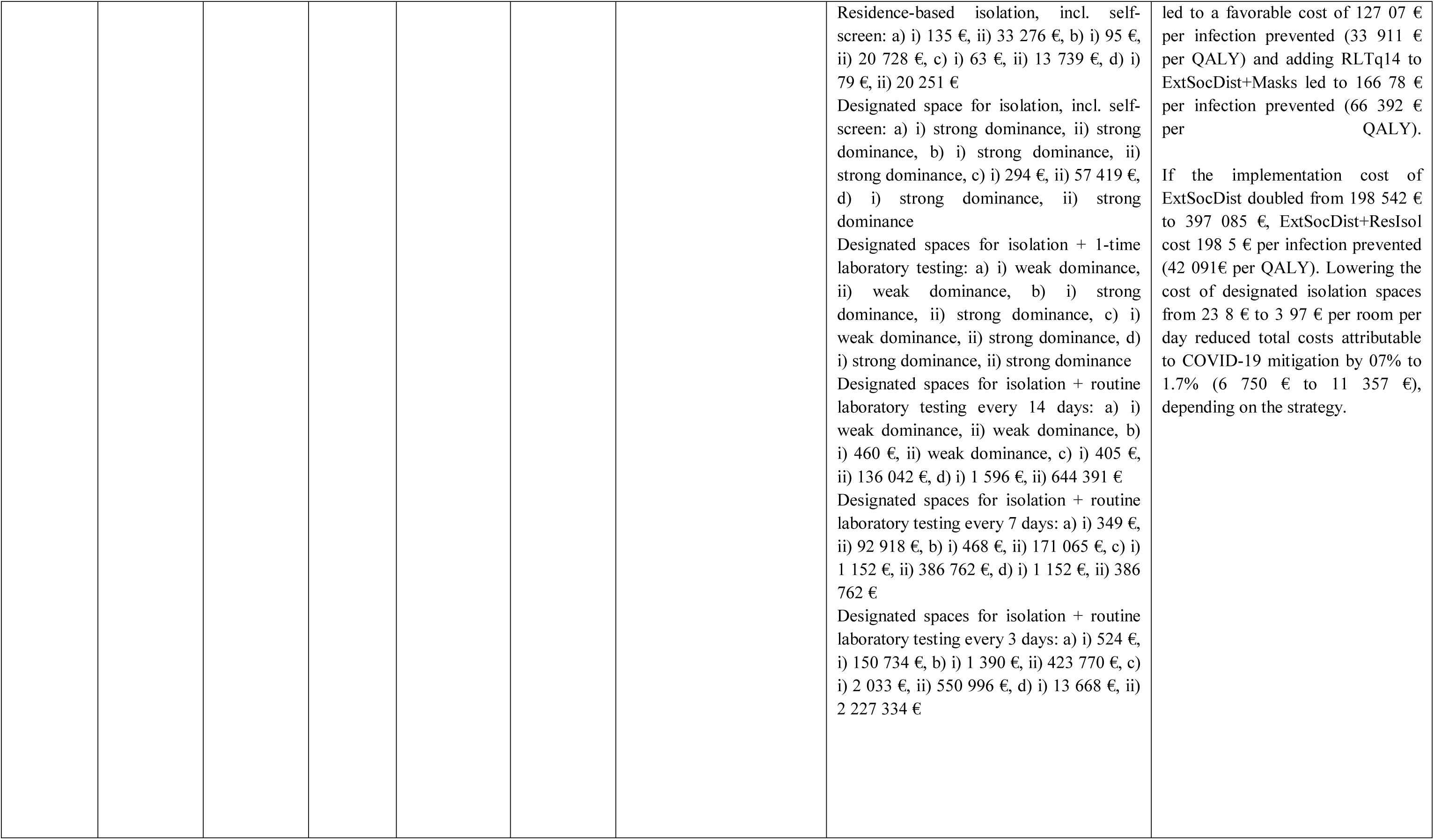
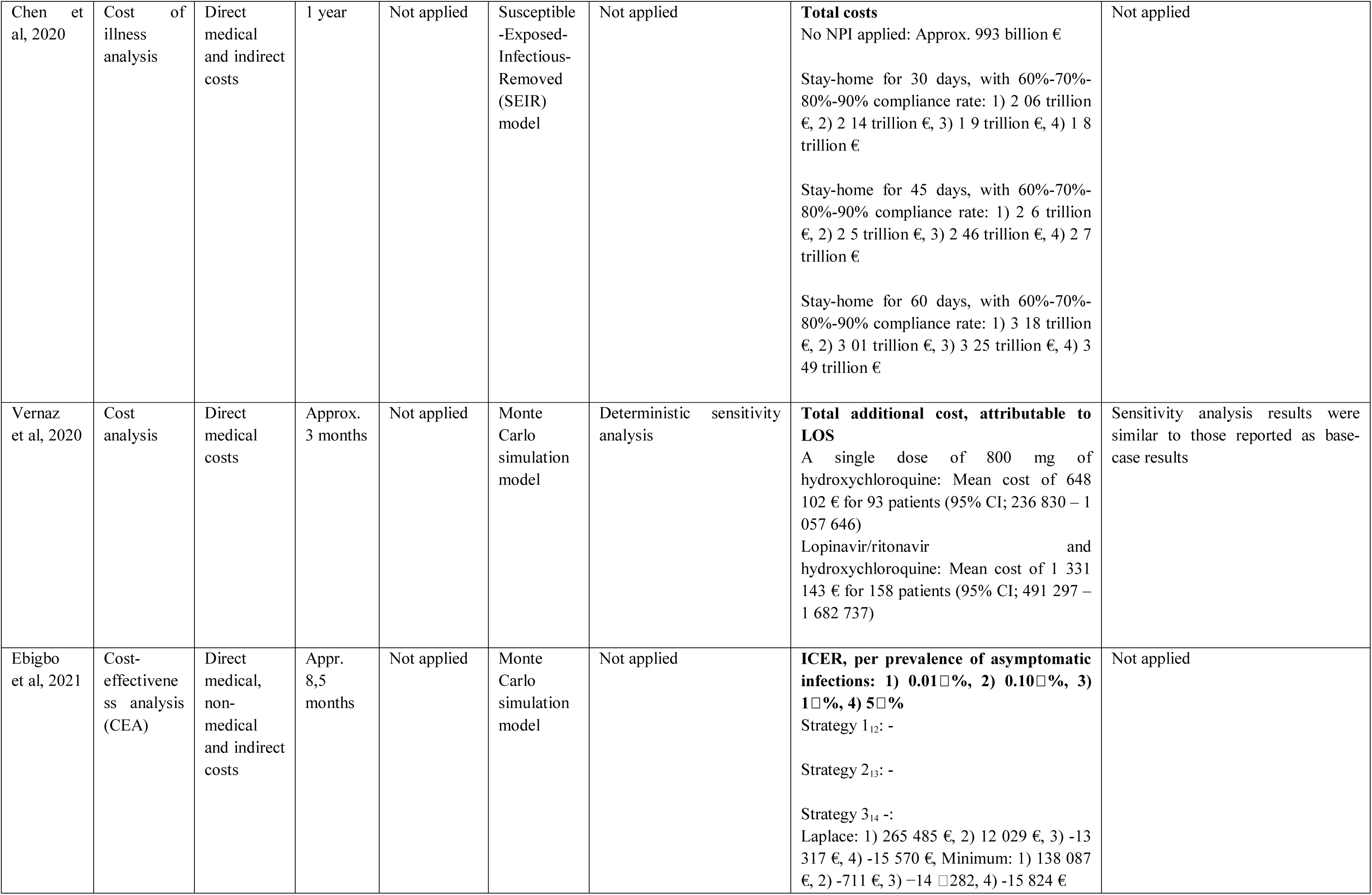
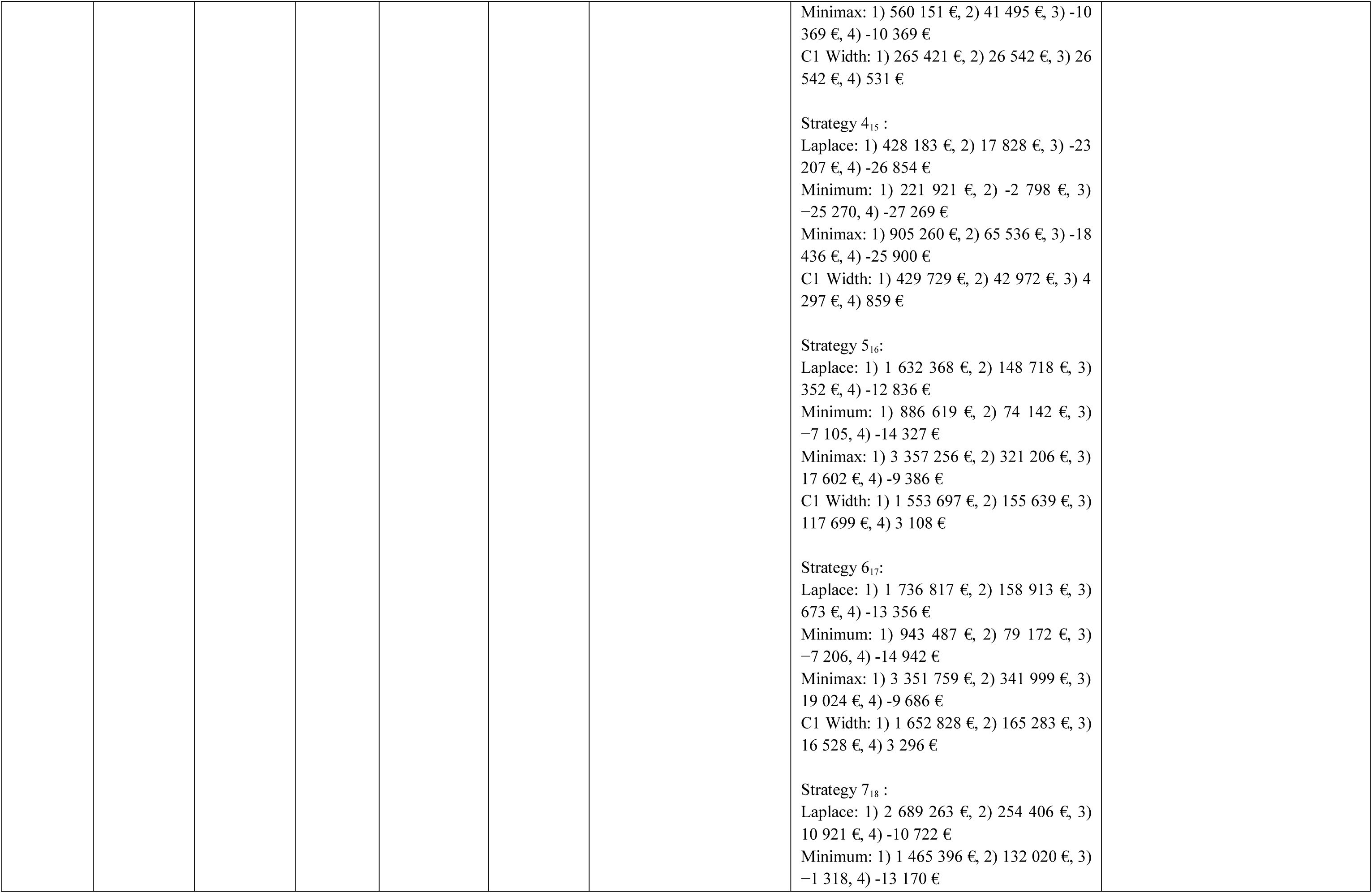
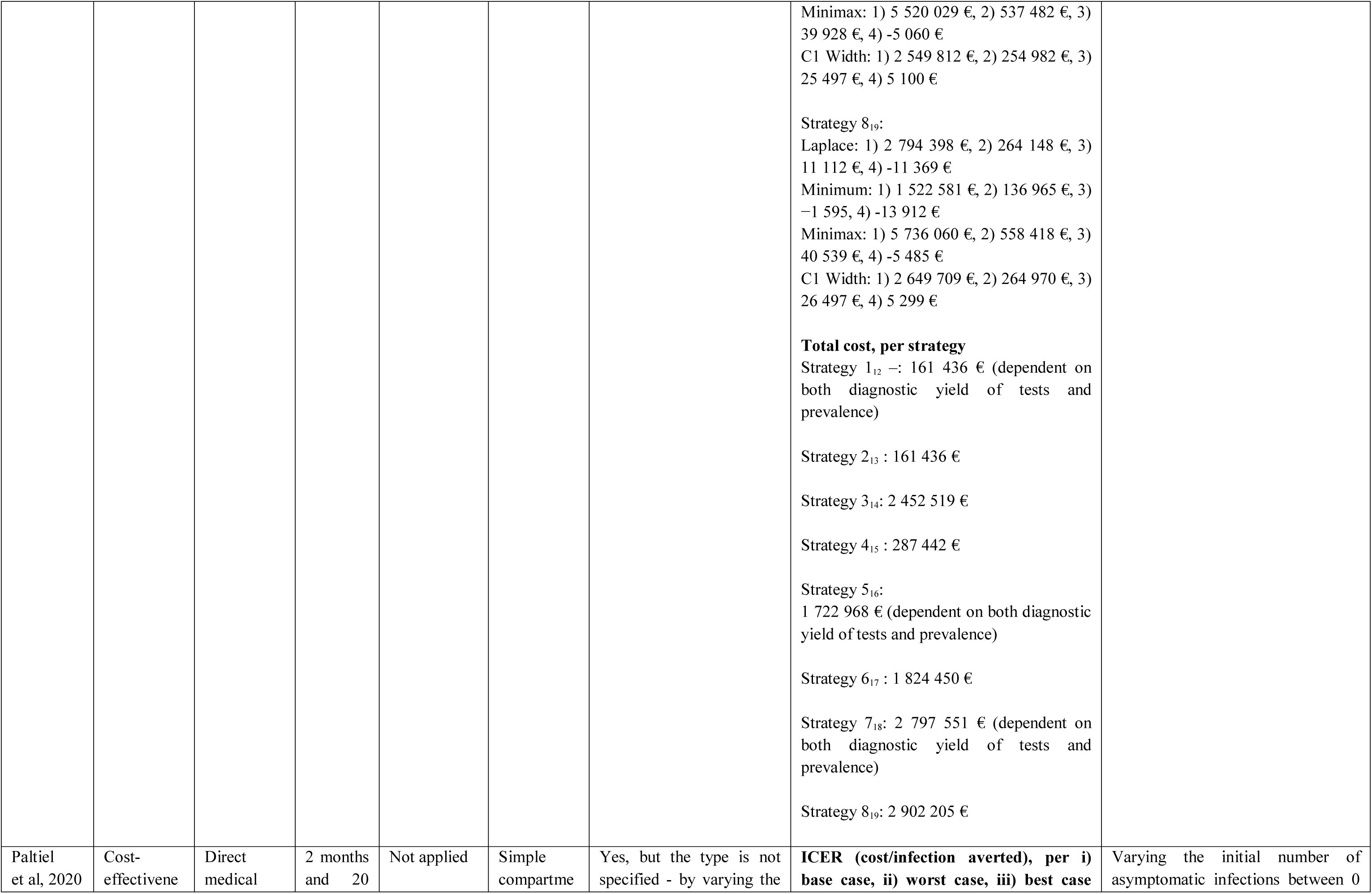
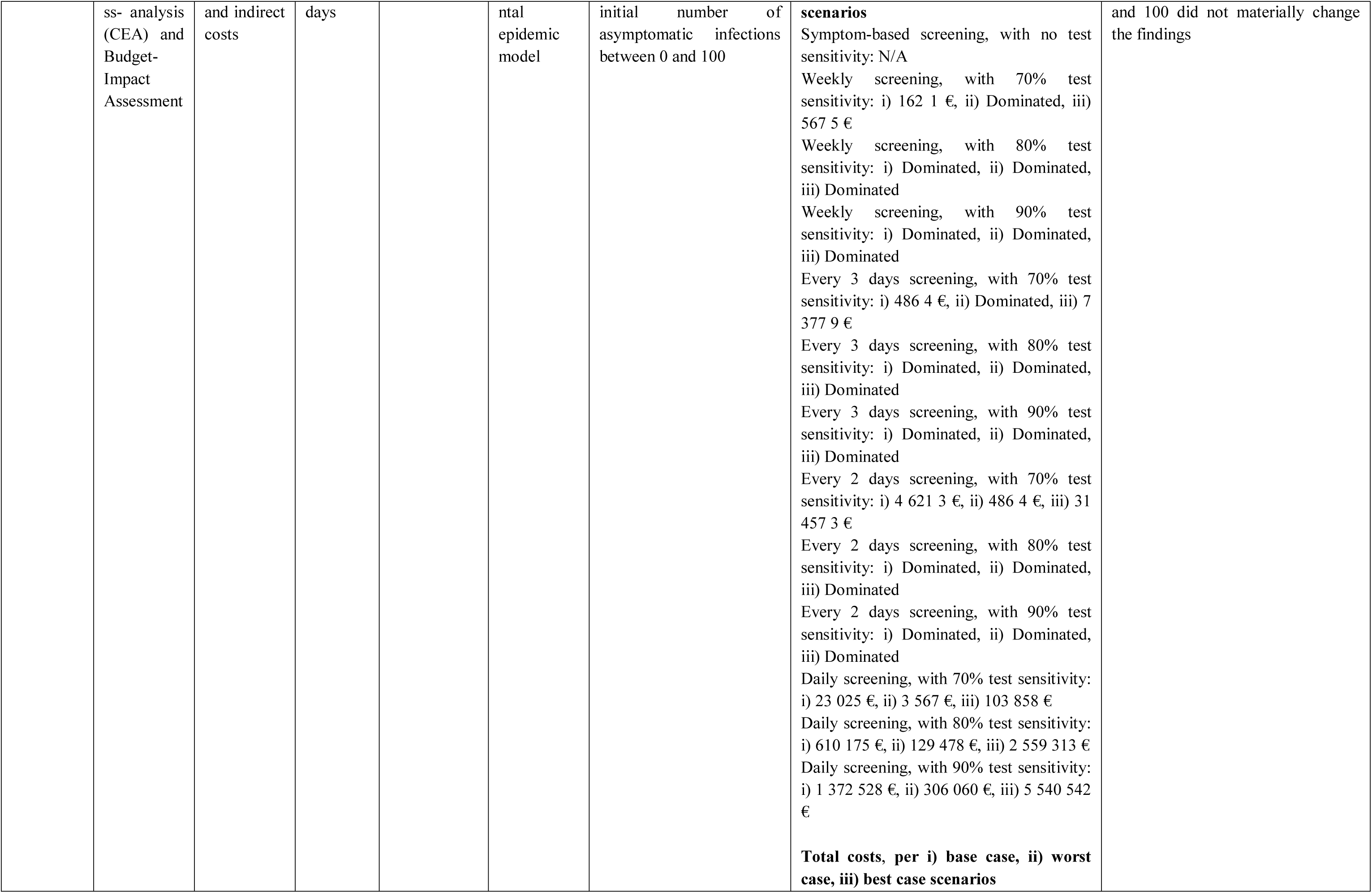
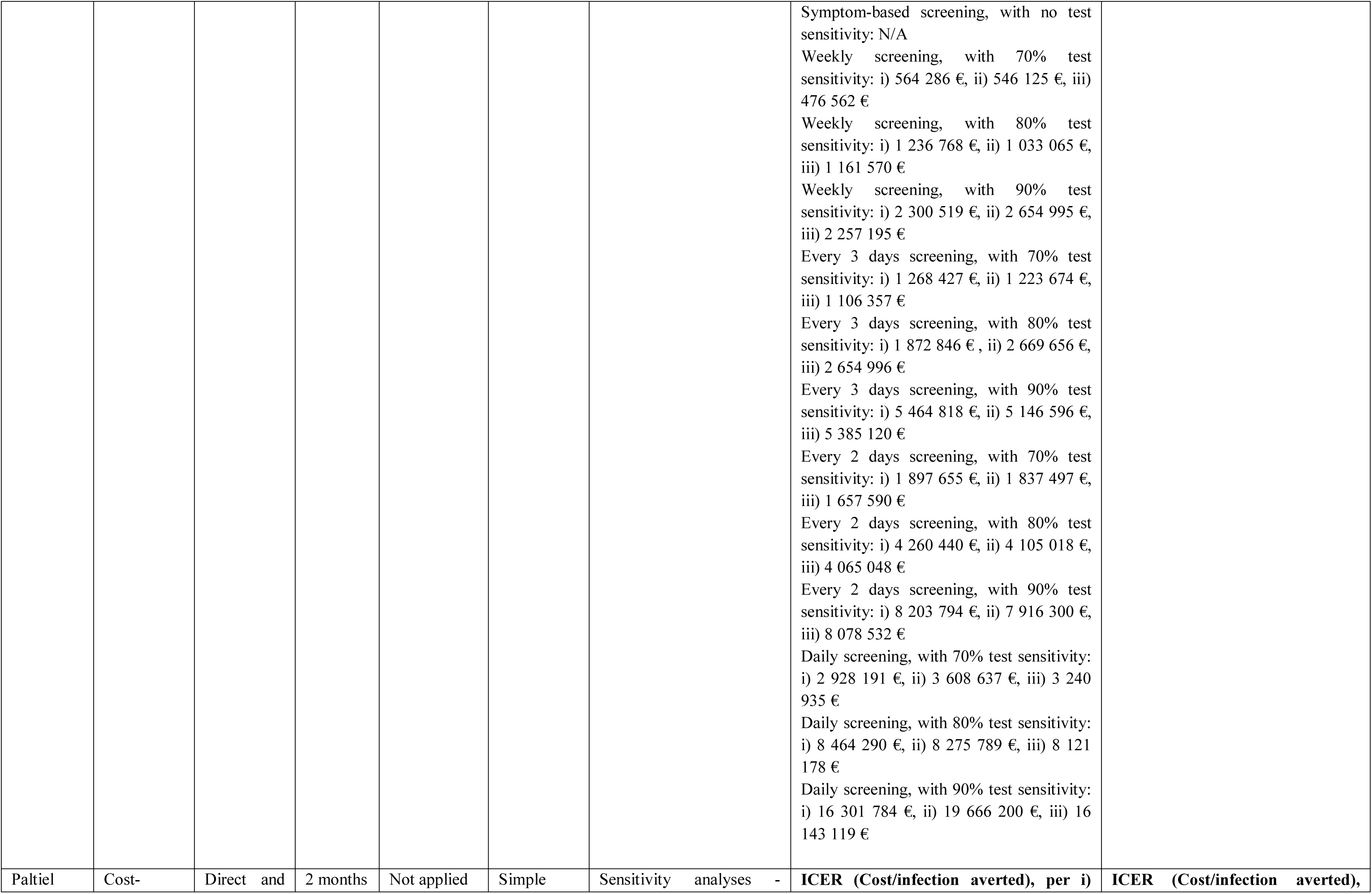
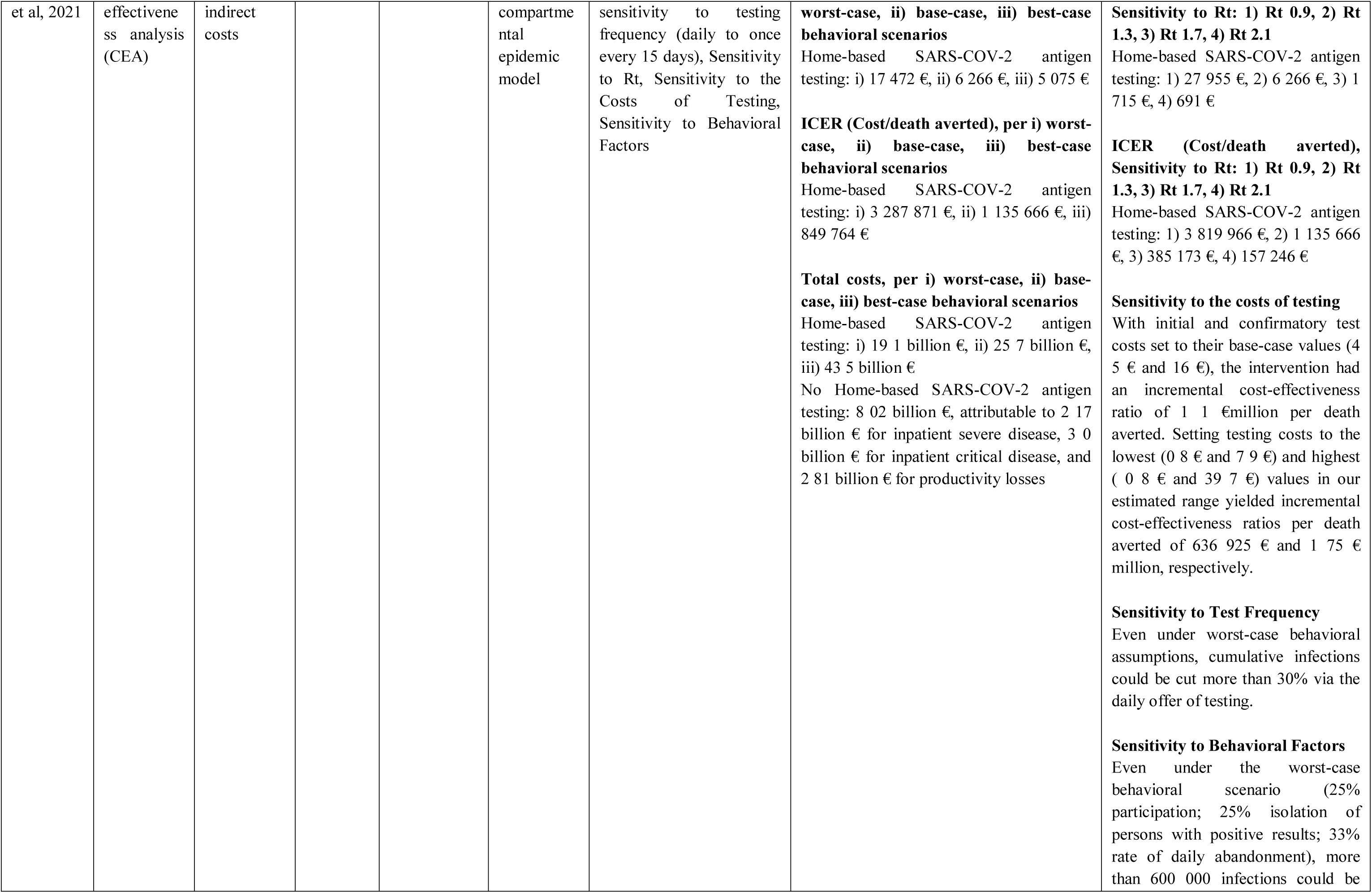
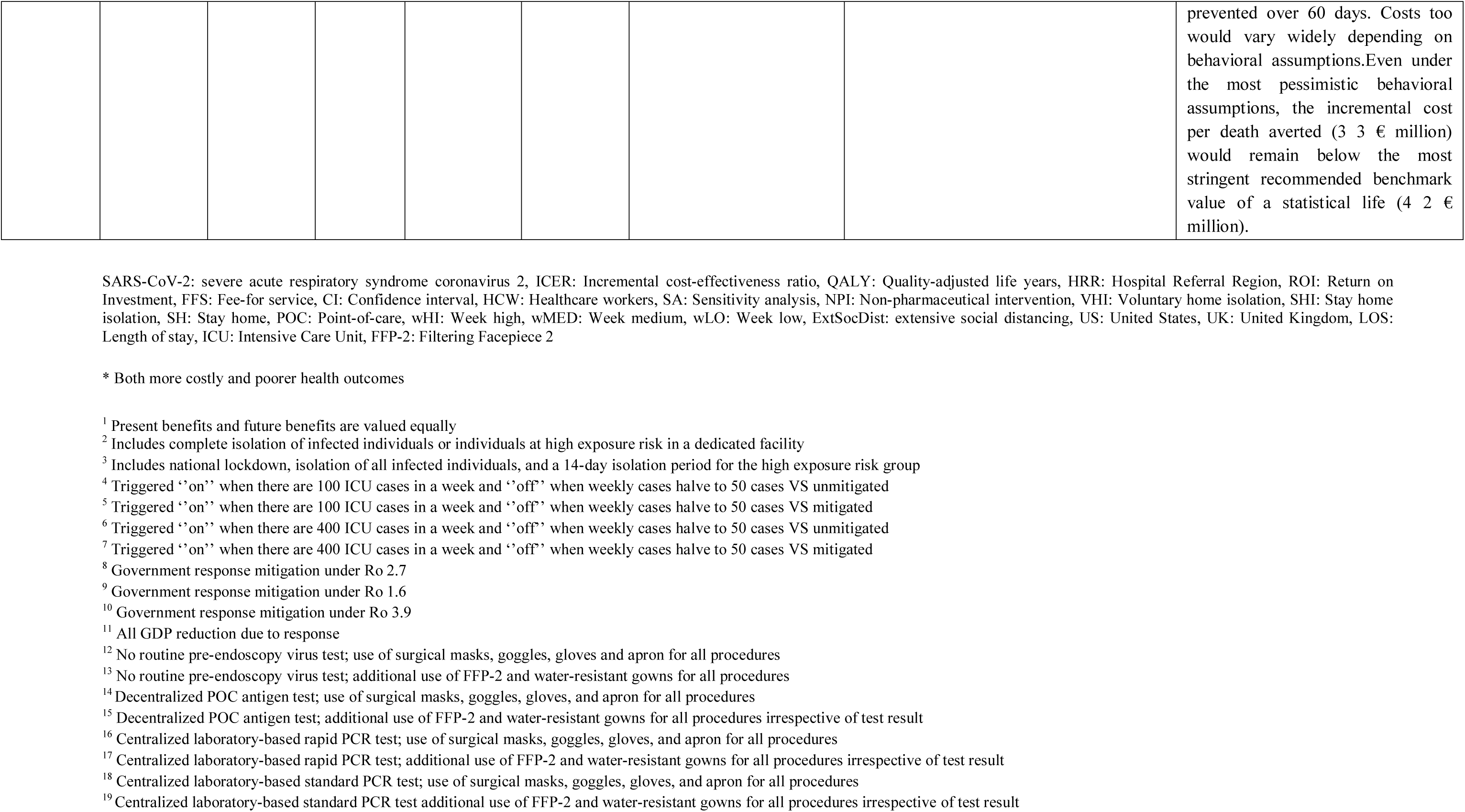
Economic evaluation outcomes concerning interventions and strategies to mitigate Covid-19 (n=31)

Two studies appraised the cost-effectiveness outcomes of different testing strategies within specific settings. Baggett et al. (2020) (40) investigated the clinical outcomes, costs, and cost-effectiveness associated with strategies for COVID-19 management among adults experiencing sheltered homelessness in Boston, USA. Daily symptom screening with subsequent PCR testing of individuals who screened positively was the most efficient strategy and was cost-saving relative to no intervention in all epidemic scenarios.

Paltiel et al. (2020) (41) examined the SARS-2 screening performance standards that would permit the safe return of students to US residential college campuses for the fall semester of 2020 and noted that screening with a less expensive, less sensitive test dominated the intervention of screening with more expensive, more accurate tests. Paltiel et al. (2021) (36), assessed the clinical and economic effects of widespread home-based SARS-CoV-2 antigen testing and noted that the cost per infection averted was 6,266 €.

#### Cost-effectiveness of quarantine, isolation, physical distancing, and physical restriction policies

Twelve studies were included to assess the cost-effectiveness across different strategies to prevent or mitigate SARS-COV-2 transmission (**Table 4**).

Kouidere et al. (2021) (35) conducted a mathematical compartmental modelling to assess three different strategies for diabetics and concluded that awareness programs and quarantine for those infected having symptoms as well as those having complications in hospitals with treatment was the most effective strategy and associated with an incremental cost-effectiveness ratio (ICER) of -0.6951 (cost per infection averted) compared to sensitisation and prevention.

The cost-effectiveness of comparing the national lockdown policy for the susceptible population with the isolation of those infected or being at high risk in Israel returning to the workforce under social distancing measures after a 14-day isolation period was appraised by Shlomai et al. (2020) (34). The national lockdown of the susceptible population was associated with a higher economic burden but contributed to a reduction in deaths making this intervention superior to the comparator only in terms of health outcomes but was not cost-effective since it contributed to high economic costs, estimated at 36,568,451 € costs per death averted, and 3.6 million € costs per QALY gained. Furthermore, Keogh-Brown et al. (2020) (33) assessed the direct and indirect costs of three different strategies in the UK, and indicated that although mitigation and suppression policies contribute to lower mortality rates, the economic impact of COVID-19 is likely to be dominated by public prevention measures rather than the direct health costs of the disease and indicated thata three-month mitigation scenario resulted in a prediction of 13.5% loss to GDP with the direct-health related economic impact of 2% of the GDP while the suppression scenario was associated with an approximately 22% loss in GDP. Furthermore, Miles et al. (2020) within the context of a cost-benefit analyses (32) noted that a full lockdown policy for four months followed by an extension for three months showed that net extra economic costs of the lockdown relative to the easing of restrictions was estimated at 116.2 billion € with the best scenario of lives not lost at 96.5 billion €. The main factors of the substantial costs in lockdown policy were associated mainly with employment restrictions and physical restrictions. Miles et al (2021) also (31) conducted an additional modelling and simulation study to assess a few physical restriction scenarios in a timeframe of 6 months in the UK and found that the strategy of relatively slowly easing restrictions comes at a cost in terms of GDP reduction of up to 697,121 € /life□year saved. Moreover, the cost per life-year saved with a short transition physical restriction policy (8 weeks medium then 18 weeks low) was associated with a cost of per life-year saved at 206,888 € while keeping the policies strict for 26 weeks was estimated at a cost of per life-year saved of 1,553,988 €.

Suppression and physical distancing interventions was assessed by Zala et al. (2020) (30) and Barnett-Howell et al. (2021) (29). Zala et al., noted that suppression policies compared to an unmitigated scenario strategy was associated with an ICER below 56 972 €, while all of the four suppression policies presented ICER results. Barnett-Howell et al. (29) investigated five physical distancing strategies, including an unmitigated intervention, to assess the value of statistical life lost. Full lockdown policies were associated with lower VSL in the US, UK and Mexico countries (US: 23 billion €, UK: 4.76 billion €, Mexico: 794 million €) while the no-action scenario, which was the unmitigated spread of COVID-19, contributed to higher VSL in those countries (US: 38.9 billion €, UK: 7.1 billion €, Mexico: 1.58 billion €).

Additional studies focusing on policies related to stay-home and shutdown orders, were investigated by Gandjour (2020) (28) and Thom et al. (2021) (27). Gandjour assessed the clinical and economic cost of a shutdown during the SARS-CoV-2 pandemic in Germany to estimate the cost of life-years gained per capita estimating the indirect costs for one year. The results showed that economic cost of the intervention of lockdown was associated with 3,472 € or extrapolated to the total population, 8% of Germany’s GDP in 2019 (ranging from 1% to 12% in the sensitivity analysis). Thom et al. (27) provided an exploratory comparison of health-related benefits and costs saved by government mitigation measures across European countries (UK, Ireland, Germany, Spain and Sweden) over a timeframe of seven months, where the authors concluded that the benefit of government COVID-19 responses might outweigh their economic costs, saving millions of QALYs (0.5 million for Sweden to approximately 5 million for Germany) and ideally those countries with more mask-wearing and testing, had better outcomes.

The last studies focusing on stay-home policies were by the same authors, Chen et al., (2020) (25, 26). Their first study (25) estimated the total medical costs by keeping the US economy open and assessed 13 interventions with different stay home isolation and voluntary home isolation compliance rates for one year. Without mitigation, the total medical costs would be a 5% of the US GDP. In their second study (26), Chen et al compared the epidemiological and economic impact of COVID-19 spread in the US under different mitigation scenarios, comprising NPIs including stay-home with varying compliance rates. The study results indicated that the preferred mitigation scenario in terms of lives saved, and infections averted is when the compliance is at 90%, the stay at home duration is 45 days; with an economic impact of 2.76 trillion €.

#### Cost-effectiveness of vaccination measures

Three included studies (44–46) focused on vaccination interventions used modelling and simulation methods to assess the cost-effectiveness of the included interventions (**Table 4**). Shaker et al. (2021) (44) appraised the universal COVID-19 vaccination versus risk-stratified vaccination approach over the timeframe of one year, and they found that the universal COVID-19 vaccination dominates the risk-stratified approach, making it a cost-effective strategy for the US population from the societal perspective and the healthcare perspective. ICER analyses indicated that in the case of societal perspective, the universal approach presents a cost of 52,033 € per death averted compared to the risk-stratified vaccination , with cost-savings estimated at 395 million €. When considering the healthcare perspective, the cost per death averted of the risk-stratified approach was 52,575 € making it the dominant option. Bartsch et al. (2021) (45) noted that that NPI implementation before vaccination was associated with less total costs compared to the case of non-implementation of NPIs. Finally, Kohli et al. (2021) (46) conducted a CUA to assess the COVID-19 vaccine vs. no vaccine for the US population over one year. Base case analysis presented good value for money outcomes where the ICER per age was found to be 74,652 € for ages 18-49 y, 6,353 € for 50-64 y while for ages 65 + the COVID-19 vaccine dominates. This strategy was also cost-effective under additional scenarios, such as with the per risk-based prioritisation scheme.

#### Cost-effectiveness of treatment measures

Our analysis included four studies (47–50) assessing pharmaceutical interventions (**Table 4**). Lee et al. (2021) (48) simulated the transmission spread of SARS-CoV-2 and the economic and clinical impact of the spread in the US over one year. The authors noted that starting treatment at an of R_e_3·5 was cost-effective and provided ICER results of 2,690 €/QALY saved from the societal perspective when treating 75% of symptomatic cases and treatment course costs 397 €, and ≤6,267 €/QALY saved from the third-payer perspective when at least 50% of all patients were treated. The study also concluded that treating 25% or 50% of symptomatic patients at R_e_ 2·5 and at a treatment cost of 397 €, was also cost-effective. Accross all R_e_ assessed, the net cost-savings were high from the third-payer perspective and even higher from the societal perspective. Sheinson et al. (2021) (50) assessed the cost-effectiveness of treating hospitalised COVID-19 patients with COVID-19 treatment vs. treating them with the best supportive care (BSC). Under the payer perspective, treatment of patients with COVID-19 was more cost-effective than BSC as it was associated with 15,784 € (with a FFS payment) and 18,593 € (with a bundled payment). The ICER results from a societal perspective were even better, making the COVID-19 treatment process a dominant strategy as treatment of patients with COVID-19 was associated with a cost of 6,508 € (with FFS payment) and 9,317 € (with bundled payment).

With regards to specific pharmaceutical regiments, Águas et al. (2021) (49) estimated that the incremental cost per life-year gained for dexamethasone treatment was 1,071 € compared to no treatment. On the contrary, within a cost-analysis performed by Vernaz et al. (2020) (47) compared standard care among pharmaceutical interventions including lopinavir/ritonavir and hydroxychloroquine (a pharmaceutical discussed early in the pandemic but not a legitimate treatment) for hospitalized patients with SARS-CoV-2 in Switzerland. The results highlighted the additional costs attributable to the length of stay presented a mean cost of 648,102 € for 93 patients.

#### Cost-effectiveness of bed provision policies

We identified one study indicating the scenario of increasing ICU bed availability . Gandjour et al. (2021) (55) performed a cost-effectiveness and return on investment (ROI) analysis and estimated that that the provision of a staffed ICU bed reserve capacity was cost-effective even for a low probability of bed utilisation and estimated that the provision of 1 ICU bed would cost 14,306 € to 22,986 € per life-year gained, while the addition of 10,000 ICU beds provided cost 18,389 € to 29,627 € per life-year gained.

#### Cost-effectiveness of personal protective equipment strategies

Risko et al. (2020) (43) conducted a cost-effectiveness analysis indicating that investing in personal protective equipment (PPE) was a cost-effective option for 166 million simulated workers over a period of 7.5 months (**Table 4**). In general, they found that an investment of USD 9.6 billion (7.62 billion €, 2021) concerning the purchasing and distribution of PPE to allow for adequate protection of all HCWs was cost-effective and lead to an ICER of cost per case averted at 57 €, and an ICER of cost per death averted of 4,159 €, which translates to 41.1 billion € (95% CI 39.6 to 42.6) in economic gains.

#### Cost-effectiveness of other combined strategies

The review included four studies (51–54) that assessed multiple combined interventions related to testing, isolation, vaccination and physical distancing strategies (**Table 4**). The value of immunisation alongside physical distancing policies was evaluated by Sandmann et al. (2021) (53), who assessed the clinical and economic value of COVID-19 vaccination in the UK and estimated that without the initial lockdown, vaccination, and increased physical distancing, 3.1 million (0.84 – 4.5) deaths would occur in the UK over ten years with a cost of 169 million €. The introduction of vaccination provided estimations of incremental net monetary values ranging from 13.7 billion € to 381.4 billion € in the best-case scenario and from 1.25 billion € to 64.8 billion € in the worst-case scenario compared to no immunization –under the healthcare perspective. In regard to the wider societal perspective, where GDP income loss was included in the relevant analyses, the incremental net monetary value of introducing vaccination vs. no vaccination was found to be extremely positive across physical distancing scenarios. Pandey et al. (2021) (54) assessed the cost-effectiveness of eight testing measures in conjunction with isolation measures over a timeframe of five months. In cases of an R_0_=2 and assuming the test cost of approximately 4 € and the societal willingness-to-pay (WTP) per year-life lost (YLL) averted of 79,417 €, the optimal strategy was daily testing plus a 2-week isolation. On contrary, at a lower R_0_ (1.5 - 1.8) weekly testing plus 1-week isolation was found to be the optimal strategy. Furthermore, the weekly testing plus 2-week isolation was also optimal if R_0_ >2 (under the same assumptions and when the test costs <318 €).

The combination of isolation with laboratory testing and self-screening in college campuses over 3.5 months was assessed by Losina et al. (2020) (52), who concluded that a comprehensive physical distancing policy with a mandatory mask-wearing policy can prevent most COVID-19 cases on college campuses and is relatively cost-effective. The last study that assessed multiple interventions was that of Ebigbo et al. (2021) (51) that appraised the use of low vs. high-risk PPE (masks, goggles, gloves and apron, and centralised laboratory-based testing) applied during routine pre-endoscopy procedures. The authors noted that routine pre-endoscopy testing combined with high-risk PPE becomes more cost-effective with rising prevalence rates of COVID-19 while in lower prevalence rates the dominant strategy appeared to be the intervention of point-of-care antigen test without routine high risk PPE use.

## DISCUSSION

This systematic review aimed to assess the economic burden of COVID-19 infection to societies and healthcare systems and to assess the strategies used to prevent and mitigate COVID-19 outbreaks in the EU/UK/EEA and OECD countries from studies published through 22^nd^ April 2021. Our findings indicate that the overall economic impact of the virus is substantial for individuals, healthcare systems and payers, while our review identified that NPIs and pharmaceutical measures, including ICU bed provision implemented within the context of the COVID-19 pandemic from both healthcare and societal perspectives and within one year time horizon are cost effective response and mitigation measures.

Overall, in the studies identified, the economic burden of COVID-19 pandemic was found to be substantial in all studies included in the systematic review, with both direct and indirect costs playing a significant role. Direct costs were primarily attributed to medical expenses from hospitalisations and ICU admissions while the indirect and societal costs yielded by NPIs, mainly from stay-home and isolation strategies, contributed to the further increase of economic costs and also resulted in a decrease in GDP. Moreover, the delays in treatment initiation of other diseases (e.g., cancer) were also found to have substantial economic impact. At the patient level, increased medical costs were also related to comorbidities such as obesity and diabetes. Regarding indirect costs, temporary and permanent productivity loss, as well as human lives lost due to COVID-19 were substantial.

The NPIs implemented for the pandemic control led to a benefit in life years in an individual or societal level compared to the no intervention scenario, although the cost benefit of such interventions differed depending on the perspective, the timeframe, the setting and the epidemiological situation of the pandemic. Considering the testing strategy, results were dependent on the cost per test and the R_0_ at the time of assessment. Overall, low-cost repeated community screening was a cost-effective approach, in combination with other NPIs, especially when the cost of testing remains low and at higher R_e_. As for lockdown strategies, studies showed that if performed early in the pandemic for a limited period of time and with sufficient compliance, they substantially could reduce the medical costs of COVID-19 from a healthcare perspective – especially prior to population immunisation. In general, mitigation scenarios resulted in less GDP loss compared to suppression ones. Finally, quarantine and physical distancing strategies were found to be effective for the containment of the COVID-19 pandemic, while it was indicated that with an increasing R, a combination of NPIs, including screening, physical distancing, and quarantine of contacts would be more efficient. With regards to PPMs they showed a benefit both in health and costs when used by HCWs, as also was the provision of ICU beds. Regarding pharmaceutical measures, vaccination was consistently found to be cost-effective, with the universal COVID-19 vaccination dominating the risk-stratified approach. Additionally, pharmaceutical treatments were also cost-effective when provided in scenarios of high transmissibility (high R_e_). Finally, the combination of testing, vaccination, physical distancing measures and mask wearing was found to be cost saving, with a significant number prevented cases and deaths.

A systematic review on previous respiratory infectious disease outbreaks prior to COVID-19 (56) concluded to similar results by pointing out the significant burden of both direct and indirect medical costs for management and response activities. Most direct costs occured from the additional personnel hours, the response planning and contact tracing activities, the provision of training and educational materials and the laboratory costs. Indirect costs were greater than direct ones, particularly when school closures and/or workplace closures were implemented, due to lower productivity (56). However, given the strictness of the NPIs in COVID-19 pandemic the economic burden has been found to be high for primary production sectors including industries associated with activities in raw materials extraction, secondary industrial sectors involving the production of finished products and tertiary sectors encompassing service provision industries (57).

This systematic review provides a wide range of cost-effective options across comparative strategies implemented to prevent or mitigate virus transmission. Strategies, including vaccination measures (ideally universal vaccinations), screening policies (with the saliva sampling to be a cost-saving option compared to nasopharyngeal swabs) and expanding a staffed ICU bed reserve capacity were found to be dominant strategies against SARS-COV-2 transmission, indicating the cost-savings as well as the economic value of their implementation. An earlier systematic review by Vandepitte et al. (2021) also showed that frequent and universal testing activities are a cost-effective strategy, highlighting that it would have a greater impact if enacted in a setting with a high R_e_. Moreover, they noted that PPM was also a cost-effective strategy depending on the compliance, the context, and the R_e_. (58). Similarly in another review, contact tracing and isolation of cases was a cost-effective NPIs, along with adequate surveillance, PPM for healthcare professionals, and vaccination. Contrastingly, physical distancing strategies including school and workplace closures were found to be effective but costly, making them the least cost-effective options. Additionally, combined NPIs were more cost-effective than individual ones, while the significance of early implementation was emphasised (59).

### Strengths and limitations

This review provides several strengths, including covering within one review both the economic burden of COVID-19 and the cost-effectiveness of the strategies and programs implemented to mitigate the pandemic. Moreover, this review followed a systematic approach to study identification, data extraction and quality apprasial with most of the included studies of good or high quality. Furthermore, this study used the Dominance Ranking Matrix approach, which summarised and interpreted the results of economic evaluation studies. On the other hand there are some limitations, as publication bias can not be excluded and as our search was performed up to the end of April 2021 it only reflects the cost-effectiveness of interventions assessed during the first waves of the pandemic with the majority of the populations unvaccinated, while most studies have a short duration on which modelling was performed. A further limitation is that most studies estimate costs and benefits based on a health care perspective, excluding wider societal effects, with a time horizon of 1 year. As we restricted our search to EU/UK/EEA/US and OECD countries, the studies primarily refer to high-income countries. Finally, as costs and resources varied between different countries, different pandemic settings and over time, and as indicated in this review, dependent on multiple other factors including population vaccination status, preexisting healthcare capacity and the infectivity of the COVID-19 variant at each time point, the comparison of cost-effectiveness measures is a complex process to interpret and the cost effectiveness of each intervention should be weighed by policymakers against the regional circumstances. Moreover, as the complete economic and health consequences are yet unknown, further research is needed on the cost-effectiveness of NPIs, while the cost of the long-term effects of COVID-19 (both physical and mental) should be also assessed in future analyses.

## CONCLUSION

This systematic review assessed economic evaluation studies concerning SARS-CoV-2 and the cost-effectiveness of strategies to prevent and mitigate virus spread within the EU/UK/EEA and OECD country context. Results of this study are based primarily on first wave of the pandemic and mostly from a health care perspective with a short time horizon; particularly within 1 year time horizon. Our review showed that SARS-CoV-2 is associated with substantial economic costs to healthcare systems, payers, and societies, both short term and long term, while interventions including testing and screening policies, vaccination and physical distancing policies were identified as those presenting cost-effective options to deal with the pandemic – dependent on population vaccination and the R_e_ at the stage of the pandemic. Policymakers could benefit from these findings as they indicate the value of both pharmaceutical and non-pharmaceutical interventions to mitigate and respond to the ongoing COVID-19 pandemic and in preparation for future respiratory pandemics.

## Data Availability

All data produced in the present work are contained in the manuscript

## ACKNOWLEDGEMENTS

We would like to thank Chrysa Chatzopoulou, Katerina Papathanasaki and Konstantinos Skouloudakis for contributing to file and data archiving and data management.

## CONTRIBUTORS

CV, JLB, and JES designed the study. KN and KZ undertook the systematic review and extracted the data with help from KA, IL and KPA. JLB and RP developed the search strategy and JLB ran the searches. KZ and KA analysed and interpreted the economic data. OC, FL, FS, AP, CD, EF and JES participated in data evaluation and interpretation along with CV, JLB, RP, JES, KN, KZ, KA, and ST. Authors CV, KA and KZ wrote the first draft of the manuscript with input from all authors. All authors reviewed and revised subsequent drafts.

## Declaration of interests

We declare no competing interests.

## Funding

This report was commissioned by the European Centre for Disease Prevention and Control (ECDC), to the PREP-EU Consortium, coordinated by Dr. Vardavas under specific contract ECD. 11986 within Framework contract ECDC/2019/001. The information and views in this manuscript are those of the authors and do not necessarily reflect the official opinion of the Commission/Agency. The Commission/Agency do not guarantee the accuracy of the data included in this study. Neither the Commission/Agency nor any person acting on the Commission’s/Agency’s behalf may be held responsible for the use which may be made of the information contained therein.

## Data sharing statement

Data sharing is not applicable to this article as no new data were created or analyzed in this study.

## Online Supplementary Appendix 1. Search Terms and Strategy

Database: Ovid MEDLINE(R) ALL <1946 to April 22, 2021>

Search Strategy:

--------------------------------------------------------------------------------

1 Economics/ (27312)

2 “costs and cost analysis”/ (49453)

3 Cost allocation/ (2008)

4 Cost-benefit analysis/ (84093)

5 Cost control/ (21571)

6 Cost savings/ (12170)

7 Cost of illness/ (28590)

8 Cost sharing/ (2590)

9 “deductibles and coinsurance”/ (1780)

10 Medical savings accounts/ (540)

11 Health care costs/ (41105)

12 Direct service costs/ (1203)

13 Drug costs/ (16530)

14 Employer health costs/ (1094)

15 Hospital costs/ (11434)

16 Health expenditures/ (21201)

17 Capital expenditures/ (1996)

18 Value of life/ (5745)

19 exp economics, hospital/ (25059)

20 exp economics, medical/ (14256)

21 Economics, nursing/ (4002)

22 Economics, pharmaceutical/ (2982)

23 exp “fees and charges”/ (30654)

24 (low adj cost).mp. (64981)

25 (high adj cost).mp. (15874)

26 (health?care adj cost$).mp. (12946)

27 (fiscal or funding or financial or finance).tw. (158630)

28 (cost adj estimate$).mp. (2411)

29 (cost adj variable).mp. (46)

30 (unit adj cost$).mp. (2677)

31 (economic$ or pharmacoeconomic$ or price$ or pricing).tw. (327155)

32 Economic evaluation.mp. (10756)

33 (Cost?effectiveness analysis or CEA).mp. (24032)

34 (Cost?utility analysis or CUA).mp. (1284)

35 (Cost?benefit analysis or CBA).mp. (27076)

36 (Cost?consequence analysis or CCA).mp. (9017)

37 (Cost?minimi?sation analysis or CMA).mp. (4218)

38 (cost?outcome or marginal analysis).mp. (225)

39 exp Cost benefit analysis/ or exp budgets/ (97196)

40 investment$.mp. or investments/ (45688)

41 or/1-40 (873724)

42 exp Coronavirus/ (68433)

43 exp Coronavirus Infections/ (82961)

44 (Coronavir* or nCov or covid or covid-19 or Middle East Respiratory Syndrome or MERS or Severe Acute Respiratory Syndrome or SARS).ti,ab,kf. (145090)

45 42 or 43 or 44 (152400)

46 41 and 45 (8520)

47 limit 46 to yr=“2020-Current” (7555)

48 limit 47 to english language (7380)

***************************

Database: Embase <1974 to 2021 April 22>

Search Strategy:

--------------------------------------------------------------------------------

1 Socioeconomics/ (143956)

2 Cost benefit analysis/ (86950)

3 Cost effectiveness analysis/ (158695)

4 Cost of illness/ (19775)

5 Cost control/ (70213)

6 Economic aspect/ (116407)

7 Financial management/ (115358)

8 Health care cost/ (195273)

9 Health care financing/ (13885)

10 Health economics/ (33536)

11 Hospital cost/ (22362)

12 (fiscal or financial or finance or funding).tw. (222499)

13 Cost minimization analysis/ (3660)

14 (cost adj estimate$).mp. (3676)

15 (cost adj variables$).mp. (221)

16 (unit adj cost$).mp. (4828)

17 investment$.mp. or investments/ (57568)

18 or/1-17 (1008351)

19 exp coronavirus/ (34348)

20 exp coronavirus infections/ (116308)

21 (Coronavir* or nCov or covid or Middle East Respiratory Syndrome or MERS or Severe Acute Respiratory Syndrome or SARS).ti,ab,tw. (144234)

22 19 or 20 or 21 (156925)

23 18 and 22 (6304)

24 limit 23 to yr=“2020-Current” (5439)

25 limit 24 to english language (5348)

***************************

## Online Supplementary Appendix 2. Quality appraisal of the economic evaluation studies for estimating COVID-19 infection and strategies for preventing/mitigating COVID-19

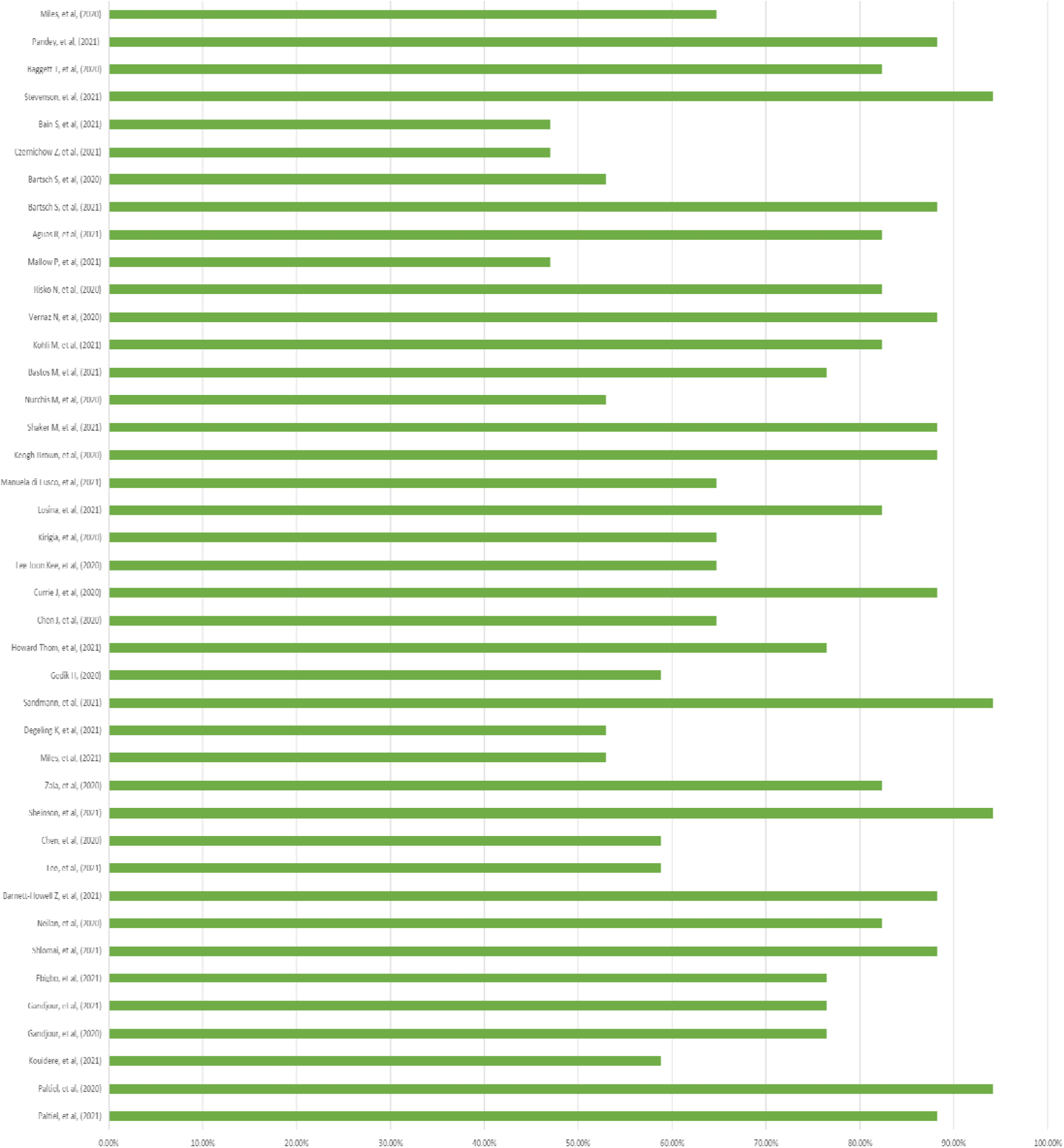

## Notes

### Competing Interest Statement

The authors have declared no competing interest.

